# The causal connections between gut microbiota and chronic pain and pain phenotypes through the gut-brain axis: A Mendelian randomization study and mediation analysis

**DOI:** 10.1101/2024.09.28.24314553

**Authors:** Jia Chen, Xuemei Luo, Luyao Wang, Yiping Luo, Xia Deng, Yuan Tian, Qiao Lu, Genwen Sun, Michael Maes, Xu Zhang

## Abstract

Recent studies have increasingly highlighted a potential link between gut microbiota (GM) and chronic pain. However, existing research has yielded mixed results, leaving many questions unanswered. We employed a two-sample Mendelian randomization design to investigate whether GM directly influences multisite chronic pain (MCP) and eight phenotypes. We then used a combination of meta-analysis, genetic colocalization analysis, and Bayesian weighted mendelian randomization to further strengthen the evidence for causal relationship. Additionally, we examined how GM might be associated with human brain structures. Finally, we used mediation analysis to explore whether the structure and function of specific brain regions play a role in the relationship between GM and MCP and the pain phenotypes.

We observed a significant causal association between 13 GM and chronic pain. Eight GM protected against pain phenotypes, whereas seven increased risk. The results indicate that GM could potentially affect the area, thickness, and volume of brain region cortices involved in sensory, emotional, and cognitive aspects of pain. Importantly, our findings indicated that the causal effect between the Genus *Odoribacter* and neck/shoulder pain could be mediated by the mean optical density in the left Fornix cres+Stria terminalis. In conclusion, different pain phenotypes were causally associated with different GM among European ancestry. Core GM may influence chronic pain by affecting complex brain networks. Notably, we constructed a basic gut-brain axis model for neck/shoulder pain, involving the Genus *Odoribacter*, left Fornix cres+Stria terminalis, and neck/shoulder pain.

## Introduction

Chronic pain (CP) is a major public health issue with significant global attention. About 30% global population experiences CP [34]. While various causes exist, CP can be idiopathic (with unknown origin) or difficult to categorize pathophysiologically [82]. Notably, CP shows a genetic link, with pain phenotypes exhibiting heritability [49].

The gut microbiota (GM) is the human body’s most complex and densely populated microbial ecosystem [33,76]. Recent research highlights its emerging role in the pathophysiology and regulation of pain [62–63]. The gut-brain axis (GBA) is a complex network of communication channels between the gut and brain [66], disruptions in which might be linked to neurological diseases including pain [27, 66].

Preclinical studies have demonstrated a link between GM and visceral pain [25,53]. Probiotics can lessen visceral pain [80] and might even affect the development of brain regions involved in pain processing [45]. Animal studies further suggest that GM influences various chronic pain conditions, including pain caused by inflammation and nerve damage [40, 62]. Other findings hint at a possible connection to nociceptive pain, complex regional pain syndrome, and headaches [50]. This suggests that GM might directly or indirectly regulate neuronal excitability in the peripheral nervous system [61–62, 74].

Despite these findings, the causal effect of GM on pain remains unclear due to potential biases. Mendelian randomization (MR) has been employed to investigate the relationship between GM and specific pain conditions, including colorectal cancer [39], irritable bowel syndrome [42], intervertebral disc degeneration [22], osteoarthritis [38,94], ankylosing spondylitis [90], and back pain [29,78]. Not only are the conclusions of the above-mentioned studies inconsistent, but also pain with unclear pathophysiology is more common in clinical practice. This underscores the need to examine pain phenotypes.

While the microbiota’s role in pain is gaining traction, many questions remain unanswered. In chronic pain, key questions include whether the GM plays a role in chronic pain and different pain phenotypes? How can GBA models be constructed for chronic pain and different pain phenotypes? MR can analyze the potential causal relationship between GM and pain by using genetic variations as instrumental variables (IVs) [37]. Furthermore, mediation analysis [67] can help decompose the impact of GM on pain.

In this study, we aimed to (1) identify potential positive GM affecting multisite chronic pain (MCP) and eight pain phenotypes, (2) analyze the effect of potential positive GM on brain structure, and (3) develop a basic GBA model for MCP and pain phenotypes to provide practical suggestions for clinical practice.

## Materials and methods

### 1.1 Study design

This study investigated the potential cause-and-effect link between GM and chronic pain using a two-sample MR approach. We utilized data on four relevant GM features and assessed their association with both MCP and eight specific pain phenotypes. We further evaluated the relationship between positive exposure and outcomes using meta-analysis and genetic colocalization analysis. Subsequently, we examined the relationship between potential positive GM and brain imaging-derived phenotypes, and brain structure. Finally, we explored whether the structure and function of specific brain regions impacted the relationship between positive GM and MCP, as well as the eight pain phenotypes, through mediation analysis.

## 1. Data sources and instruments

To minimize bias caused by sample overlap, we obtained exposure (GM) and outcome (MCP and pain phenotype) data from independent sources. Table 1 summarizes the data sources and sample sizes. Ethical approval for the original studies was publicly confirmed, and this analysis did not require separate ethical approval.

**Table 1.**
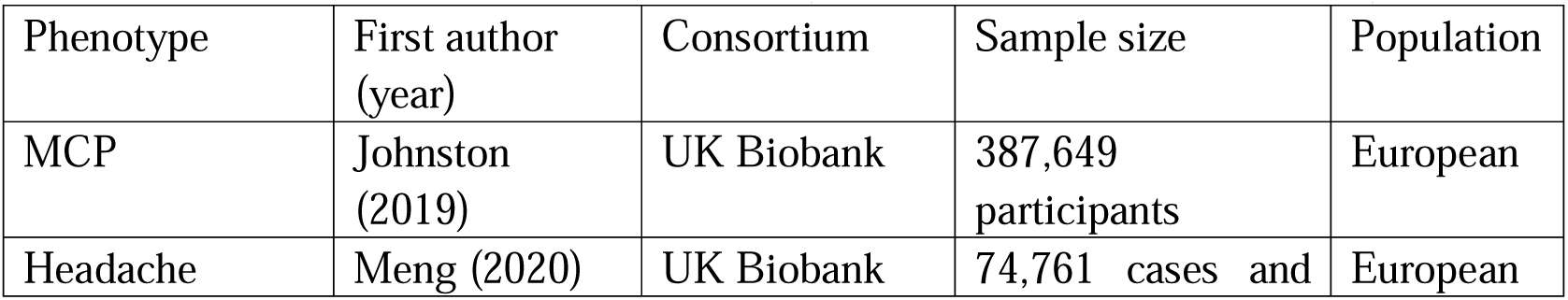
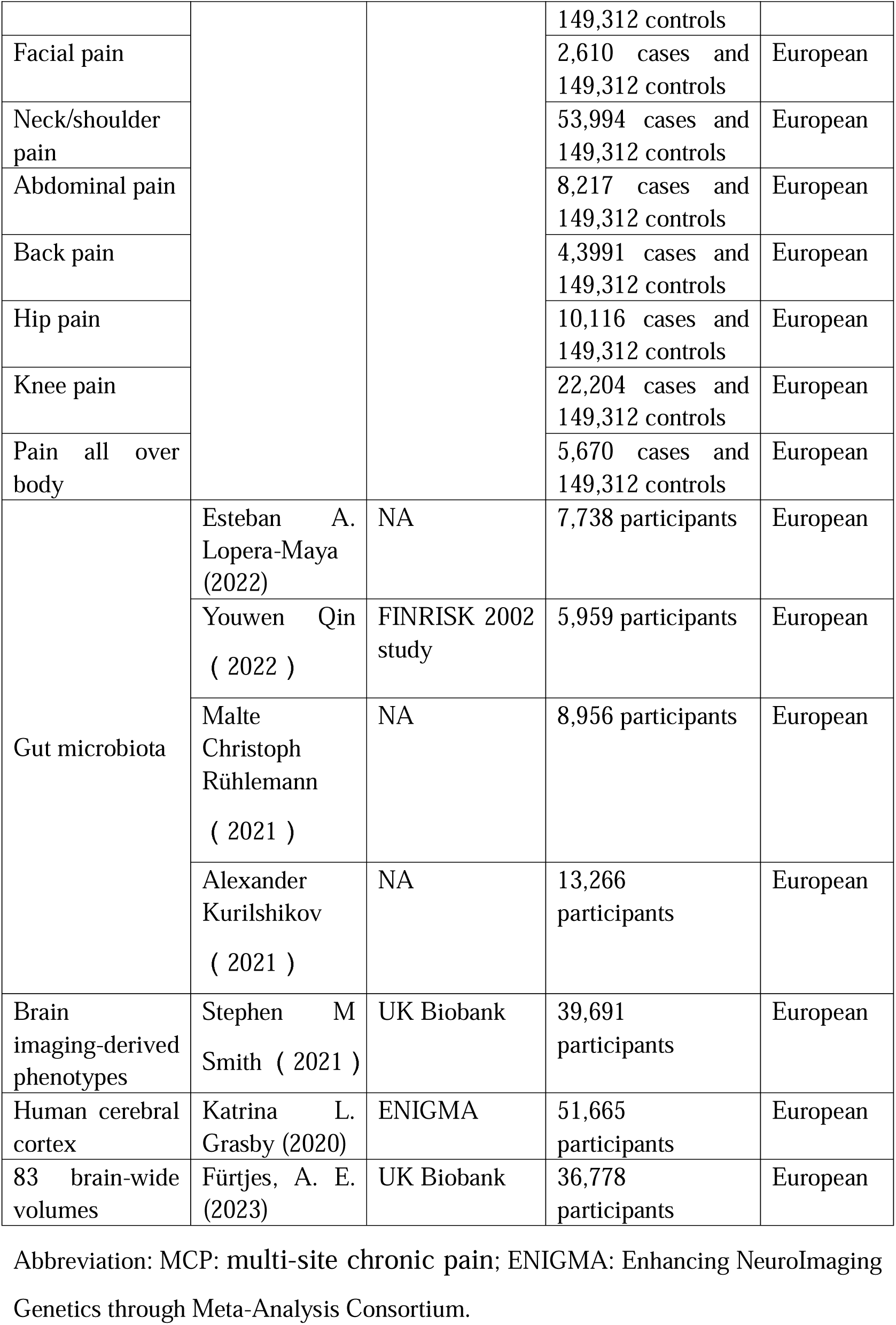
Description of GWAS summary statistics used for each phenotype.

### 2.1 MCP

We obtained data on MCP from the UK Biobank Consortium, a large collection of genetic and health information. This database includes genome-wide association studies (GWAS) for a large population of European ancestry (387,649 individuals) [31]. MCP referred to chronic pain lasting 3 months or more in the head, face, neck/shoulder, back, stomach/abdomen, hip, and knee.

### 2.2 Eight pain phenotypes

Data on eight individual pain conditions were collected from UK Biobank participants (ranging from 151,922 to 226,683 participants depending on the specific pain type) [49]. A validated questionnaire was utilized to assess participants’ experiences with pain in the past month that impacted their daily activities, including pain in the head, face, neck/shoulder pain, back, stomach/abdomen, hips, knees, and all over the body. We excluded participants who chose “prefer not to say” and focused on pain in seven specific locations (head, neck/shoulder, back, abdomen, face, knee, and hip). Individuals who reported pain in a specific location were classified as cases, regardless of other pain selections. Participants who reported “none of the above” were classified as controls. We also included “pain all over the body” as a potential indicator of chronic widespread pain, which is a distinct condition from localized chronic pain.

### 2.3 GM

Data 1: MiBioGen consortium study (14,306 Europeans): This study used 16S rRNA gene sequencing and analyzed 211 microbial taxa (9 phyla, 16 classes, 20 orders, 35 families, 131 genera) from European participants[36].

Data 2: German single-country GWAS study (8,956 Germans): This study analyzed gut bacteria using 16S rRNA gene sequencing[65]. We examined 233 different bacterial groups (4 phyla, 8 classes, 6 orders, 10 families, 29 genera; 65, 62, and 49 at 97% OTU, 99% OTU, and ASV levels) to understand their abundance. Additionally, we analyzed the presence or absence of 198 bacterial features (2 classes, 1 order, 2 families, 17 genera; 65, 62, and 49 on 97% OTU, 99% OTU, and ASV levels), based on taxonomic annotation or ASV/OTU clustering.

Data 3: Dutch Microbiome Project study (7,738 individuals): This study used shotgun metagenomic sequencing[44] and analyzed 207 microbial taxa (5 phyla, 10 classes, 13 orders, 26 families, 48 genera, and 105 species) and 205 functional pathways.

Data 4: FINRISK 2002 study (5,959 individuals): This study used shotgun metagenomic sequencing [60] and analyzed 471 distinct Genome Taxonomy Database (GTDB) taxa (11 phyla, 19 classes, 24 orders, 62 families, 146 genera, and 209 species).

### 2.4 Brain

#### 2.4.1 Brain imaging-derived phenotypes

Summary statistics for brain imaging-derived (BID) phenotypes were obtained from a large GWAS conducted by Smith et al. [70] using data from the UK Biobank, which included 39,691 European individuals. This dataset comprises nearly 4,000 BID phenotypes encompassing brain structure, function, connectivity, and microstructure.

#### 2.4.2 Human brain cortical data

We leveraged brain magnetic resonance imaging (MRI) data from a large-scale analysis (genome-wide association meta-analysis) involving over 51,000 individuals of European descent across 60 separate studies. This analysis was conducted by a research group within the Enhancing NeuroImaging Genetics through Meta-Analysis (ENIGMA) Consortium. The study focused on two key aspects of the brain’s outer layer (cortex): the total surface area (SA) and average thickness (TH). Additionally, they examined 34 specific regions within the cortex known to have distinct functions, as defined by the Desikan-Killiany atlas [24]. For further details, please refer to the original ENIGMA study. In our analysis, we specifically used data on the SA and TH of these brain regions, accounting for overall brain size and genetic background.

#### 2.4.3 Brain-wide volumes

Researchers developed a technique called genomic principal component analysis (PCA), which allowed them to analyze brain scans and identify general underlying patterns in brain structures (morphometry) across the entire brain; the analysis takes into account the potential underlying genetic influences [21]. This technique was applied to genetic data associated with 83 distinct brain volume measurements in over 36,000 UK Biobank participants. Unlike traditional methods that focus on genetic similarity between individuals, Genomic PCA identifies key genetic components that capture the overall variation in brain volume across different brain regions.

## 3. Genetic variants selection criteria

We identified genetic variations (single nucleotide polymorphisms, SNPs) that strongly influence GM using strict criteria, including a very low P-value (<1×10^-5^) [78], a 10000-kb window around a leading SNP, and minimal linkage disequilibrium (LD, r^2^ < 0.001, kb=10000) with other nearby SNPs to minimize confounding effects. Additionally, the chosen SNPs had a minor allele frequency (MAF) > 0.01 to ensure adequate statistical power. After this initial filtering, we further ensured that the variations were reflective of the effects of exposure on outcome; any SNPs with F-statistics < 10 or failure to harmonize, including those harboring incompatible alleles or palindromic characteristics with intermediate allele frequencies, were excluded. Correct inference direction from exposure to outcome was identified using the Steiger filtering approach, to lessen the influence of reverse association. Finally, we used the NHGRI-EBI Catalog (https://www.ebi.ac.uk/gwas/) to retrieve these SNPs and excluded SNPs related to confounding factors and outcomes (education, living habits, smoking, diet) [97]. Figure 1 summarizes this selection process.

**Figure 1:**
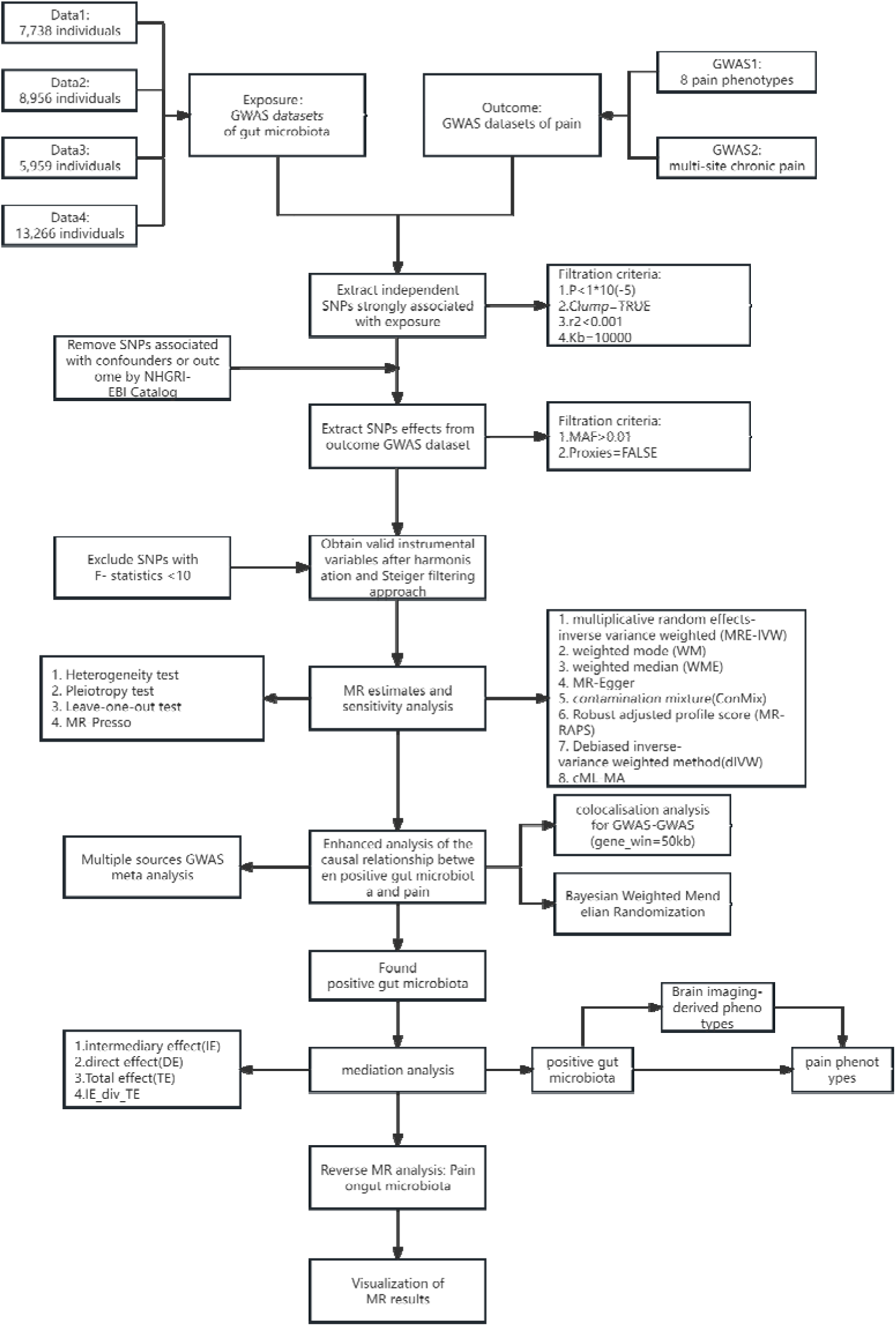
Flow chart of study

**Figure 2a.**
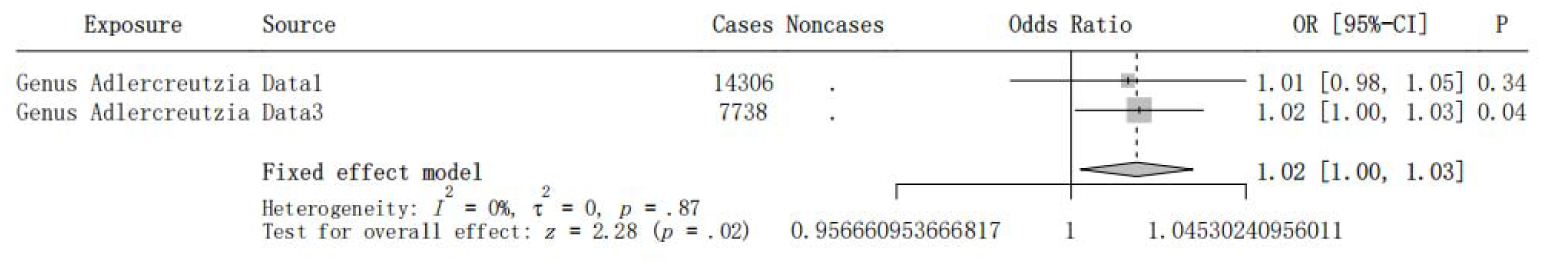
The Forest plots of Meta analysis of Genus Adlercreutzia on MCP

**Figure 2b.**
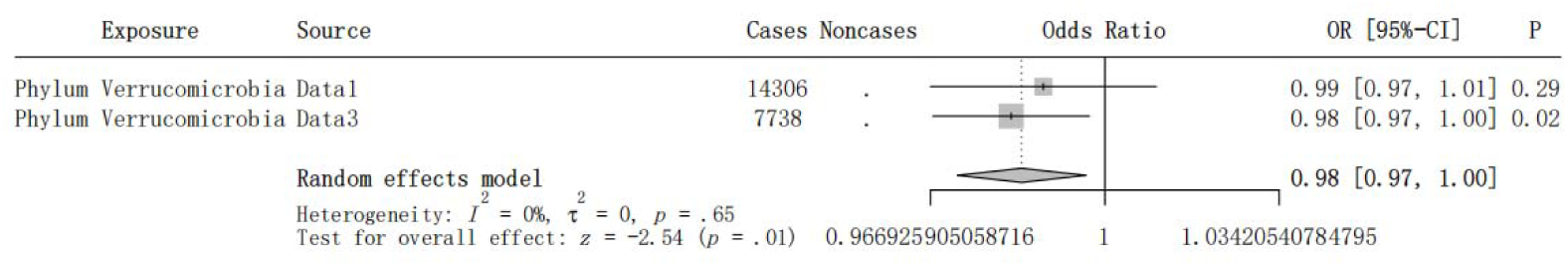
The Forest plots of Meta analysis of Phylum Verrucomicrobia on MCP

**Figure 2c.**
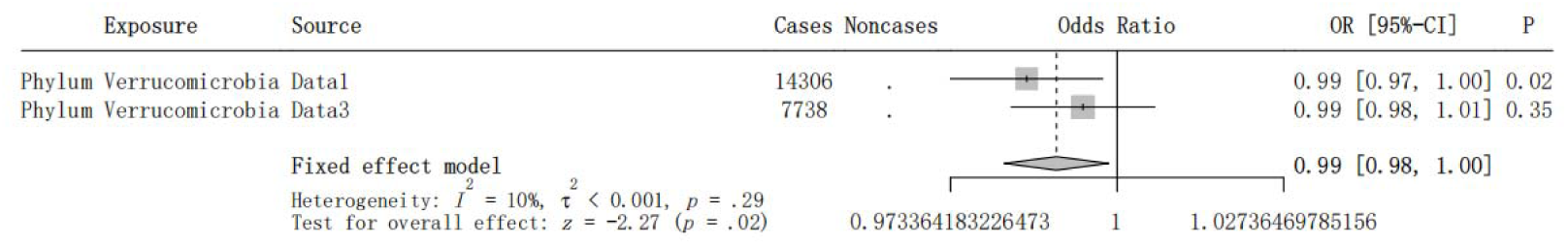
The Forest plots of Meta analysis of Phylum Verrucomicrobia on Headache

**Figure 2d.**
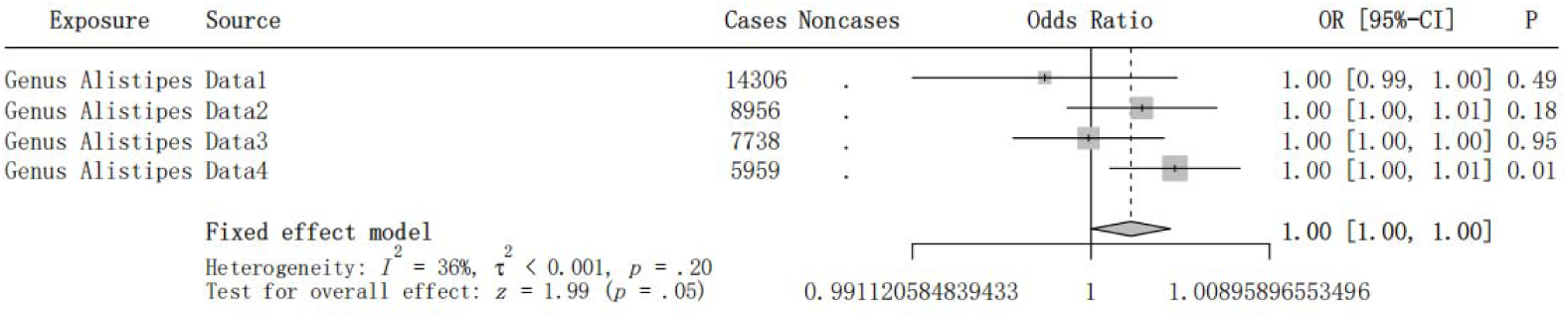
The Forest plots of Meta analysis of Genus Alistipes on Facialpain

**Figure 2e.**
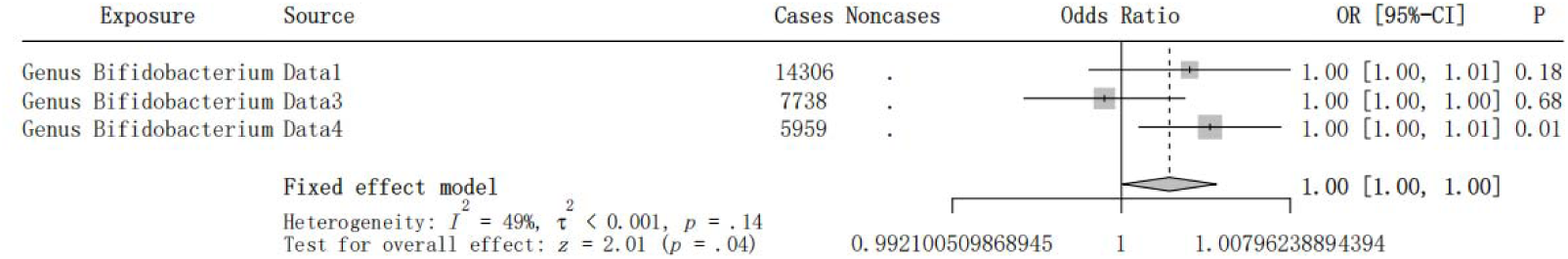
The Forest plots of Meta analysis of Genus Bifidobacterium on Facialpain

**Figure 2f.**
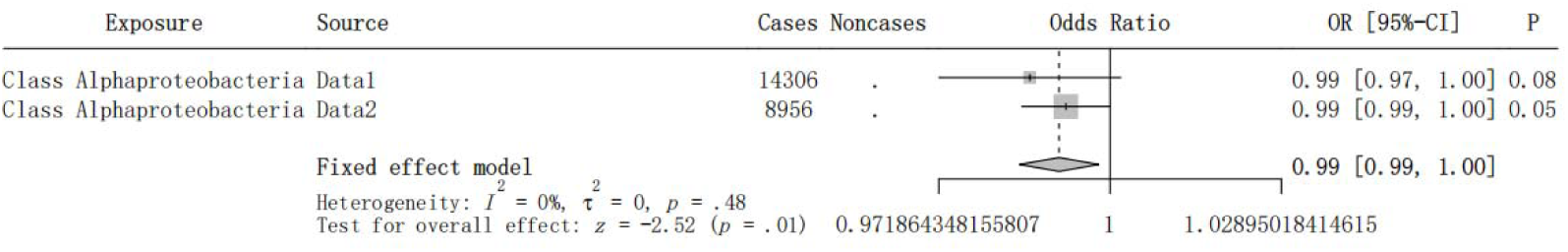
The Forest plots of Meta analysis of Class Alphaproteobacteria on Neckshoulderpain

**Figure 2g.**
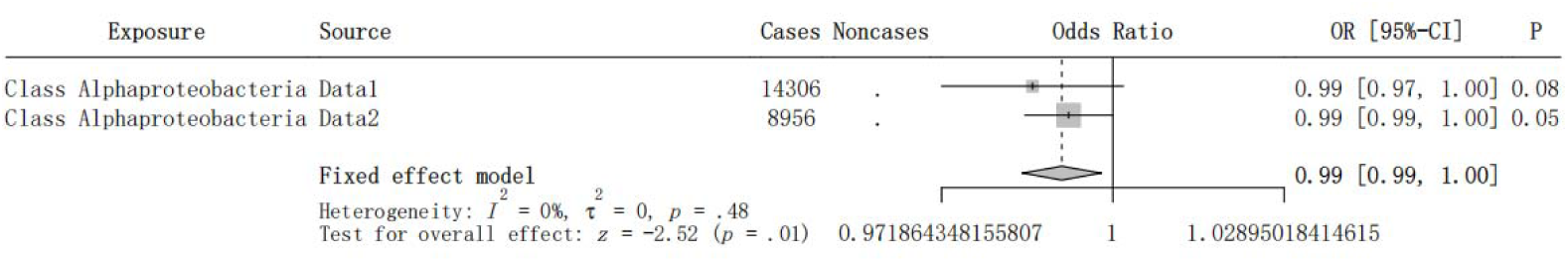
The Forest plots of Meta analysis of Genus Odoribacter on Neckshoulderpain

**Figure 2h.**
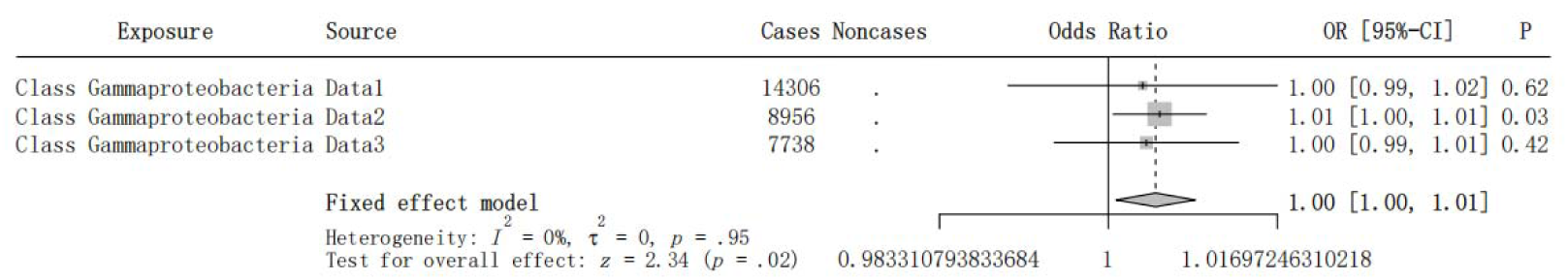
The Forest plots of Meta analysis of Class Gammaproteobacteria on Abdominalpain

**Figure 2i.**
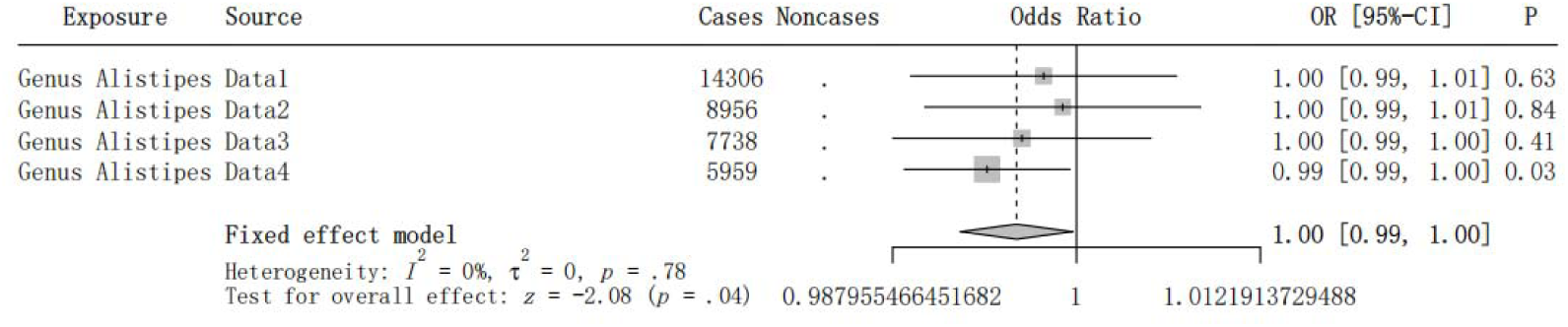
The Forest plots of Meta analysis of Genus Alistipes on Abdominalpain

**Figure 2j.**
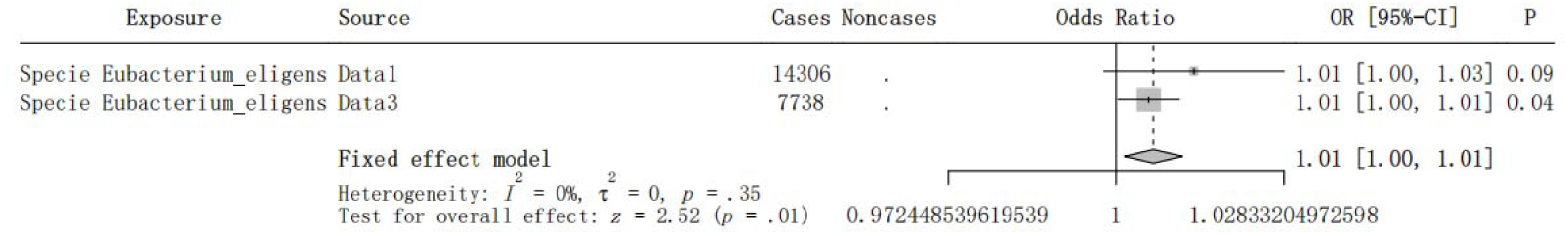
The Forest plots of Meta analysis of Specie Eubacterium_eligens on Abdominalpain

**Figure 2k.**
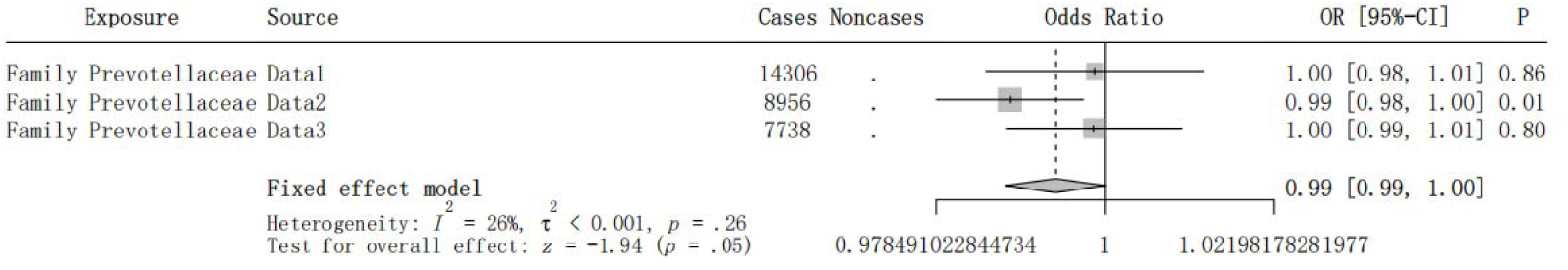
The Forest plots of Meta analysis of Family Prevotellaceae on Backpain

**Figure 2l.**
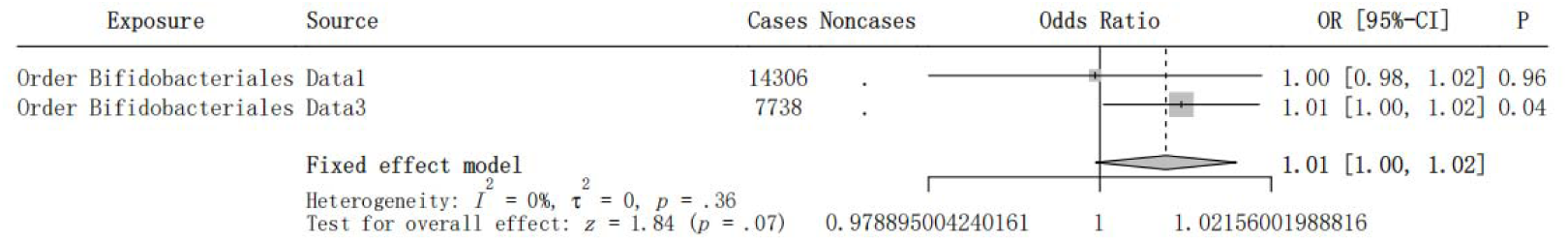
The Forest plots of Meta analysis of Order Bifidobacteriales on Backpain

**Figure 2m.**
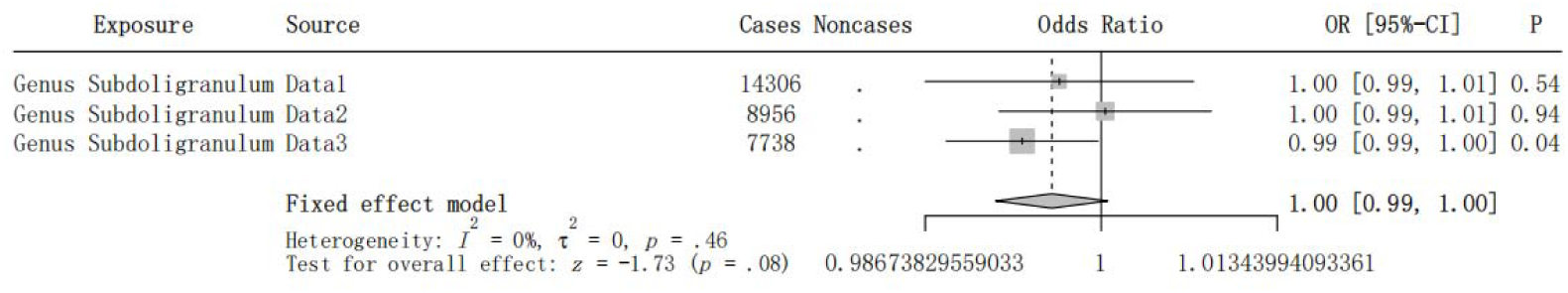
The Forest plots of Meta analysis of Genus Subdoligranulum on Hippain

**Figure 2n.**
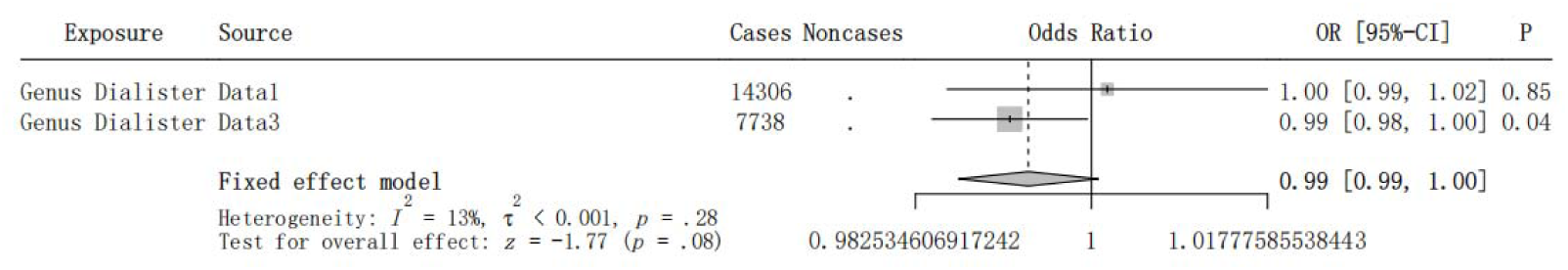
The Forest plots of Meta analysis of Genus Dialister on Kneepain

**Figure 2o.**
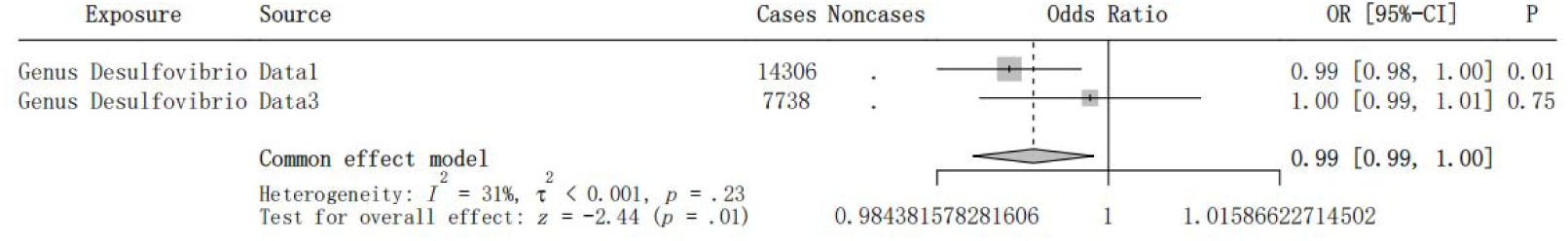
The Forest plots of Meta analysis of Genus Desulfovibrio on pain all over body

**Figure 2p.**
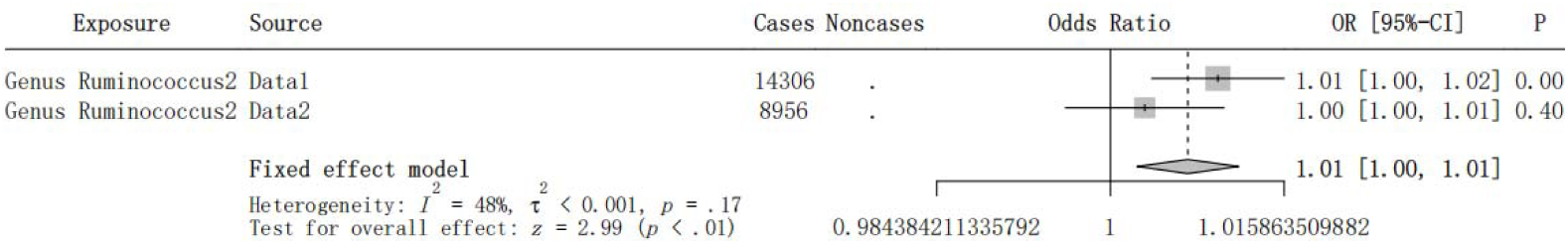
The Forest plots of Meta analysis of Genus Ruminococcus2 on pain all over body

**Figure 3a.**
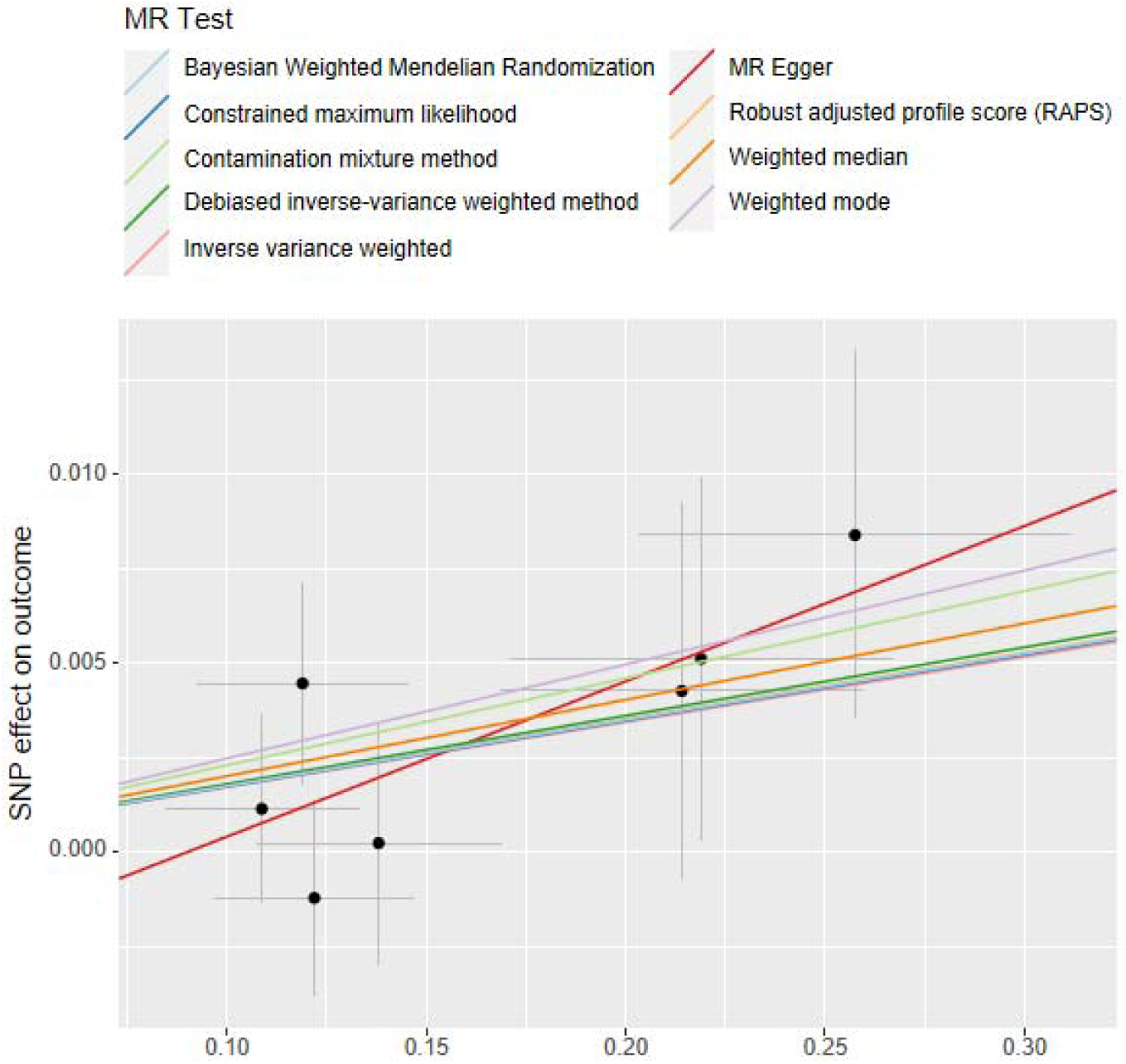
The scatter plots of MR: Genus Adlercreutzia on MCP

**Figure 3b.**
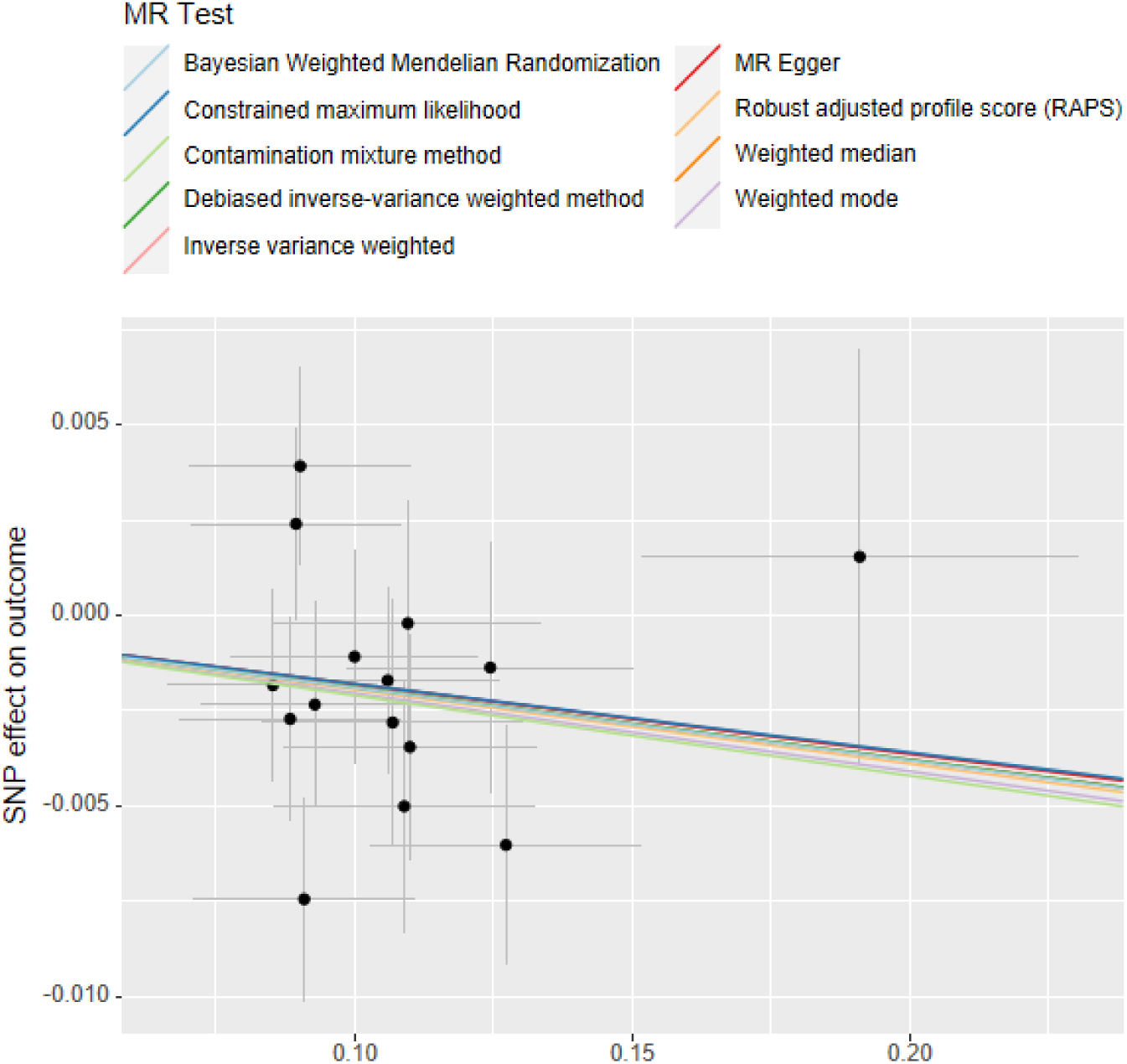
The scatter plots of MR: Phylum Verrucomicrobia on MCP

**Figure 3c.**
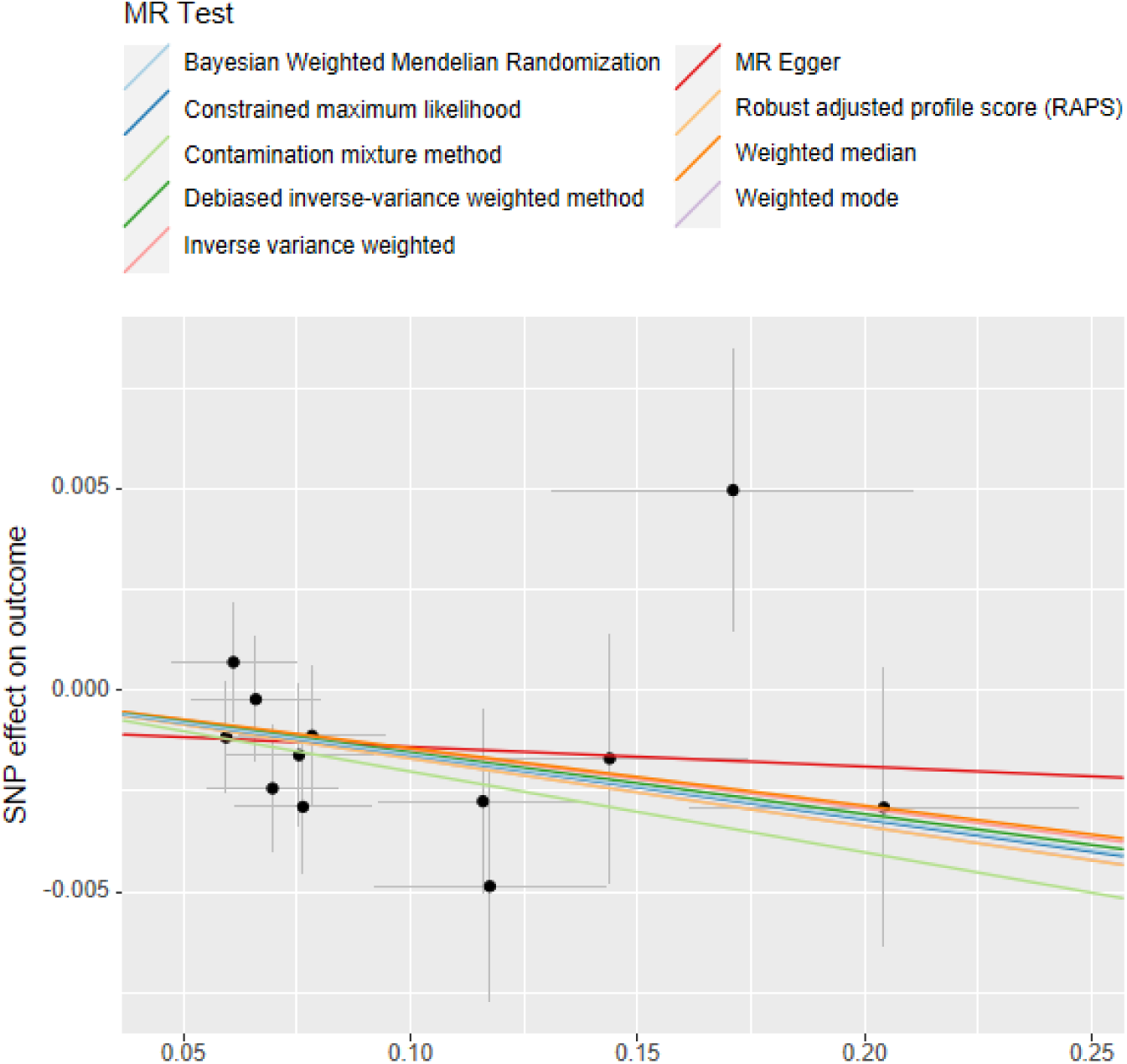
The scatter plots of MR: Phylum Verrucomicrobia on Headache

**Figure 3d.**
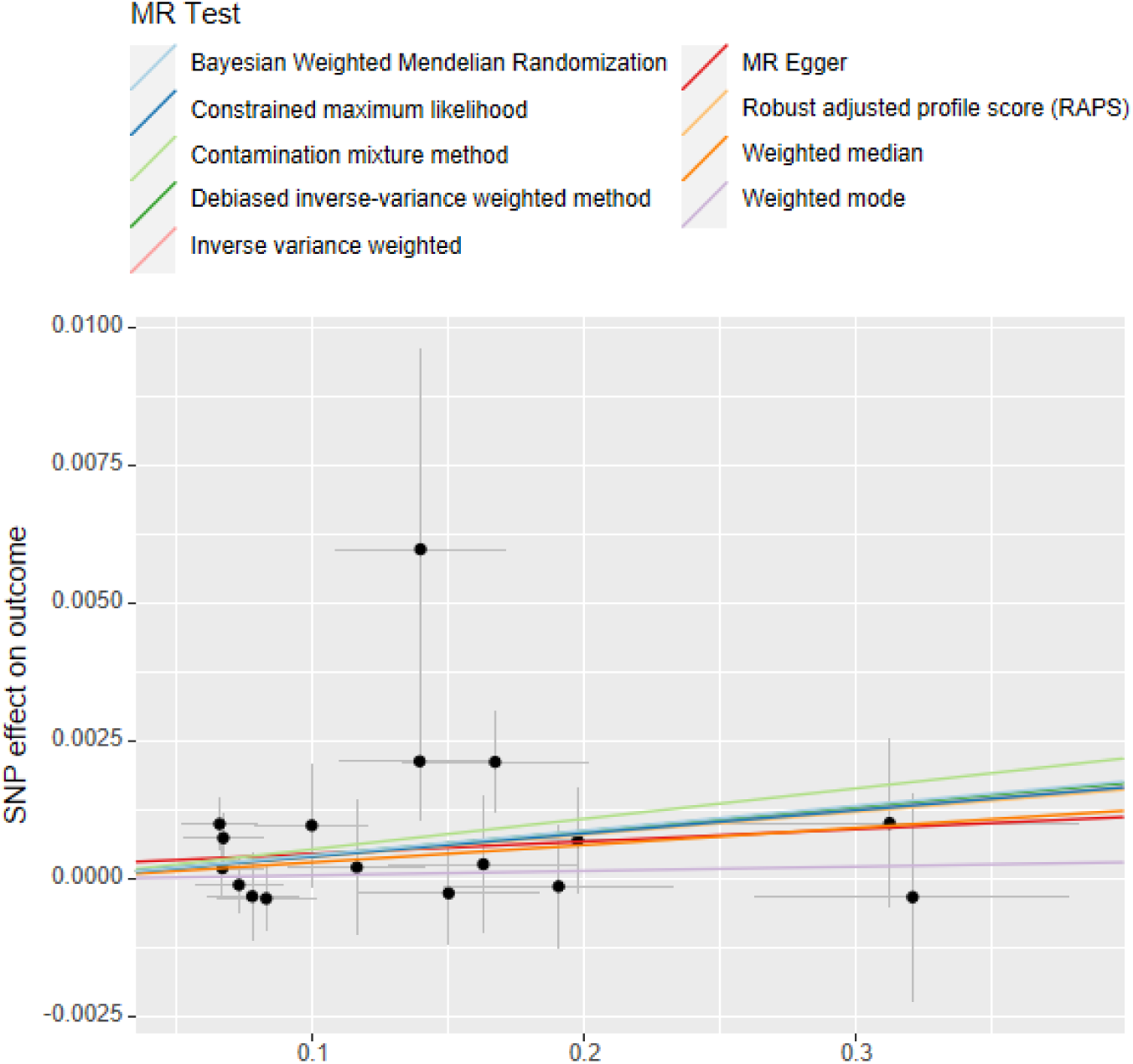
The scatter plots of MR: Genus Alistipes on Facialpain

**Figure 3e.**
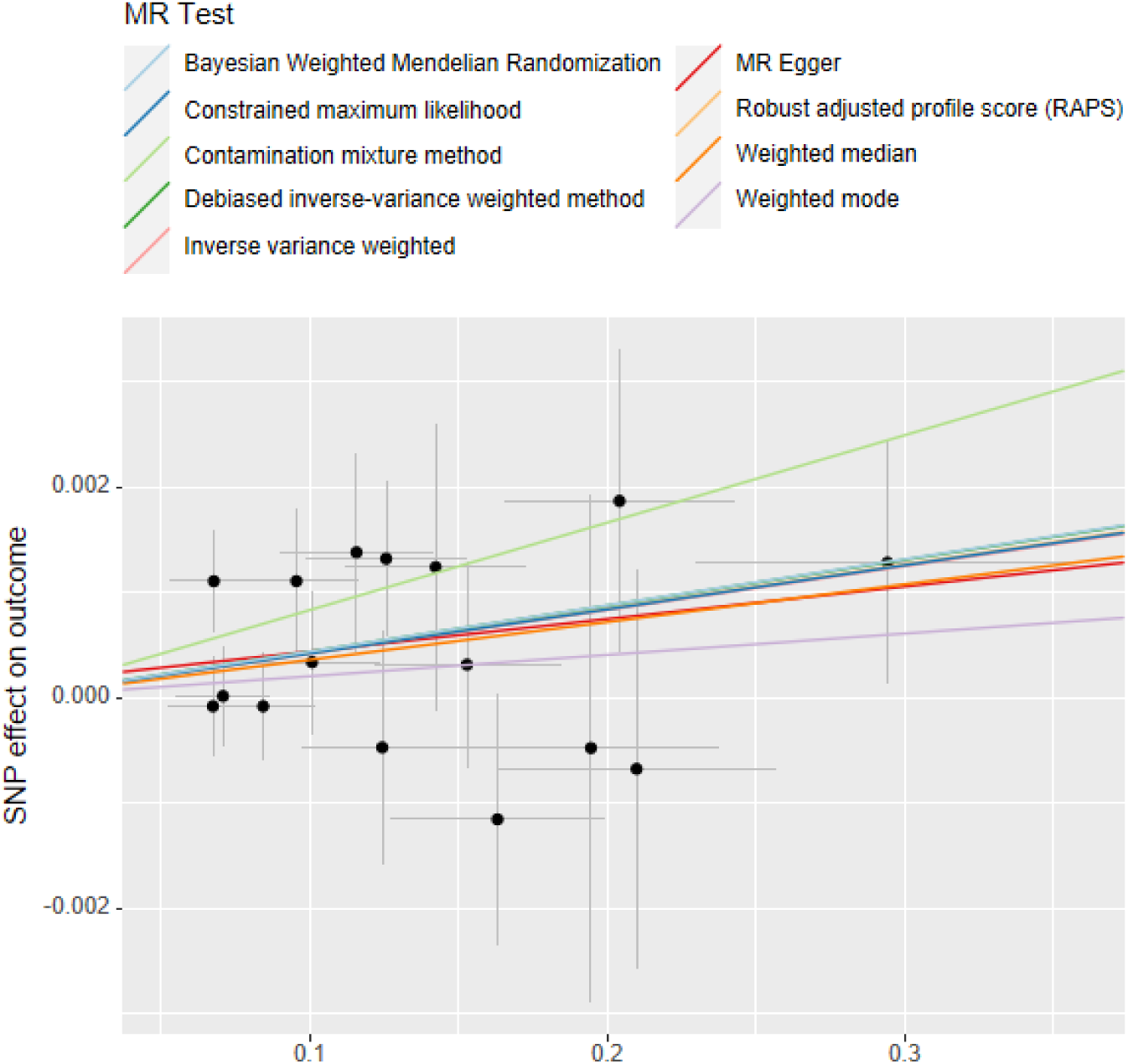
The scatter plots of MR: Genus Bifidobacterium on Facialpain

**Figure 3f.**
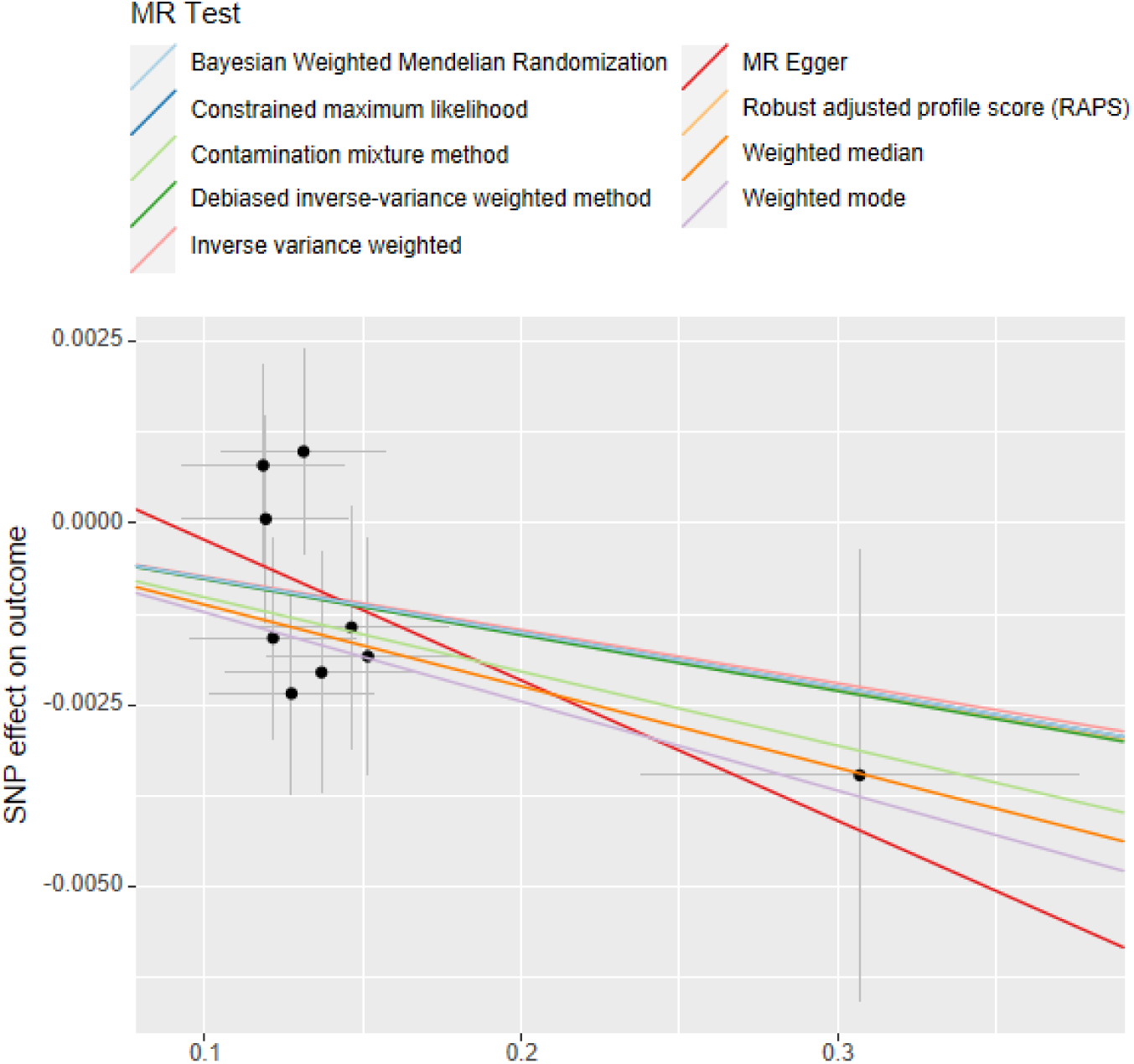
The scatter plots of MR: Class Alphaproteobacteria on Neckshoulderpain

**Figure 3g.**
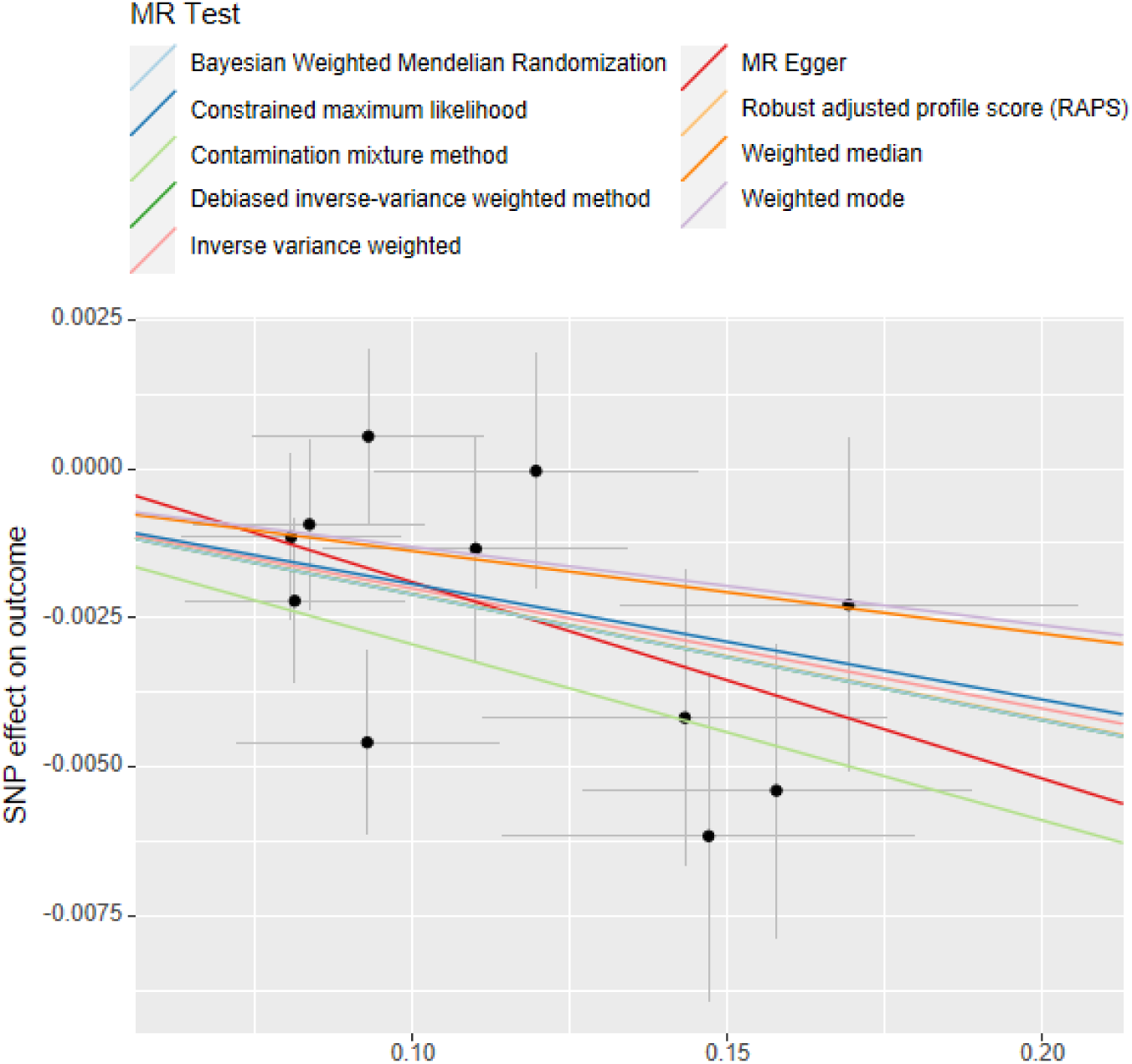
The scatter plots of MR: Genus Odoribacter on Neckshoulderpain

**Figure 3h.**
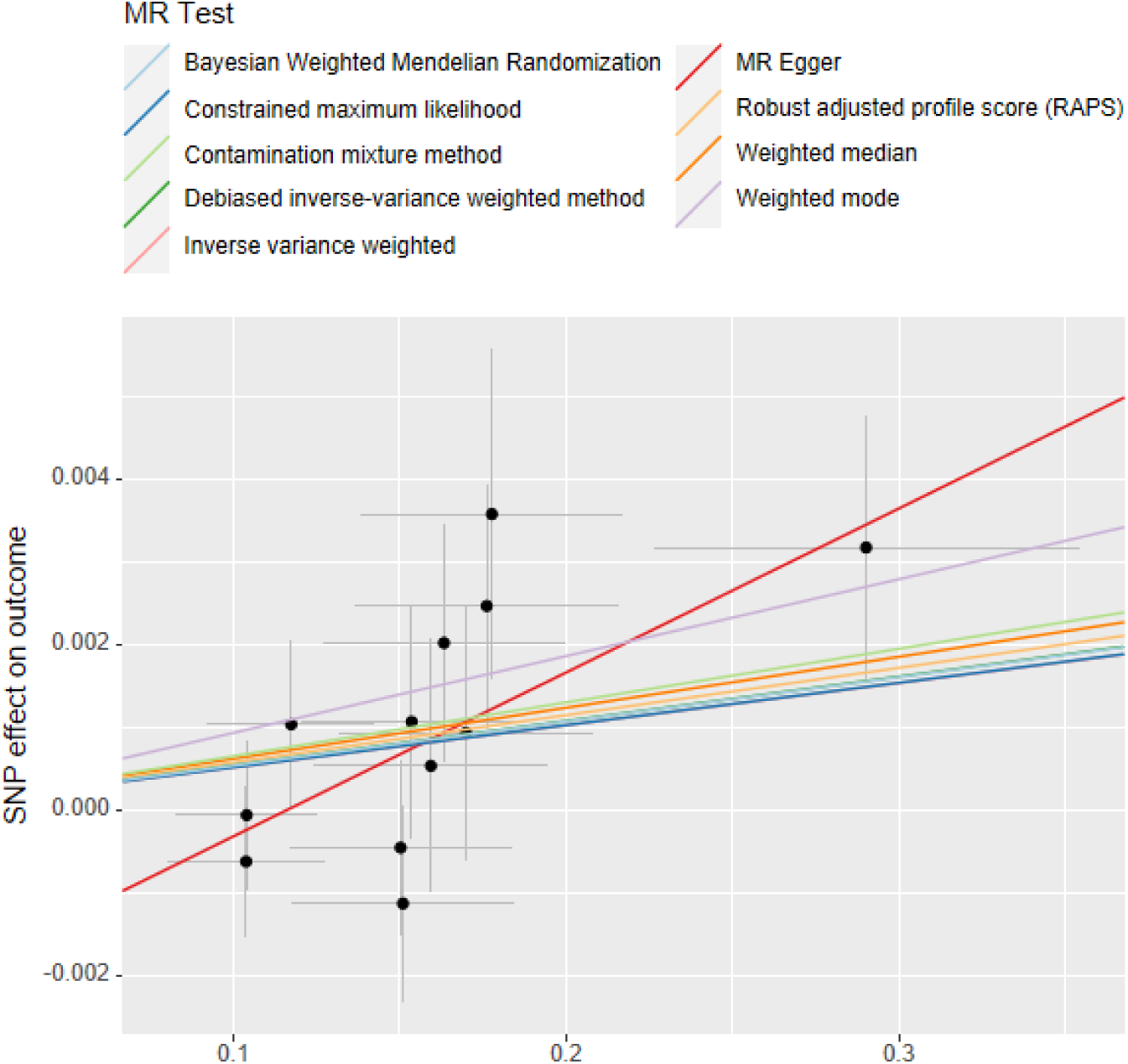
The scatter plots of MR: Class Gammaproteobacteria on Abdominalpain

**Figure 3i.**
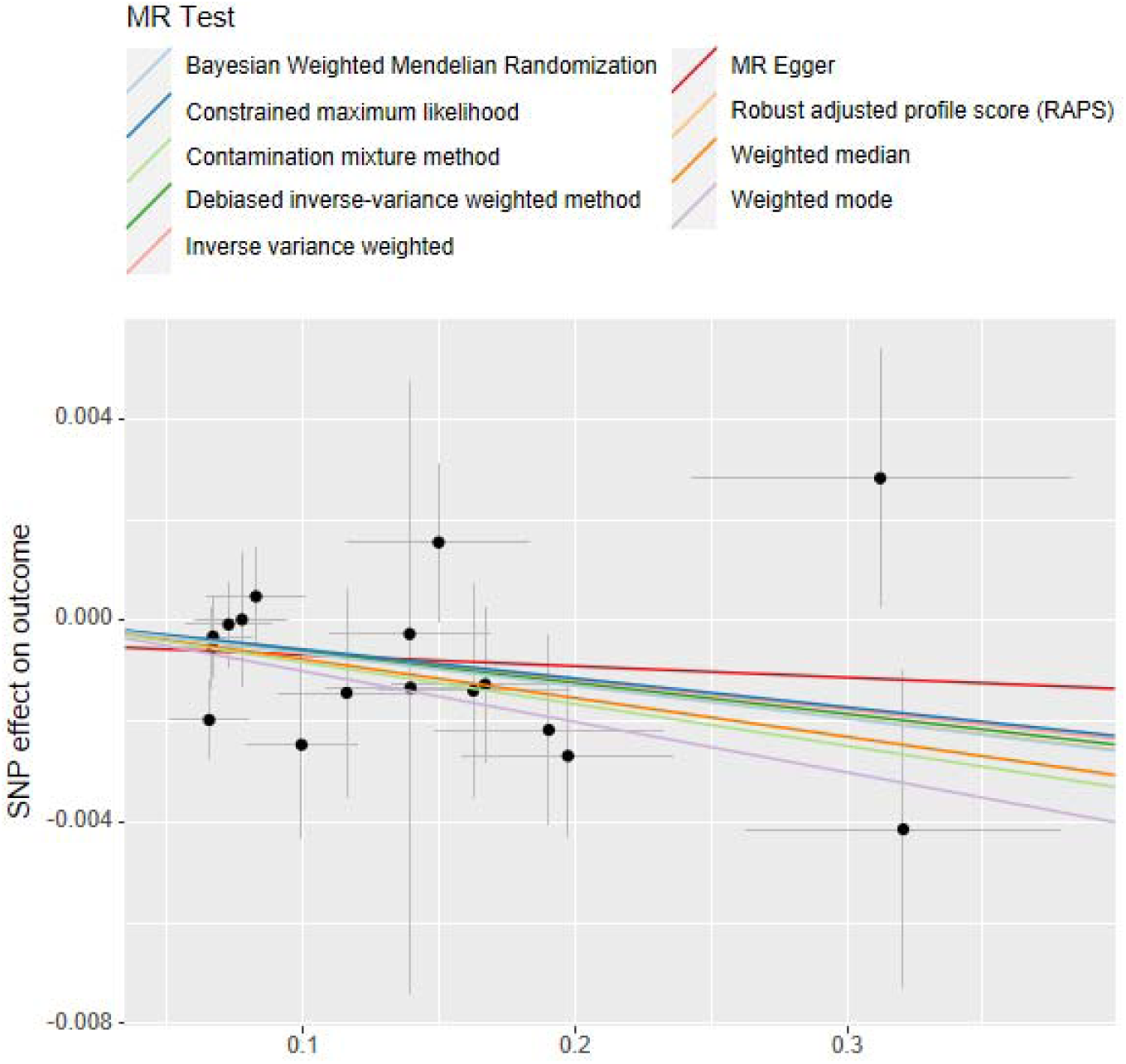
The scatter plots of MR: Genus Alistipes on Abdominalpain

**Figure 3j.**
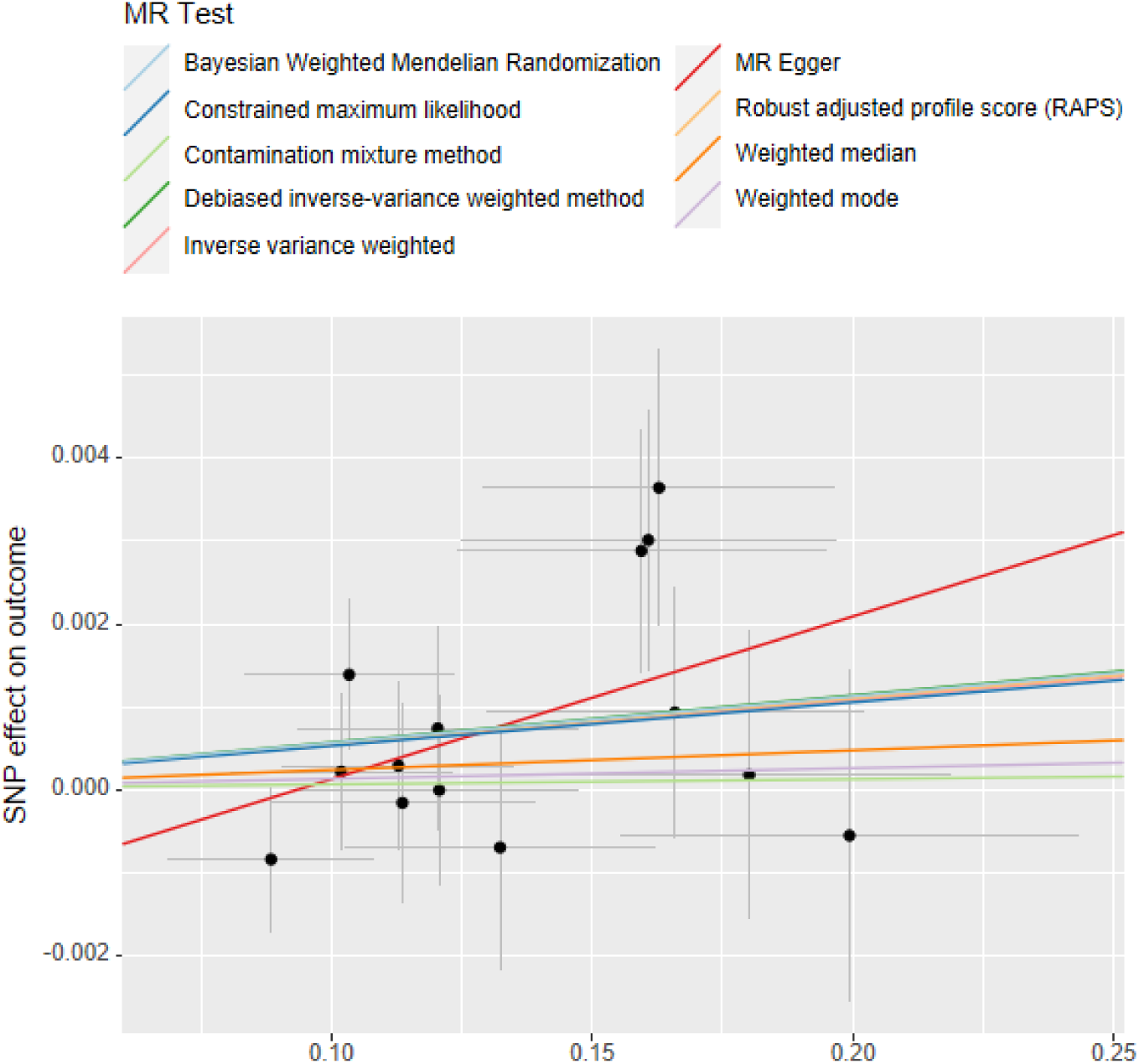
The scatter plots of MR: Specie Eubacterium_eligens on Abdominalpain

**Figure 3k.**
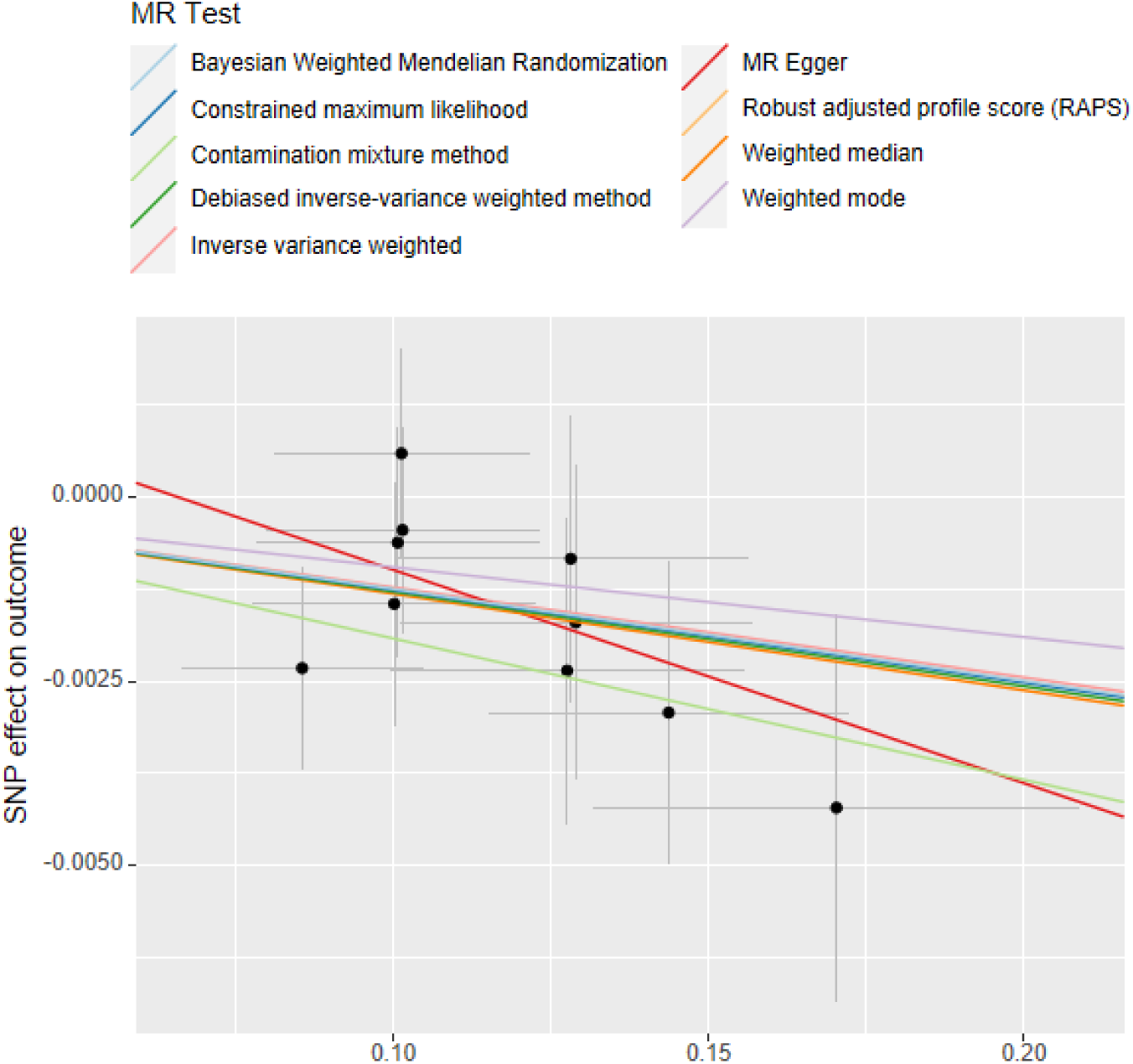
The scatter plots of MR: Family Prevotellaceae on Backpain

**Figure 3l.**
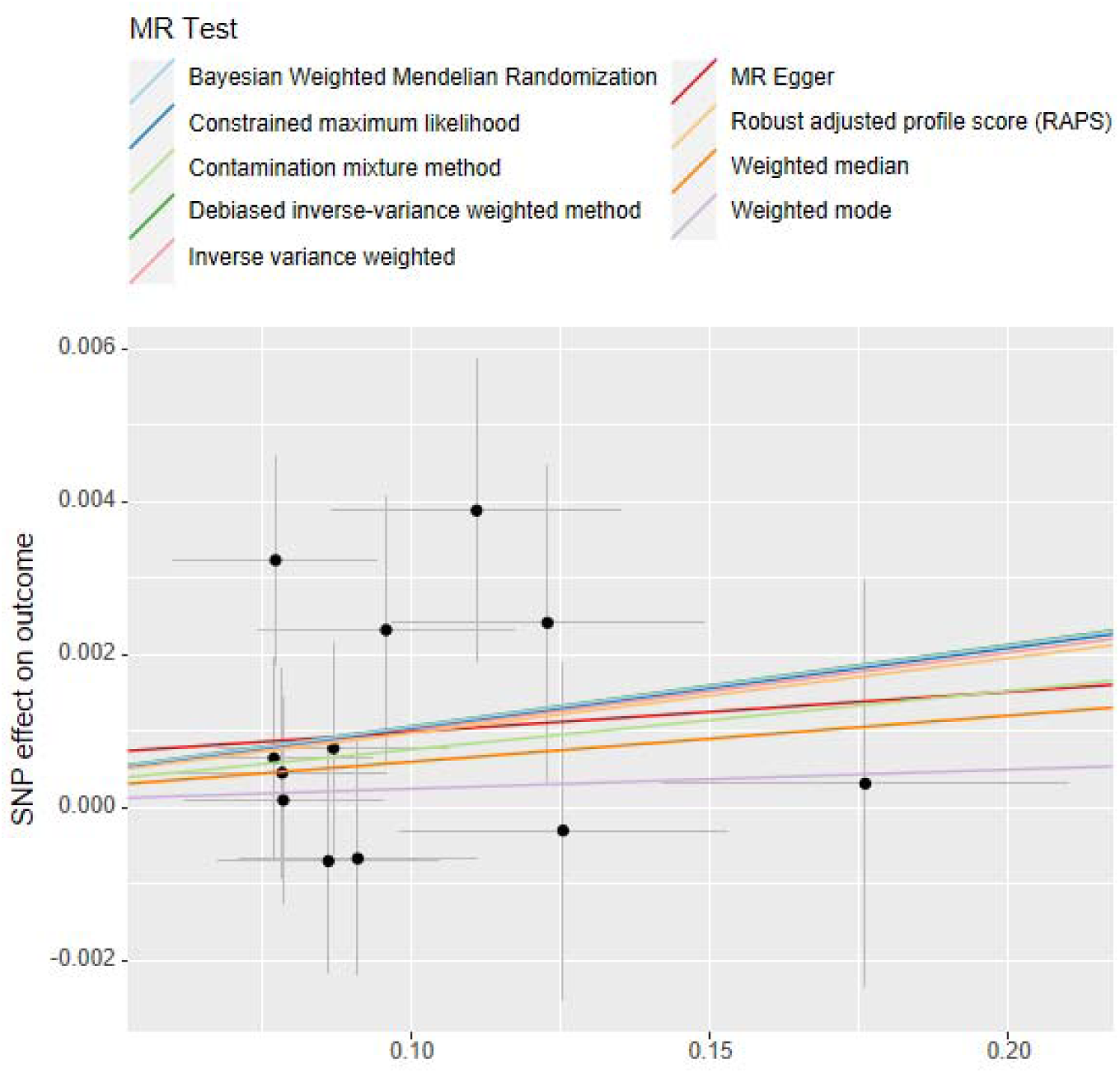
The scatter plots of MR: Order Bifidobacteriales on Backpain

**Figure 3m.**
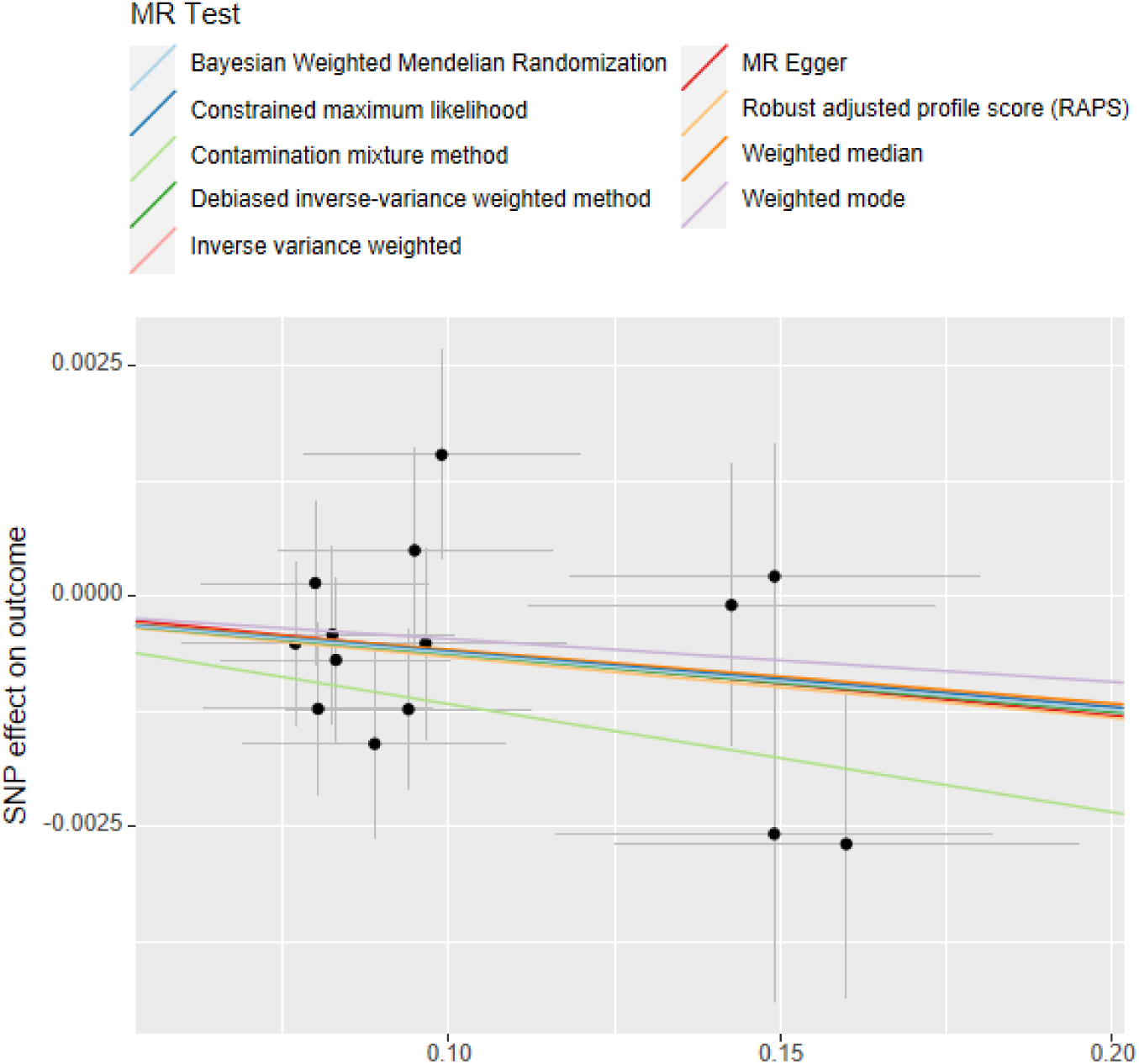
The scatter plots of MR: Genus Subdoligranulum on Hippain

**Figure 3n.**
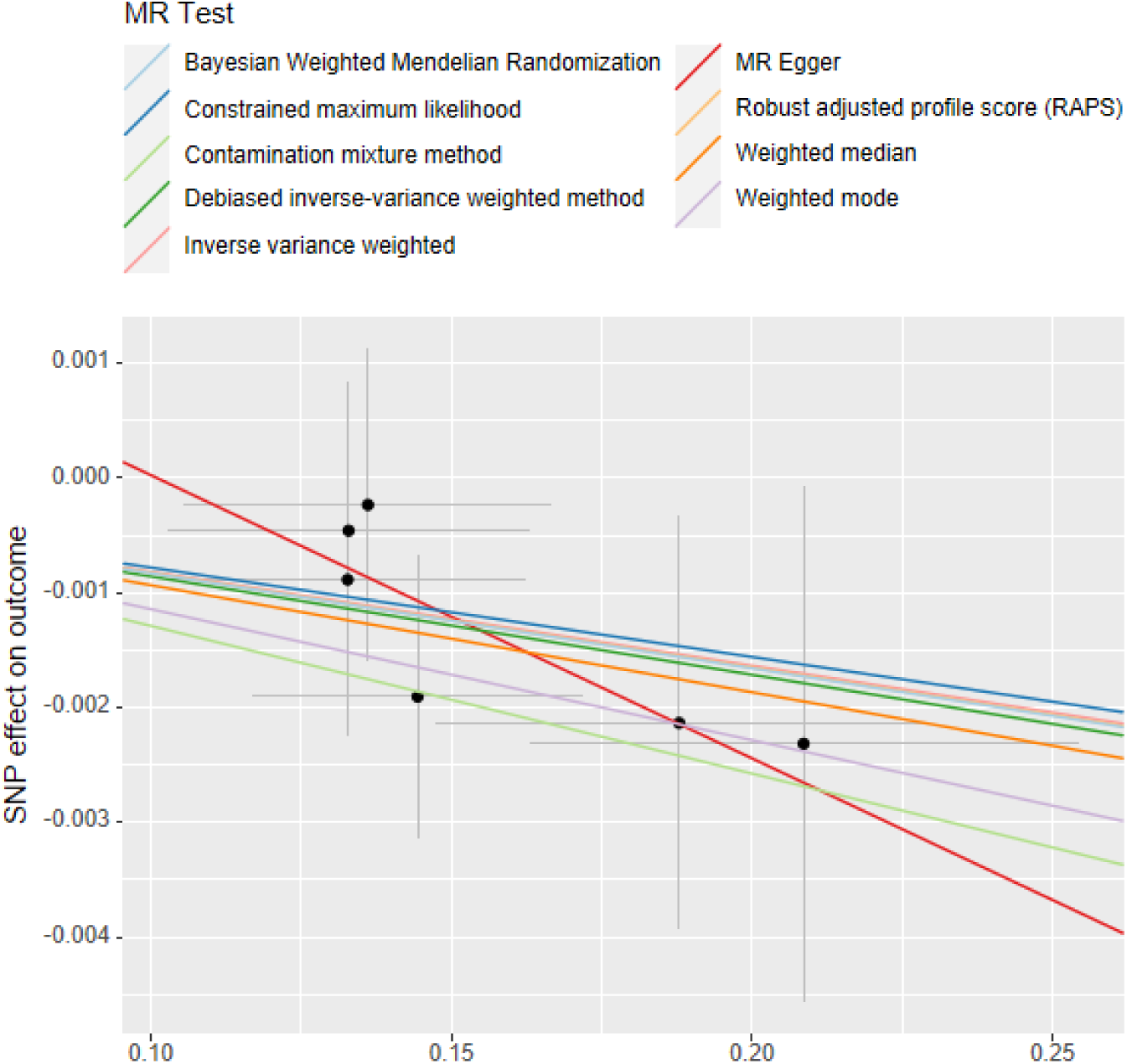
The scatter plots of MR: Genus Dialister on Kneepain

**Figure 3o.**
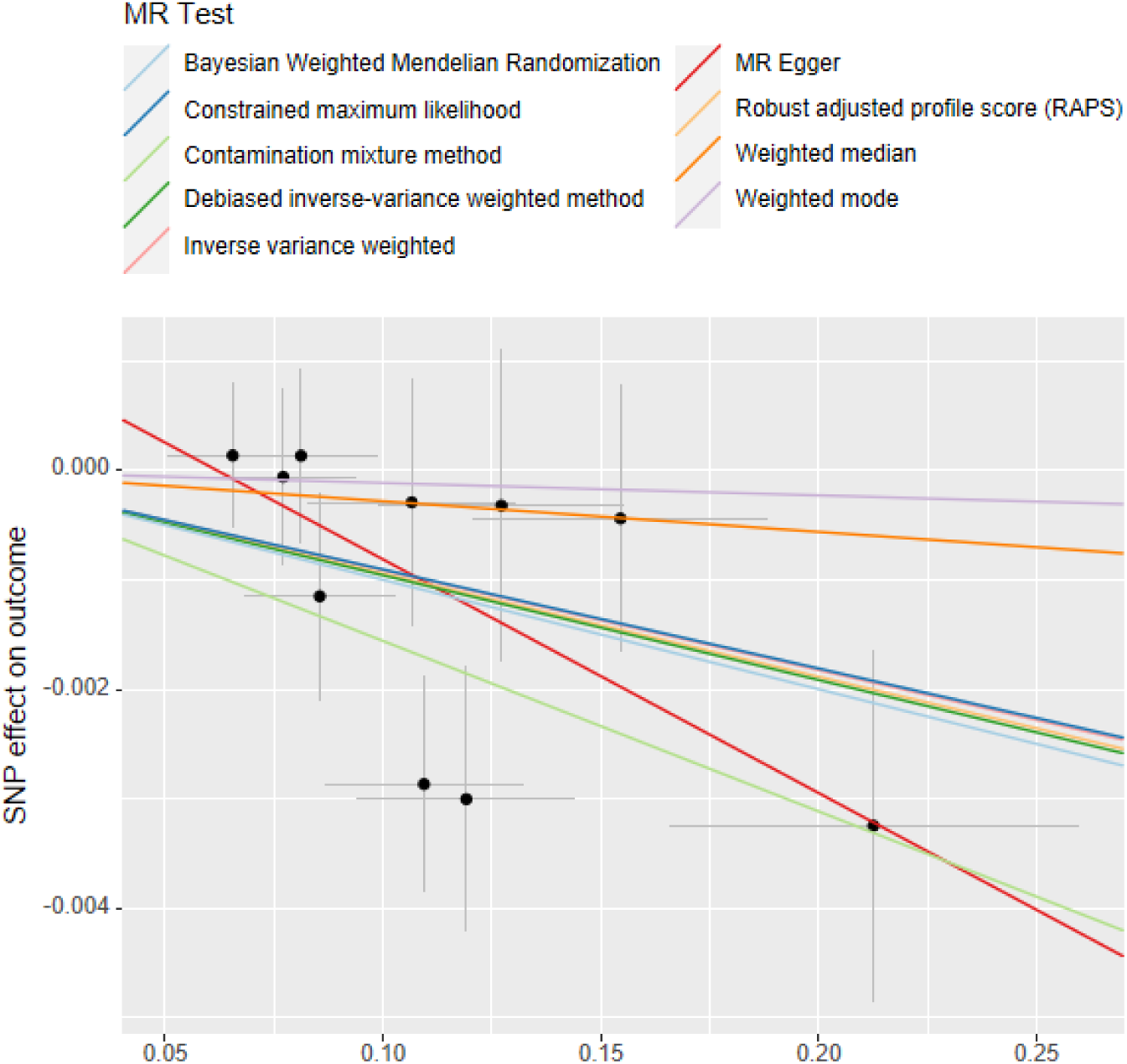
The scatter plots of MR: Genus Desulfovibrio on pain all over body

**Figure 3p.**
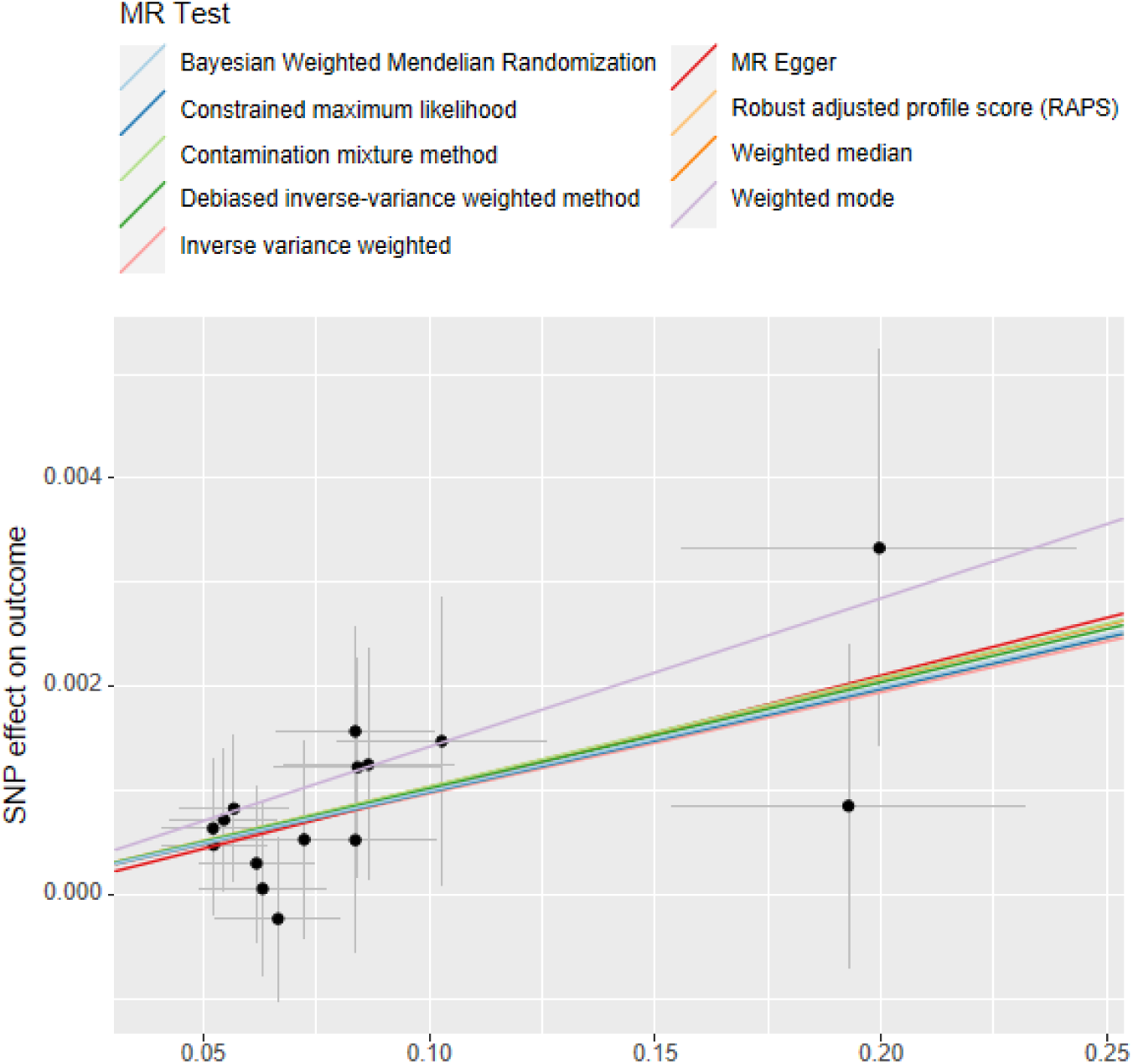
The scatter plots of MR: Genus Ruminococcus2 on pain all over body

**Figure 4a.**
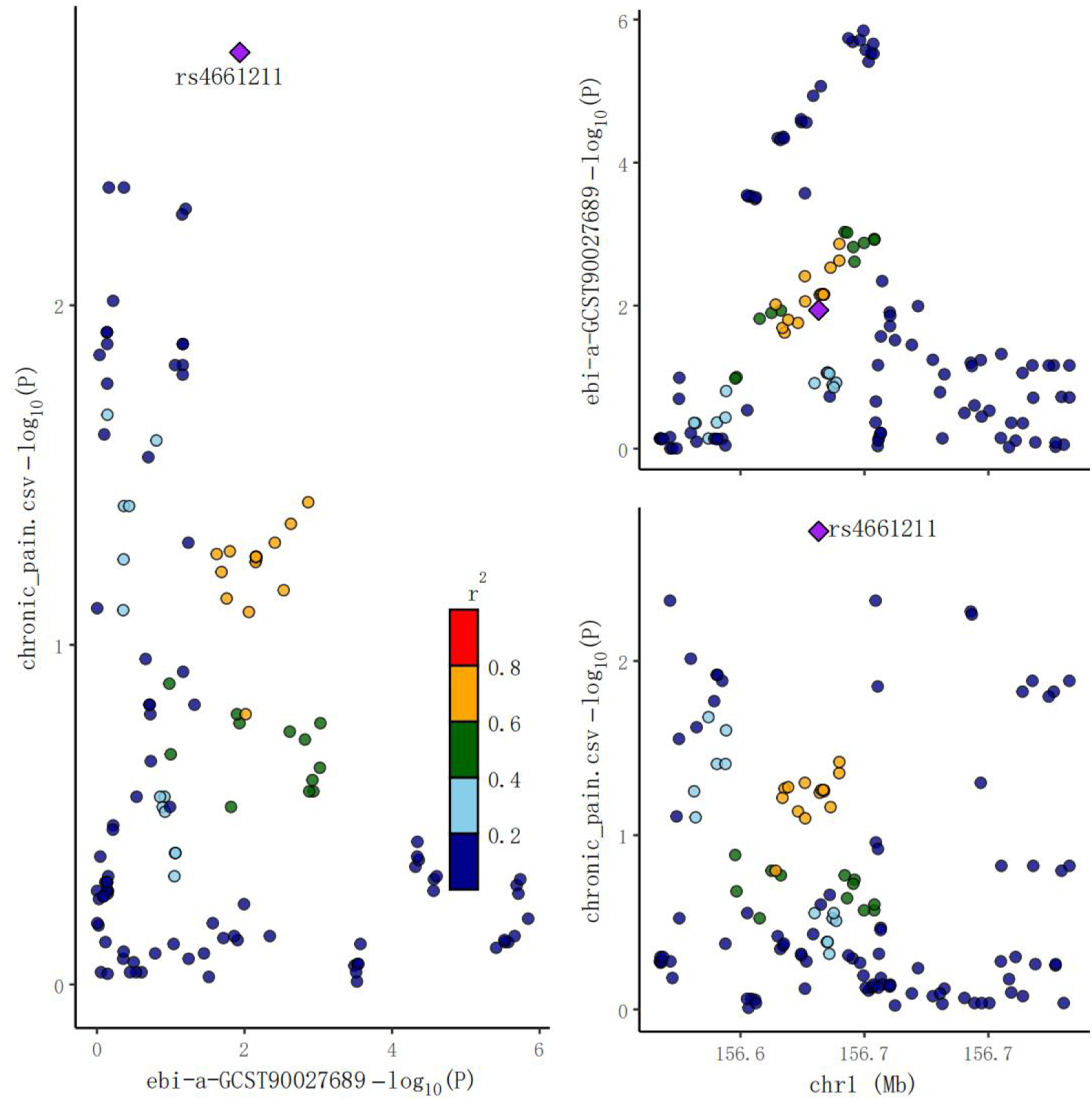
The locuscomparer plot of coloc analysis: Genus Adlercreutzia on MCP

**Figure 4b.**
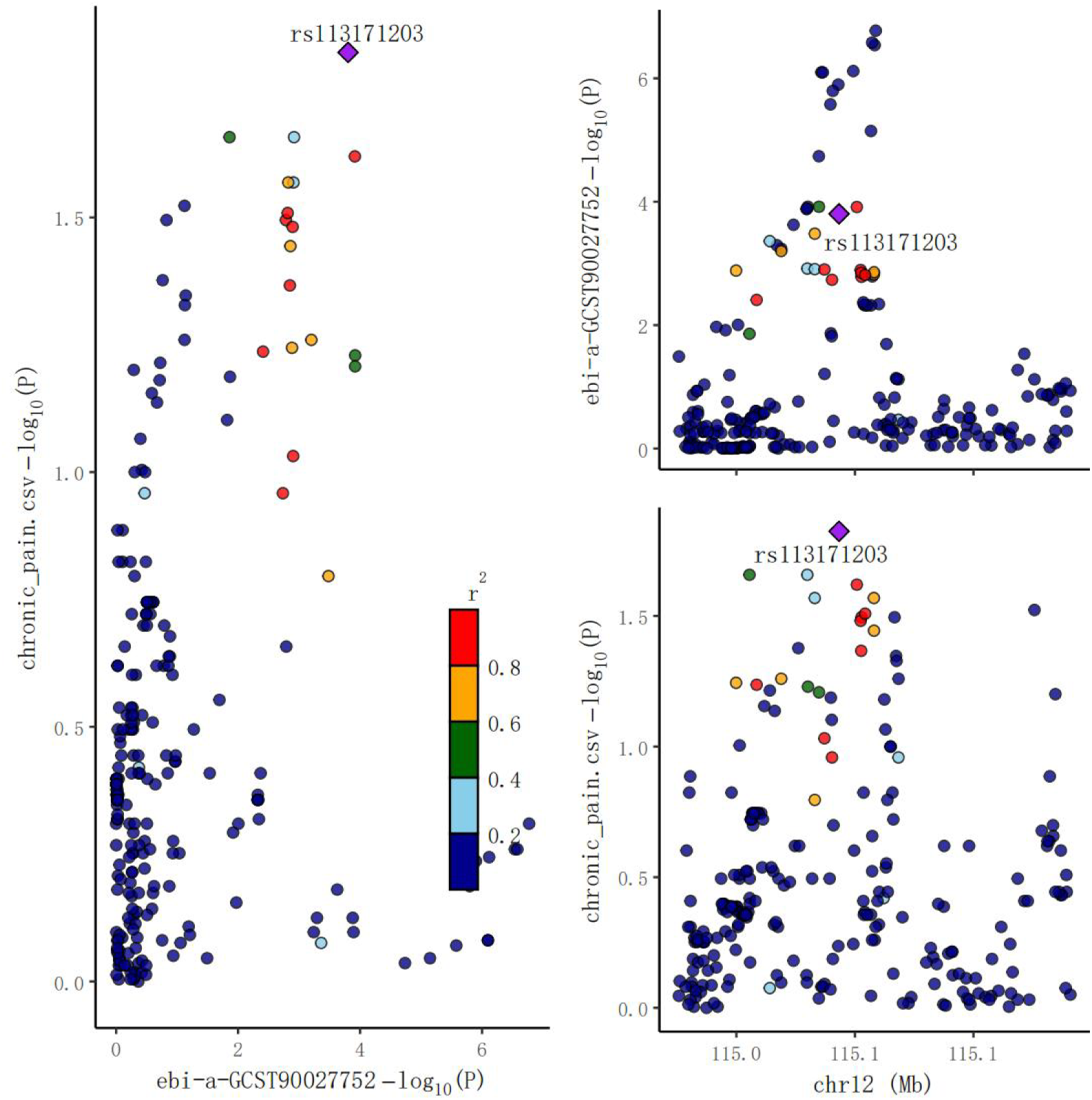
The locuscomparer plot of coloc analysis: Phylum Verrucomicrobia on MCP

**Figure 4c.**
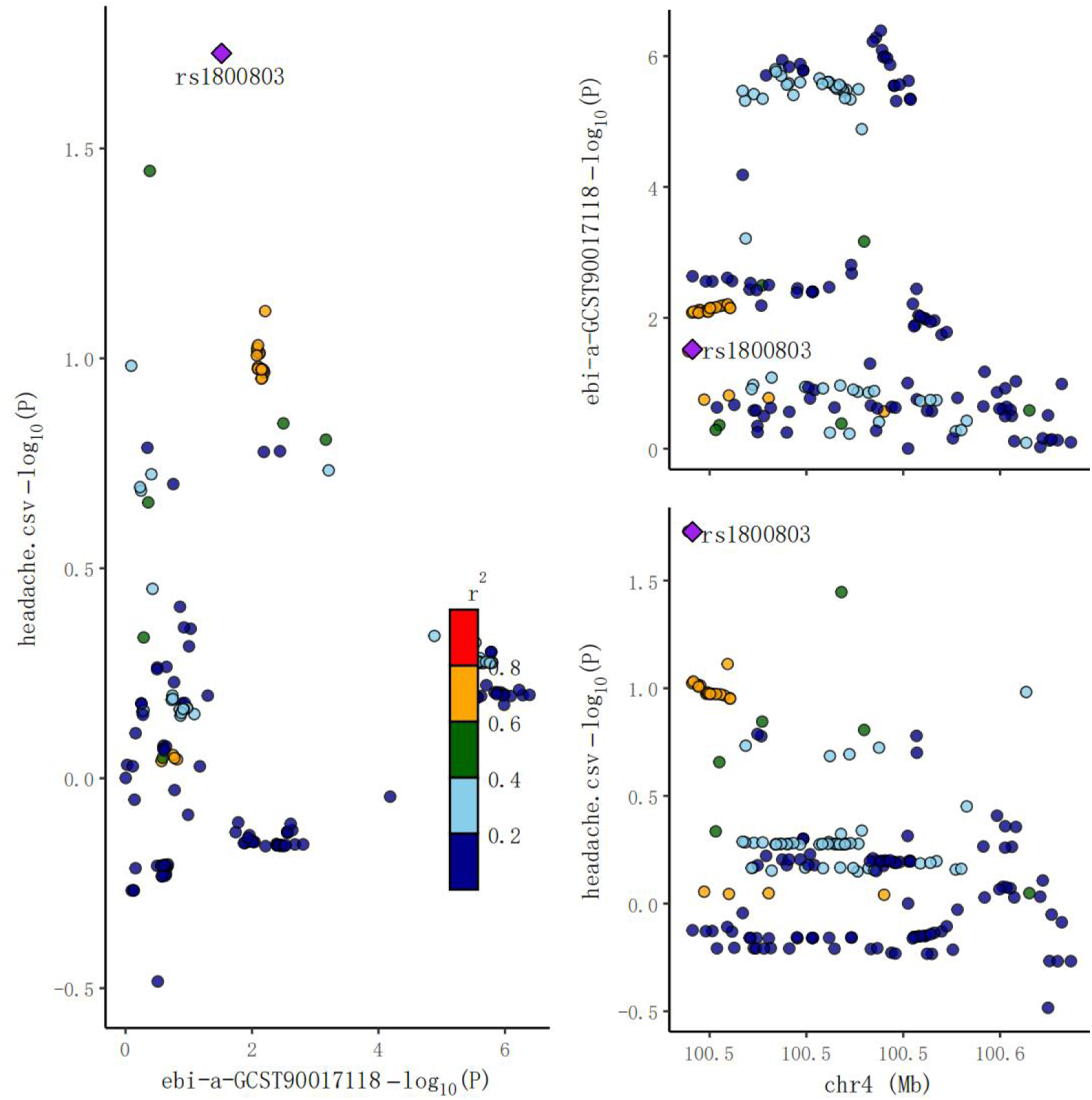
The locuscomparer plot of coloc analysis: Phylum Verrucomicrobia on Headache

**Figure 4d.**
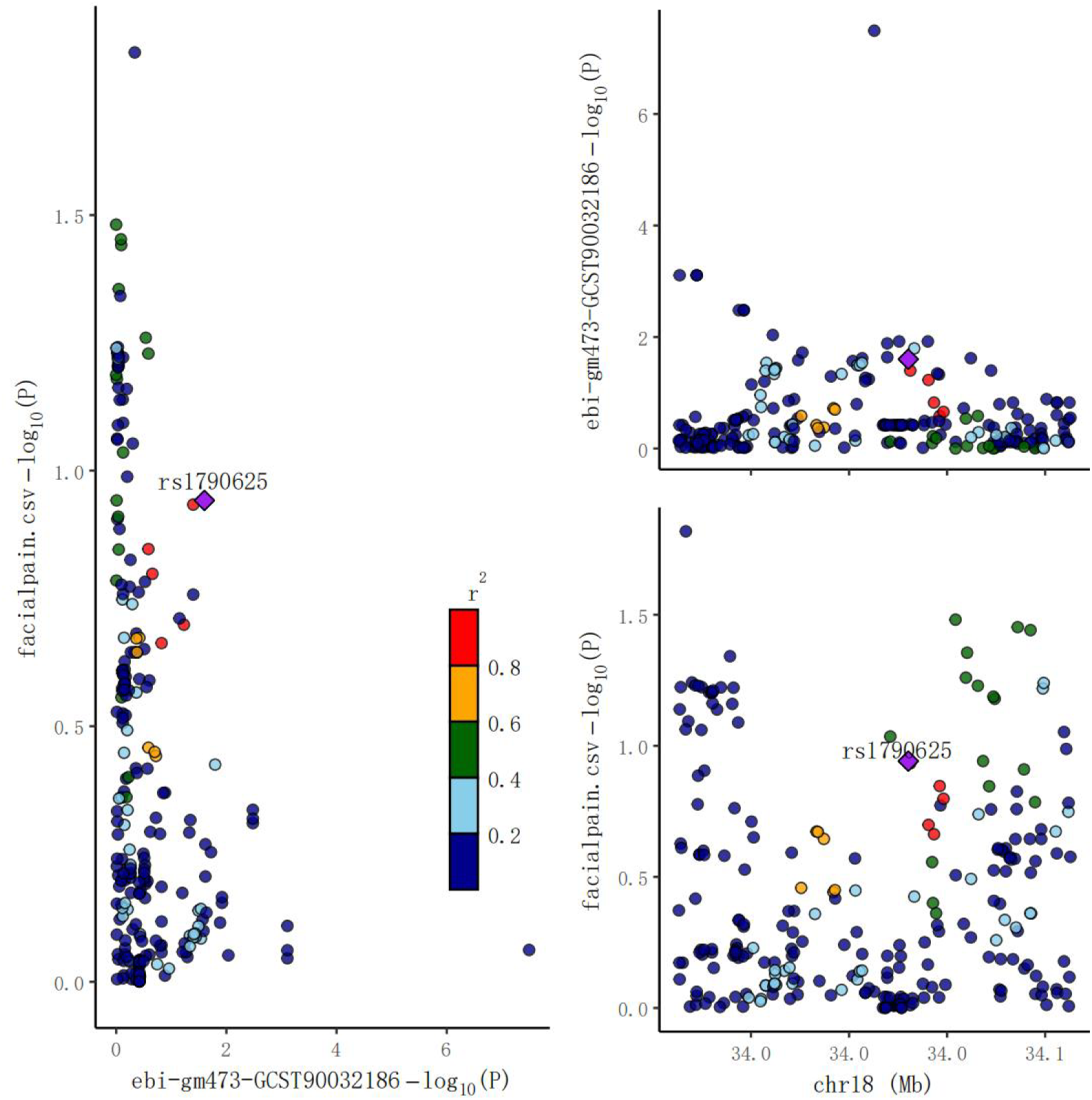
The locuscomparer plot of coloc analysis: Genus Alistipes on Facialpain

**Figure 4e.**
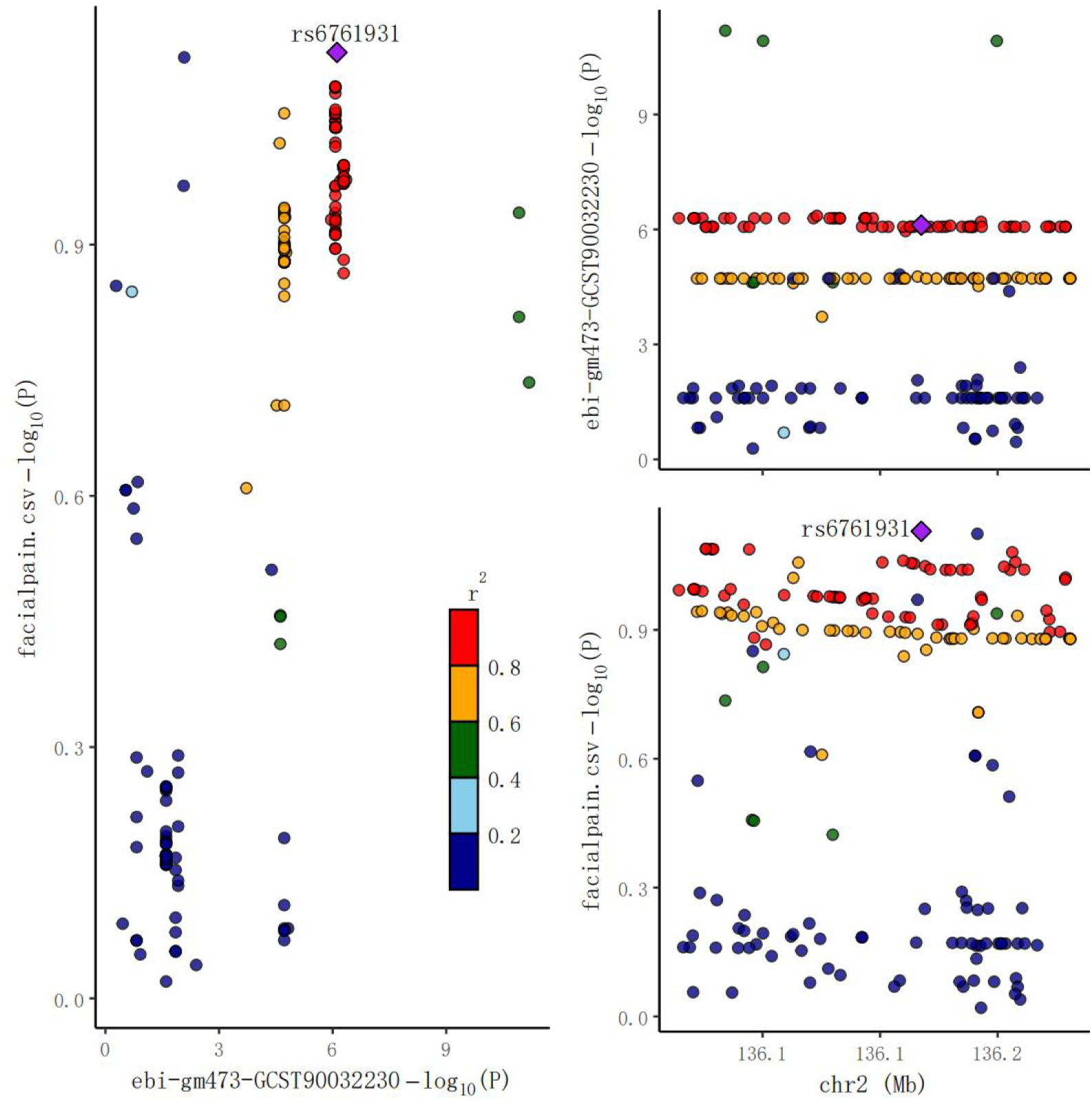
The locuscomparer plot of coloc analysis: Genus Bifidobacterium on Facialpain

**Figure 4f.**
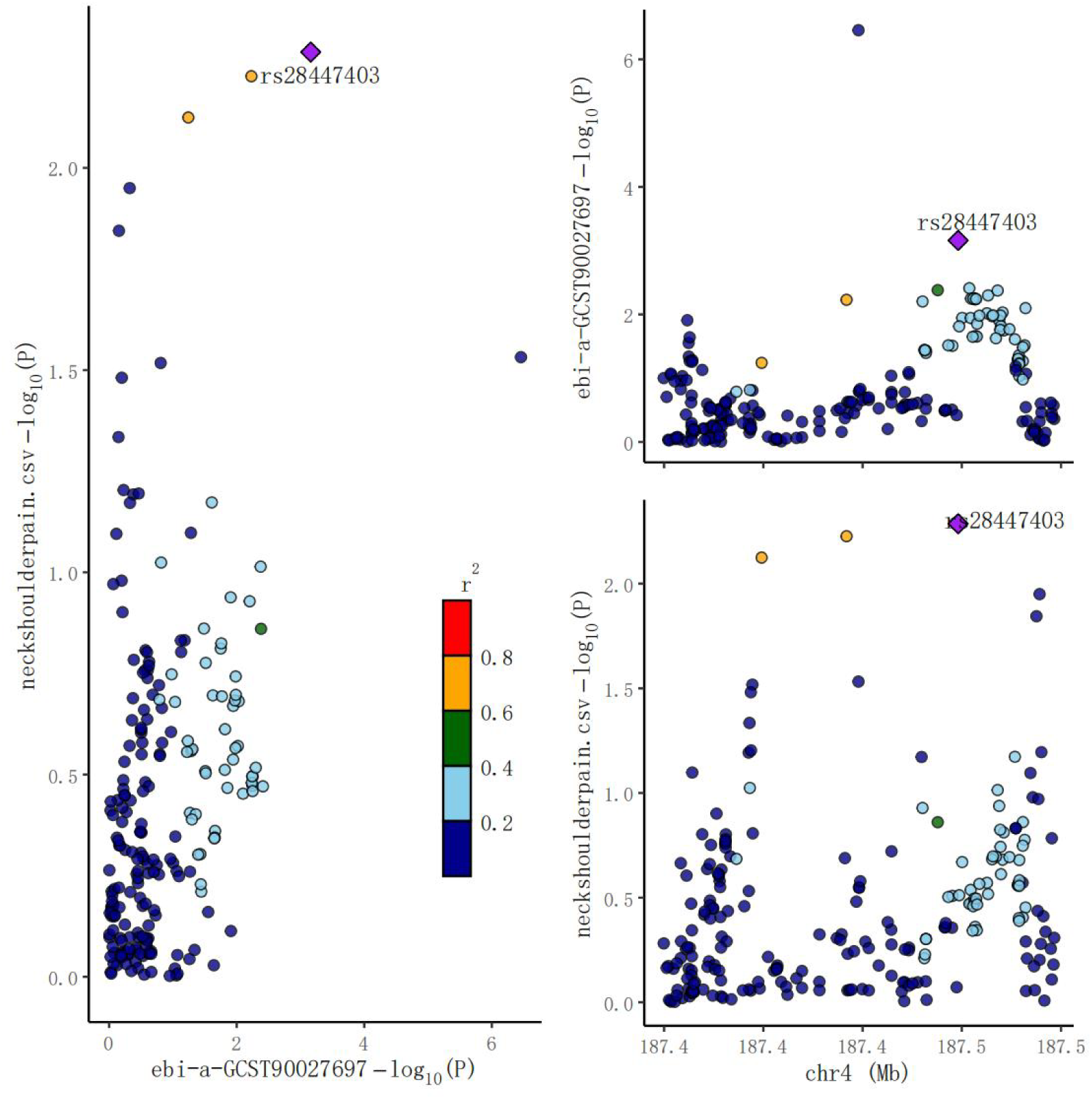
The locuscomparer plot of coloc analysis: Genus Odoribacter on Neckshoulderpain

**Figure 4g.**
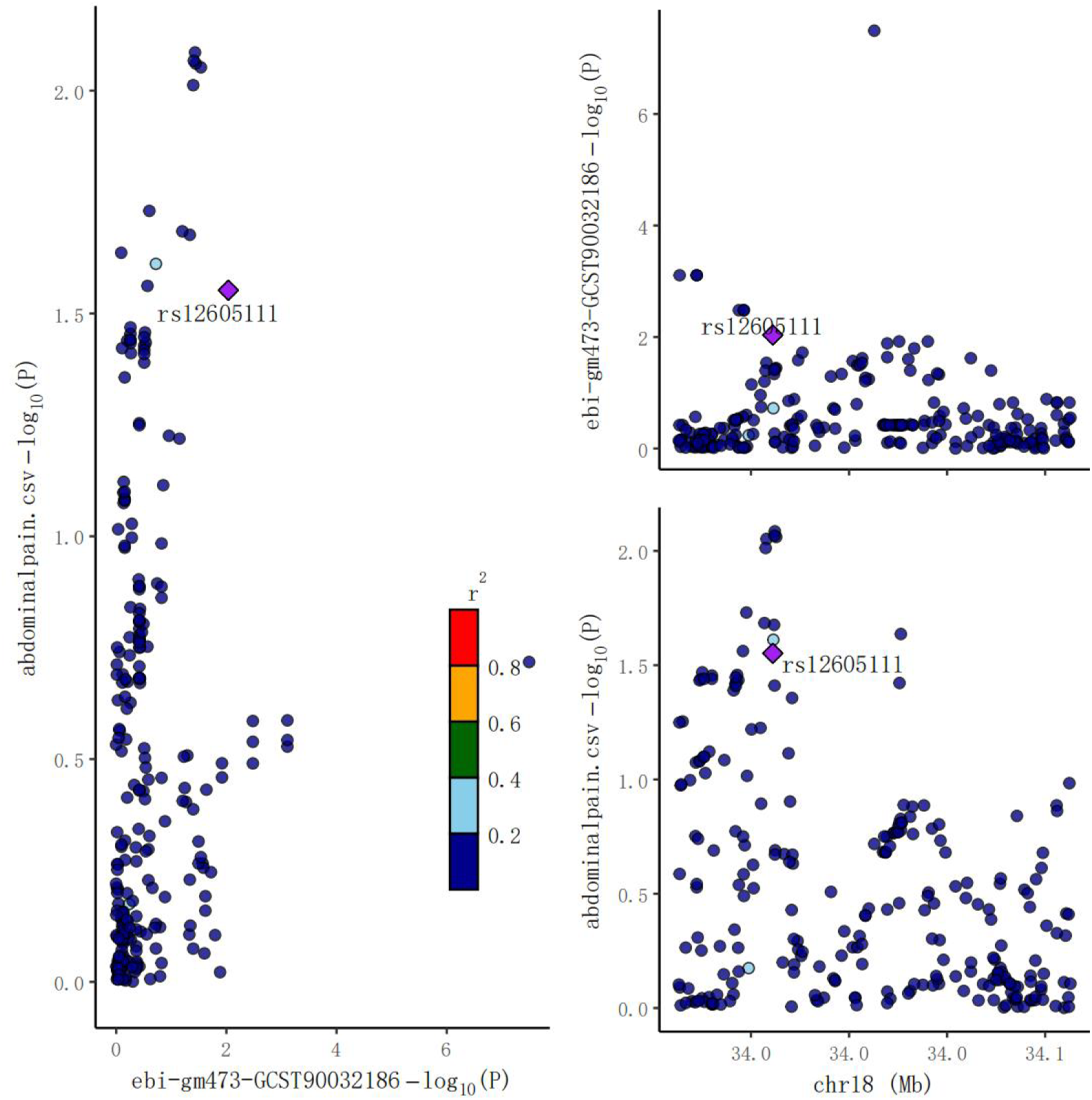
The locuscomparer plot of coloc analysis: Genus Alistipes on Abdominalpain

**Figure 4h.**
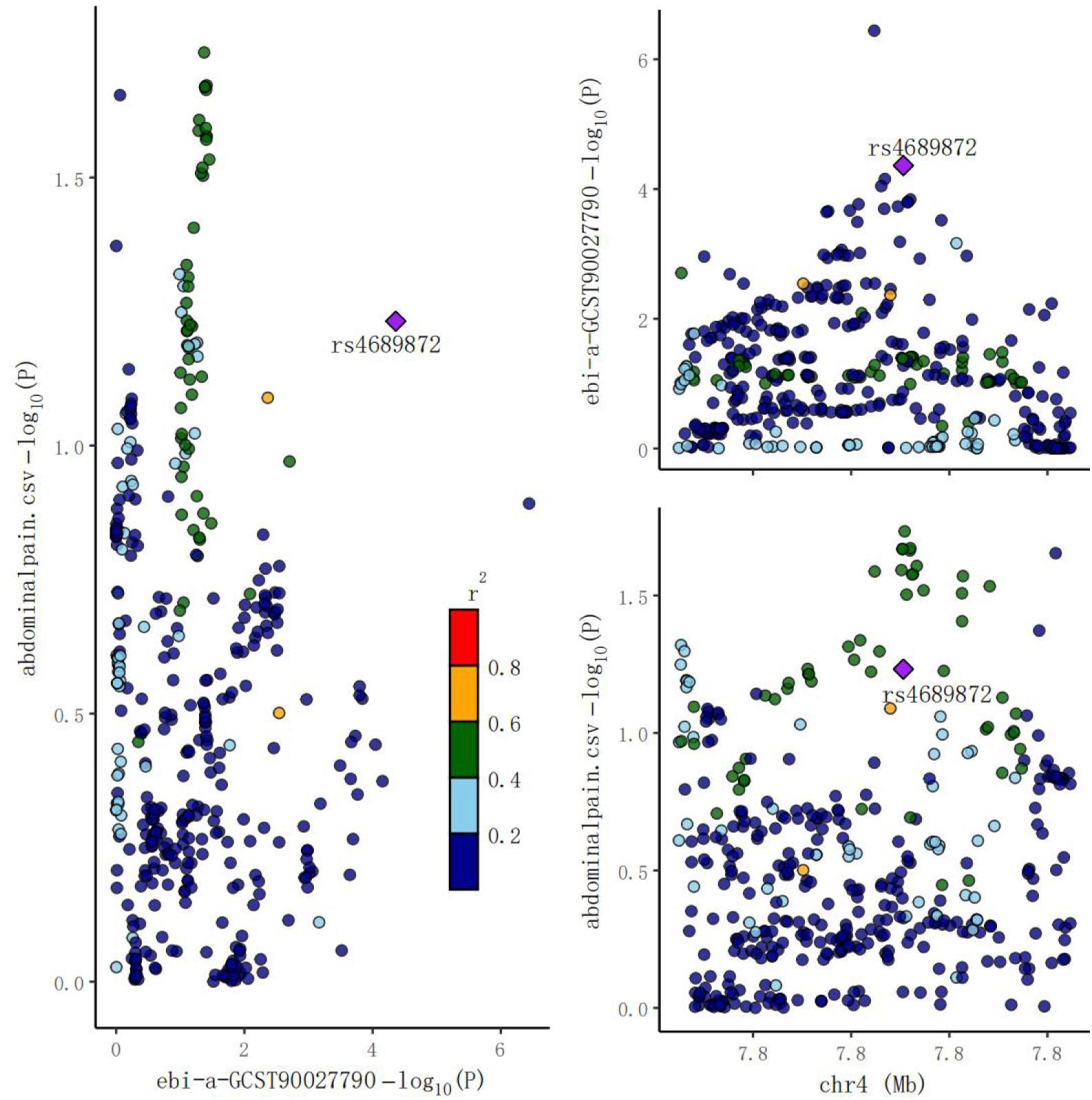
The locuscomparer plot of coloc analysis: Specie Eubacterium_eligens on Abdominalpain

**Figure 4i.**
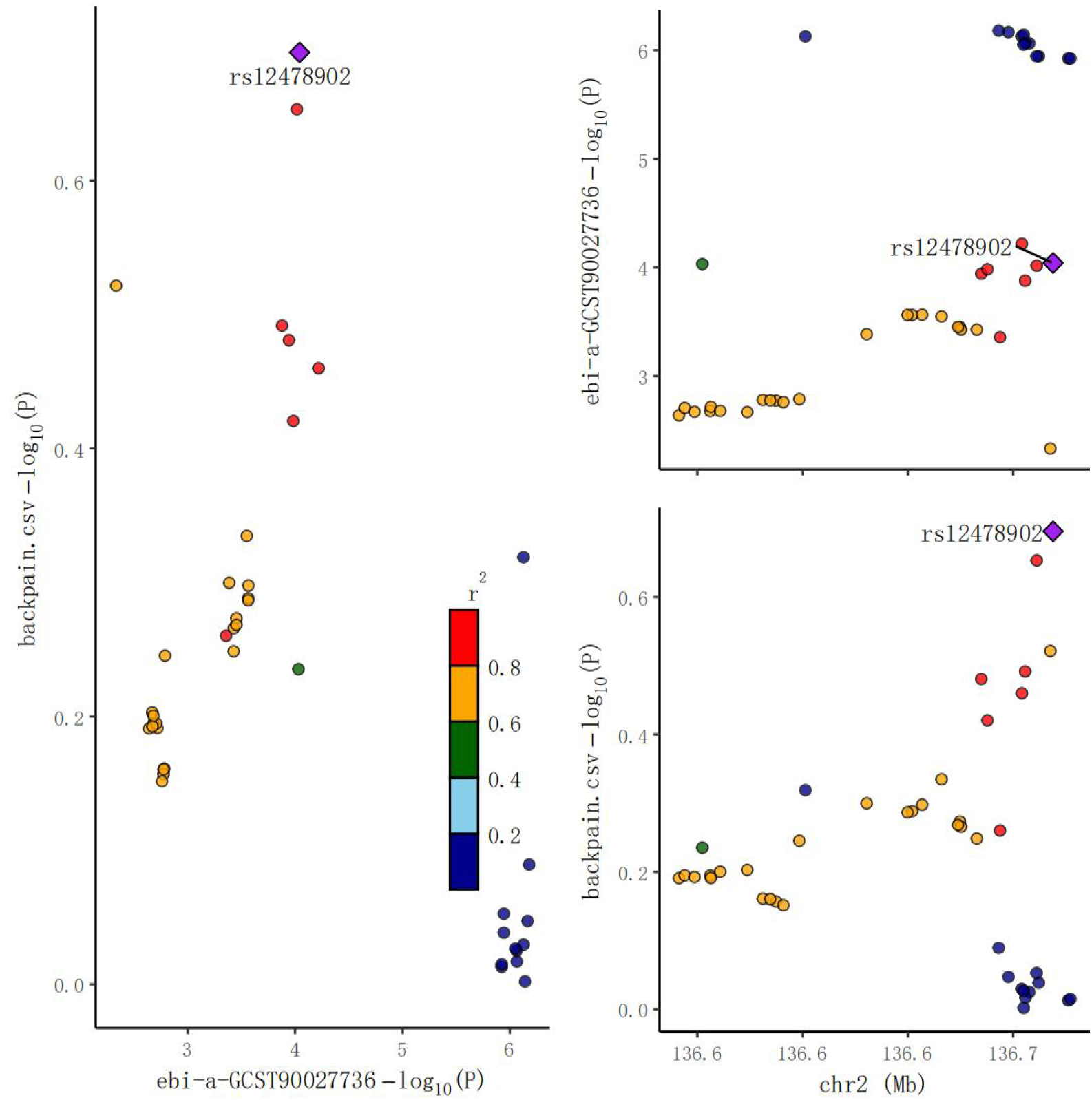
The locuscomparer plot of coloc analysis: Order Bifidobacteriales on Backpain

**Figure 4j.**
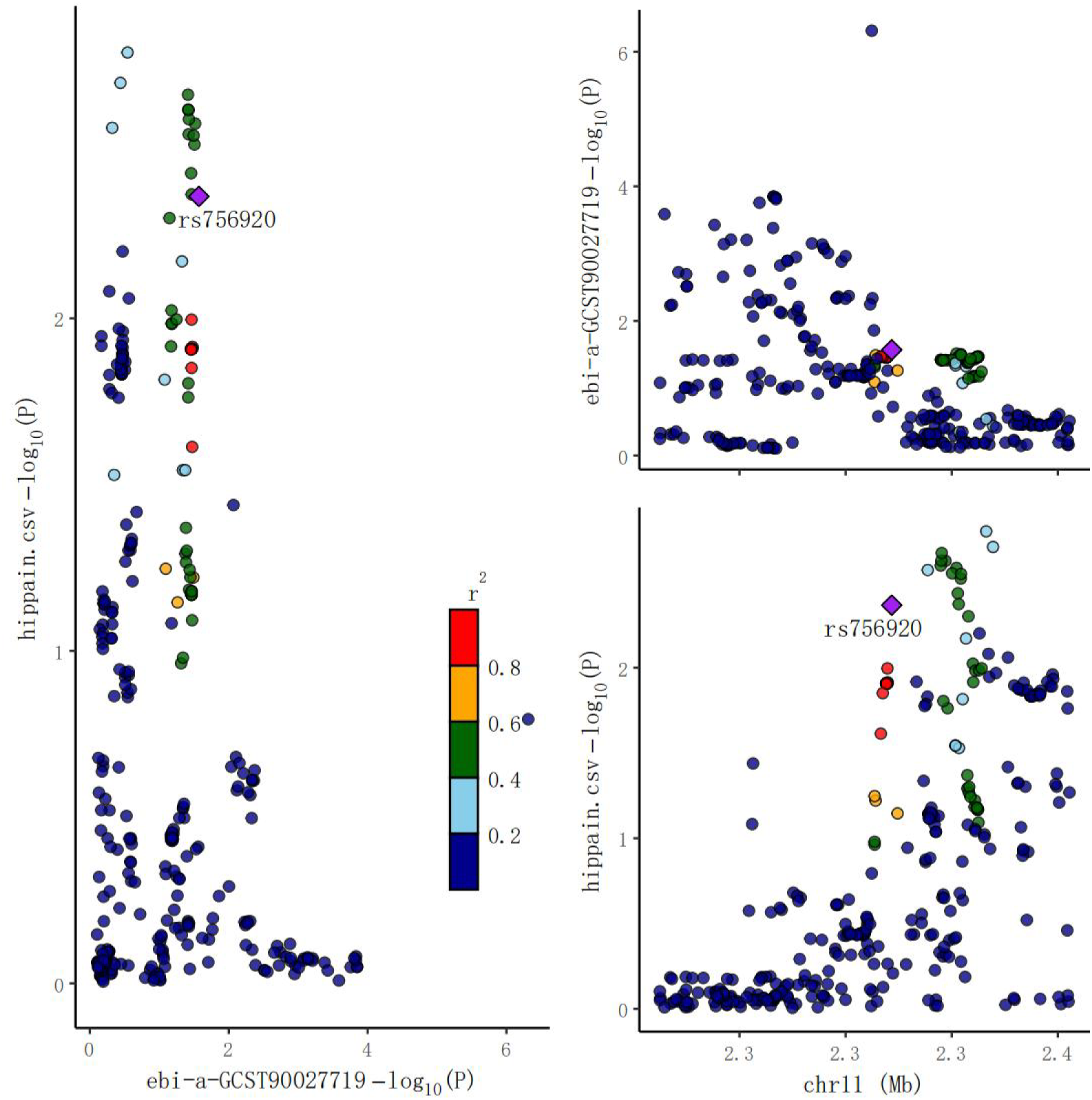
The locuscomparer plot of coloc analysis: Genus Subdoligranulum on Hippain

**Figure 4k.**
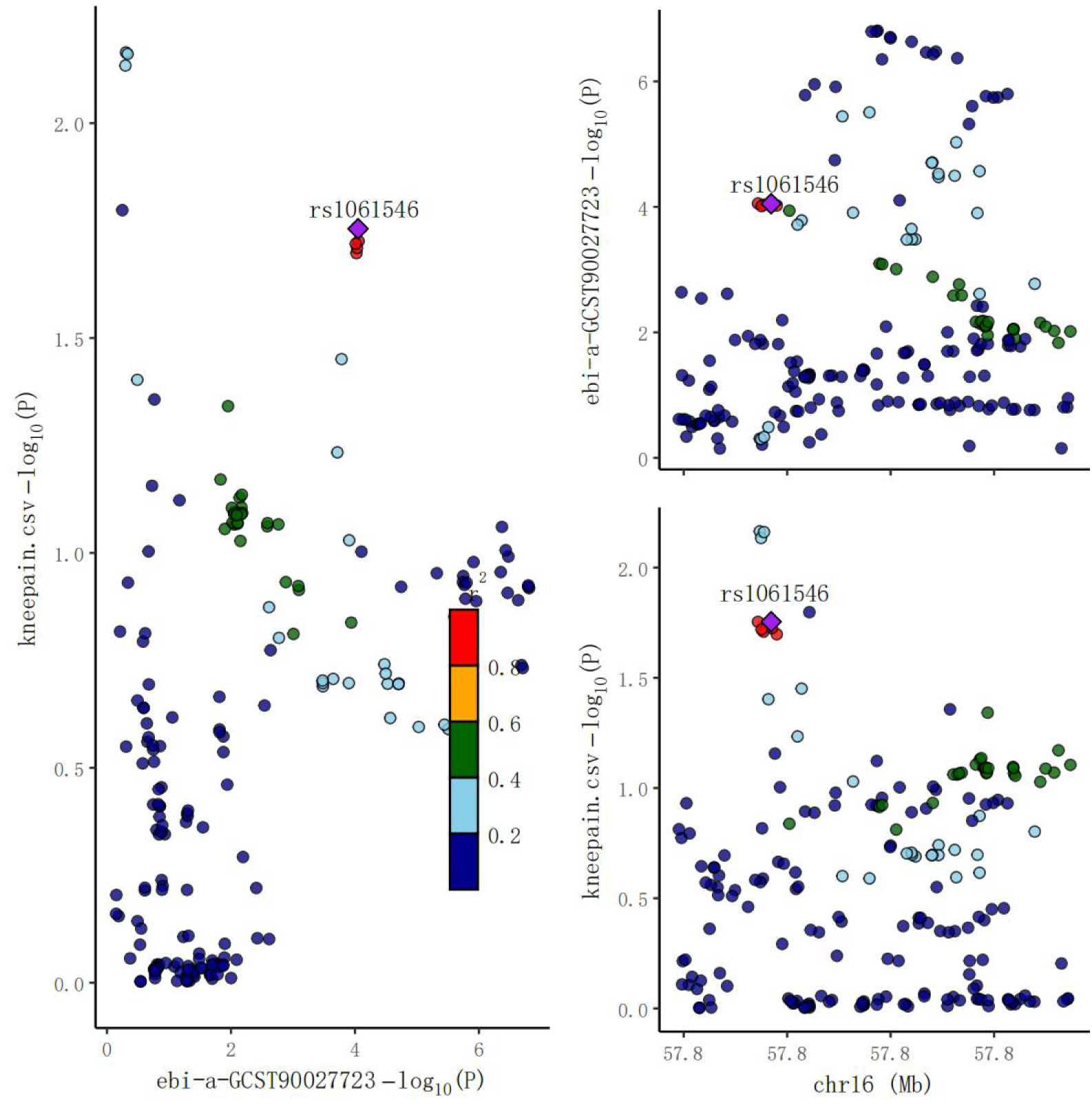
The locuscomparer plot of coloc analysis: Genus Dialister on Kneepain

**Figure 4l.**
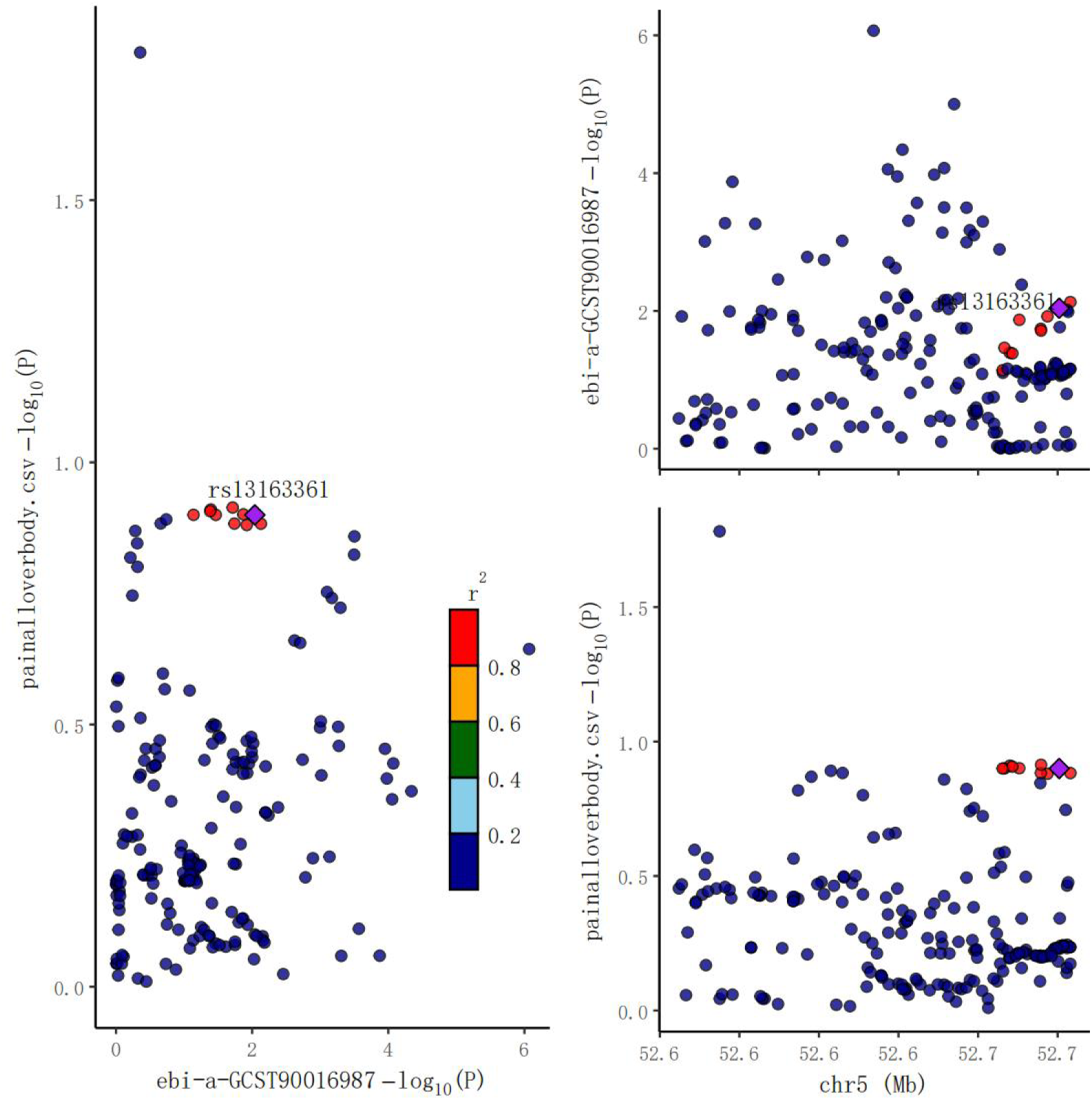
The locuscomparer plot of coloc analysis: Genus Desulfovibrio on pain all over body

**Figure 4m.**
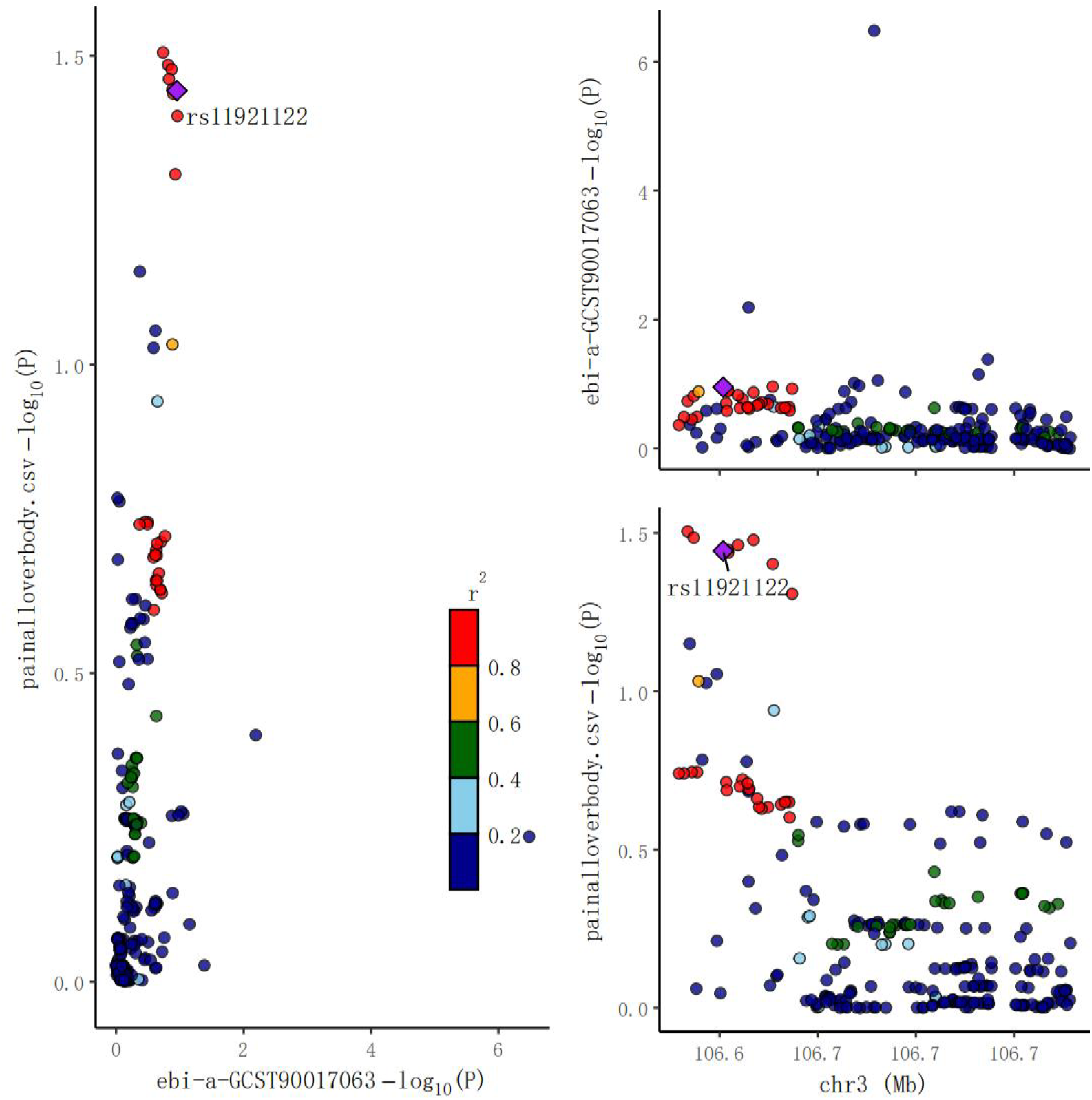
The locuscomparer plot of coloc analysis: Genus Ruminococcus2 on pain all over body

## 4. Statistical analyses

We used R version 4.2.0 for all statistical analyses. We employed various statistical methodologies to evaluate the causal influence of GM on chronic pain phenotypes. Commonly used approaches included the multiplicative random effects–inverse-variance weighted (MRE-IVW) [56], weighted mode (WM) [26], weighted median (WME) [8], and MR-Egger regression [7]. Additionally, we incorporated more recent methods like contamination mixture (ConMix) [14], MR-robust adjusted profile score (MR-RAPS)[96], debiased inverse-variance weighted (dIVW) method [94], and cML-MA [93] to enhance the robustness of our findings. False discovery rate (FDR) correction was applied to strengthen the statistical power of the results. Given the potential bias in IVW estimation when pleiotropic IVs were utilized [13], several sensitivity analyses were conducted to address pleiotropy in the causal estimates, including Cochran’s Q statistic for heterogeneity (P>0.05) [9], MR-Egger regression for horizontal pleiotropy [7]and its intercept term (P>0.05), MR-PRESSO [84] for further evaluation of horizontal pleiotropy, leave-one-out analysis to evaluate the influence of individual SNPs on the observed associations, and power calculations using a non-centrality parameter-based approach [11] and publicly available mRnd web tool (http://cnsgenomics.com/shiny/mRnd/). Any results with evidence of confounding by pleiotropy were excluded (Online Methods). We also limited our analysis to pairs of variables supported by at least three genetic variants.

We used meta-analysis to identify the positive GM, which were then subjected to mediation analysis. To verify whether the screened positive GM and pain phenotype share the same causal variation in a given region (50 kb above and below each SNP), we employed genetic colocalization analysis (coloc). Furthermore, Bayesian weighted MR (BWMR) was implemented to strengthen the evidence for a causal relationship between the identified positive GM and pain phenotypes [95]. We also examined the impact of these potential positive GM on specific brain regions within the cerebral cortex.

To assess the potential mediating role of the intermediary effect (IE) on the relationship between the exposure (a) and the outcome (c’), we conducted a mediation analysis using an interactive mediation framework and Sobel tests (http://quantpsy.org/sobel/sobel.htm) (Figure 1(b-2). The IVW method was employed to estimate the causal effects, including IE (a*b), direct effect (DE; c’ - (a*b)), and total effect (TE; c’). Statistical significance was determined by a two-sided P-value threshold of 0.05.

We employed FDR correction to account for multiple comparisons when evaluating statistical significance. Associations with an FDR of <0.05 were considered statistically significant. However, we expanded the p-value of the meta-analysis to 0.1 when no p-value was less than 0.05. We acknowledge the exploratory nature of this study and report both raw P and FDR values.

## 5 Results

### 5.1 Preliminary results of MR: GM on MCP and eight pain phenotypes

#### 5.1.1 GM on MCP

We observed that 4 taxa (2 orders and 2 families) were related to MCP in Data 1, 8 taxa (2 genera, 2 OTU97, 2 OTU99, and 2 TestASV) were related to MCP in Data 2, 11 taxa (1 phylum, 1 class, 1 order, 2 families, 2 genera, and 4 species) were related to MCP in Data 3, and 18 taxa (1 class, 2 orders, 1 family, 7 genera, and 7 unclassified) were related to MCP in Data 4. (see Supplementary 1)

#### 5.1.2 GM on Headache

We observed that 7 taxa (1 phylum, 1 class, 1 order, 2 families, and 2 genera) were related to headache in Data 1, 9 taxa (1 class, 1 order, 1 family, 1 OTU97, 1 OTU99, and 4 TestASV) were related to headache in Data 2, 7 taxa (1 order, 1 family, 2 genera, and 3 species) were related to headache in Data 3, and 17 taxa (1 order, 2 families, 6 genera, 2 species, and 6 unclassified) were related to headache in Data 4.(see Supplementary 2)

#### 5.1.3 GM on Facial Pain

We observed that 6 taxa (1 order, 2 families, and 3 genera) were related to facial pain in Data 1, 9 taxa (1 genus, 4 OTU97, 3 OTU99, and 1 TestASV) were related to facial pain in Data 2, 5 taxa (1 family, 1 genus, and 3 species) were related to facial pain in Data 3, and 22 taxa (2 phyla, 1 order, 2 families, 9 genera, 2 species, and 7 unclassified) were related to facial pain in Data 4. (see Supplementary 3)

#### 5.1.4 GM on Neck/Shoulder Pain

We observed that 4 taxa (1 phylum, 2 families, and 1 genus) were related to neck/shoulder pain in Data 1, 5 taxa (1 class, 1 family, 1 OTU97, 1 OTU99, and 1 TestASV) were related to neck/shoulder pain in Data 2, 5 taxa (1 class, 1 order, 1 family, 1 genus, and 1 species) were related to neck/shoulder pain in Data 3, and 20 taxa (1 family, 10 genera, 2 species, and 7 unclassified) were related to neck/shoulder pain in Data 4.(see Supplementary 4)

#### 5.1.5 GM on Abdominal Pain

We observed that 8 taxa (1 class, 2 orders, 2 families, and 3 genera) were related to abdominal pain in Data 1, 8 taxa (1 class, 1 genus, 3 OTU97, 2 OTU99, and 1 TestASV) were related to abdominal pain in Data 2, 3 taxa (3 species) were related to abdominal pain in Data 3, and 19 taxa (2 orders, 1 family, 9 genera, 3 species, and 4 unclassified) were related to abdominal pain in Data 4. (see Supplementary 5)

#### 5.1.6 GM on Back Pain

We observed that 5 taxa (1 phylum, 1 class, 1 order, and 2 genera) were related to back pain in Data 1, 6 taxa (1 family, 1 genus, 2 OTU97, and 2 OTU99) were related to back pain in Data 2, 3 taxa (1 order, 1 family, and 1 species) were related to back pain in Data 3, and 16 taxa (2 classes, 2 families, 7 genera, 1 species, and 4 unclassified) were related to back pain in Data 4. (see Supplementary 6)

#### 5.1.7 GM on Hip Pain

We observed that 7 taxa (1 family and 6 genera) were related to hip pain in Data 1, 8 taxa (3 OTU97, 3 OTU99, and 2 TestASV) were related to hip pain in Data 2, 8 taxa (1 family, 4 genera, and 3 species) were related to hip pain in Data 3, and 17 taxa (2 orders, 2 families, 8 genera, and 5 unclassified) were related to hip pain in Data 4. (see Supplementary 7)

#### 5.1.8 GM on Knee Pain

We observed that 11 taxa (2 phyla, 2 classes, 3 orders, and 4 genera) were related to knee pain in Data 1, 3 taxa (3 OTU99) were related to knee pain in Data 2, 5 taxa (1 family and 4 genera) were related to knee pain in Data 3, and 21 taxa (2 orders, 5 families, 9 genera, and 5 unclassified) were related to knee pain in Data 4. (see Supplementary 8)

#### 5.1.9 GM on Pain All Over the Body

We observed that 5 taxa (5 genera) were related to pain all over the body in Data 1, 11 taxa (1 class, 1 order, 1 family, 2 OTU97, 5 OTU99, and 1 TestASV) were related to pain all over the body in Data 2, 10 taxa (1 class, 2 orders, 3 families, 2 genera, and 2 species) were related to pain all over the body in Data 3, and 21 taxa (11 genera, 3 species, and 7 unclassified) were related to pain all over the body in Data 4. (see Supplementary 9) After adjusting the p-values by FDR, Genus *Odoribacter* was still significantly associated with neck/shoulder pain (P_IVW_ = 1.203×10^-4, P_FDR_ = 0.048).

### 5.2 Results of meta-analysis: GM on MCP and eight pain phenotypes

In the second step, we conducted a meta-analysis based on the preliminary results from Section 5.1 in this exploratory study (see Table 2, Figure 2 and 3).

**Table 2:**
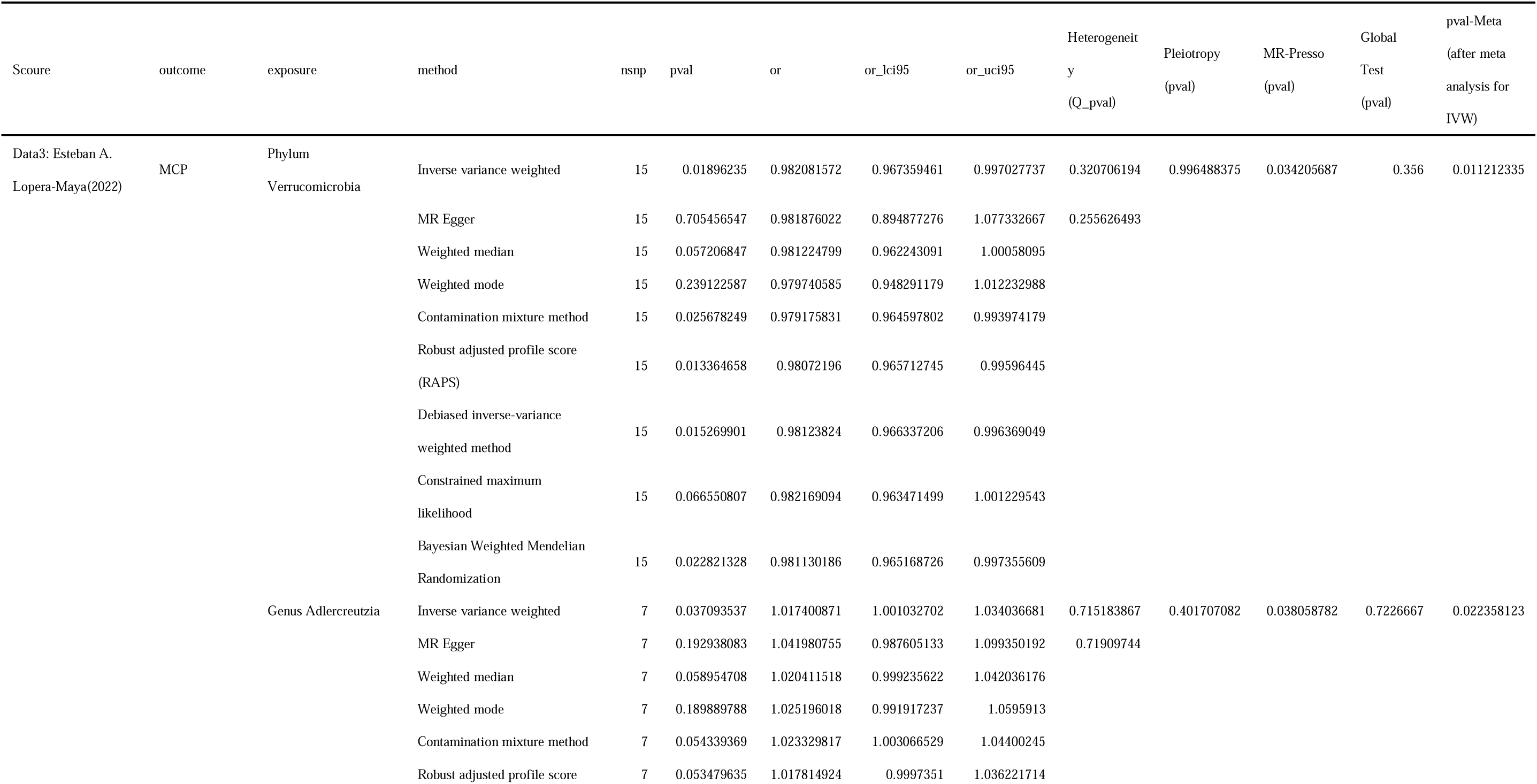
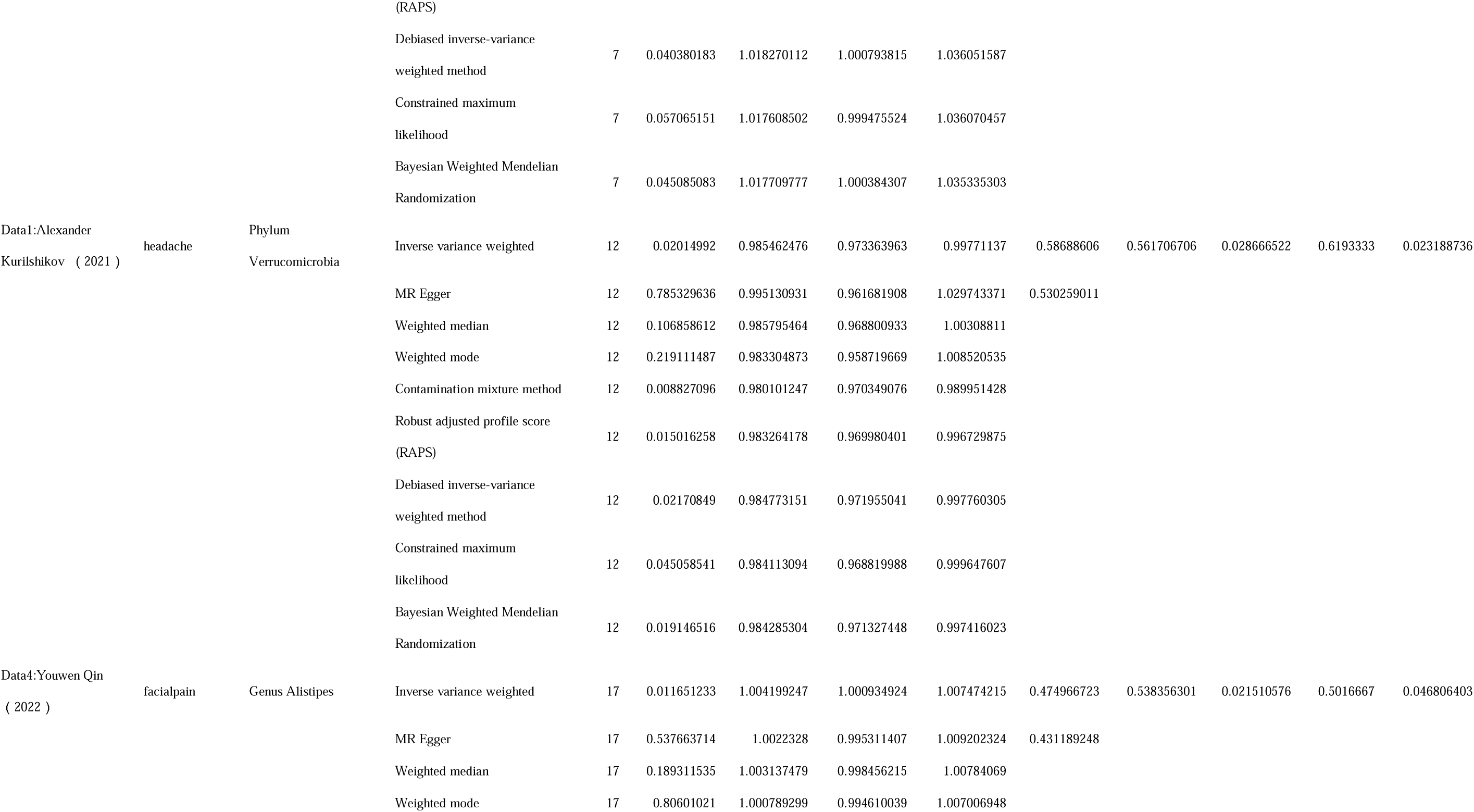
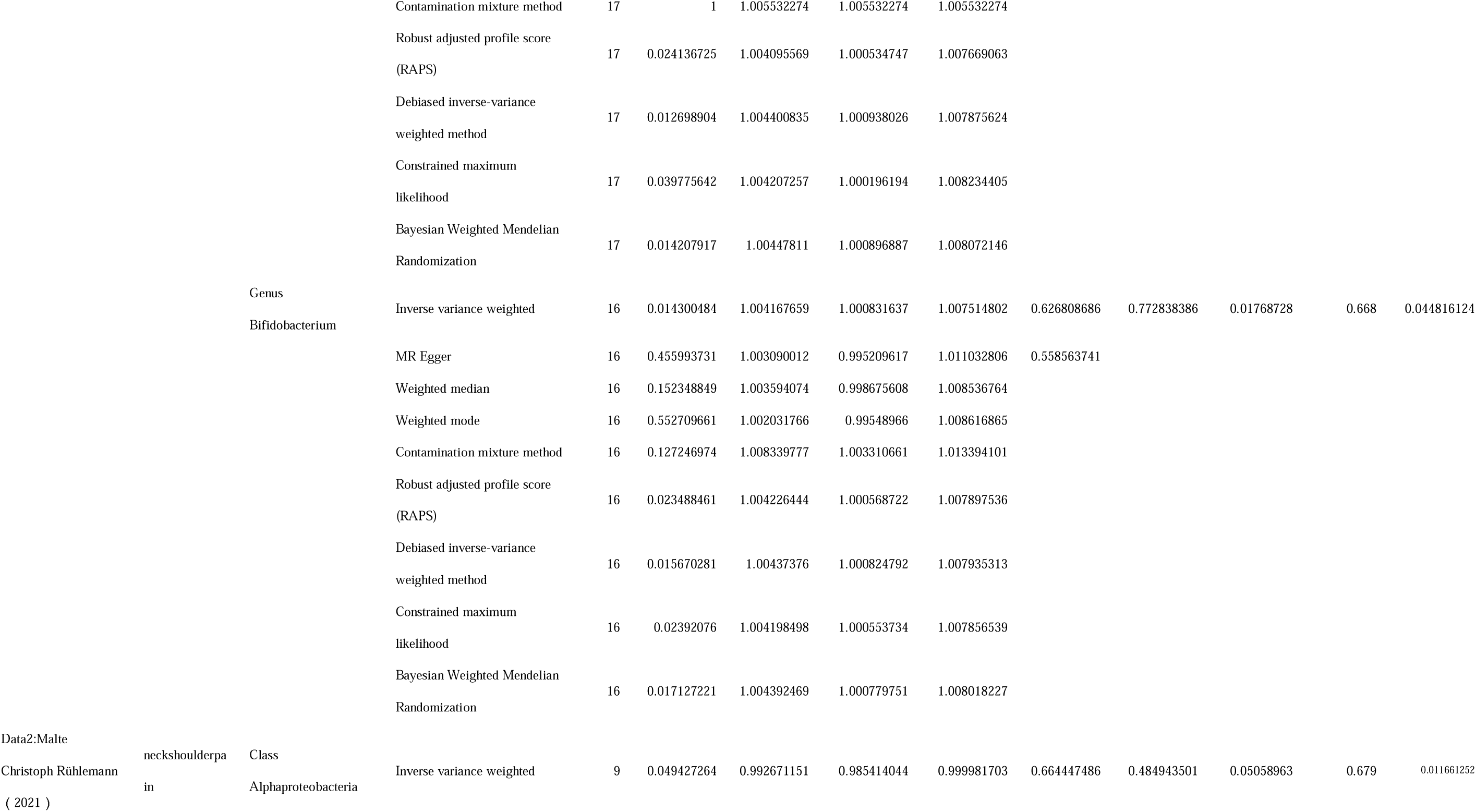
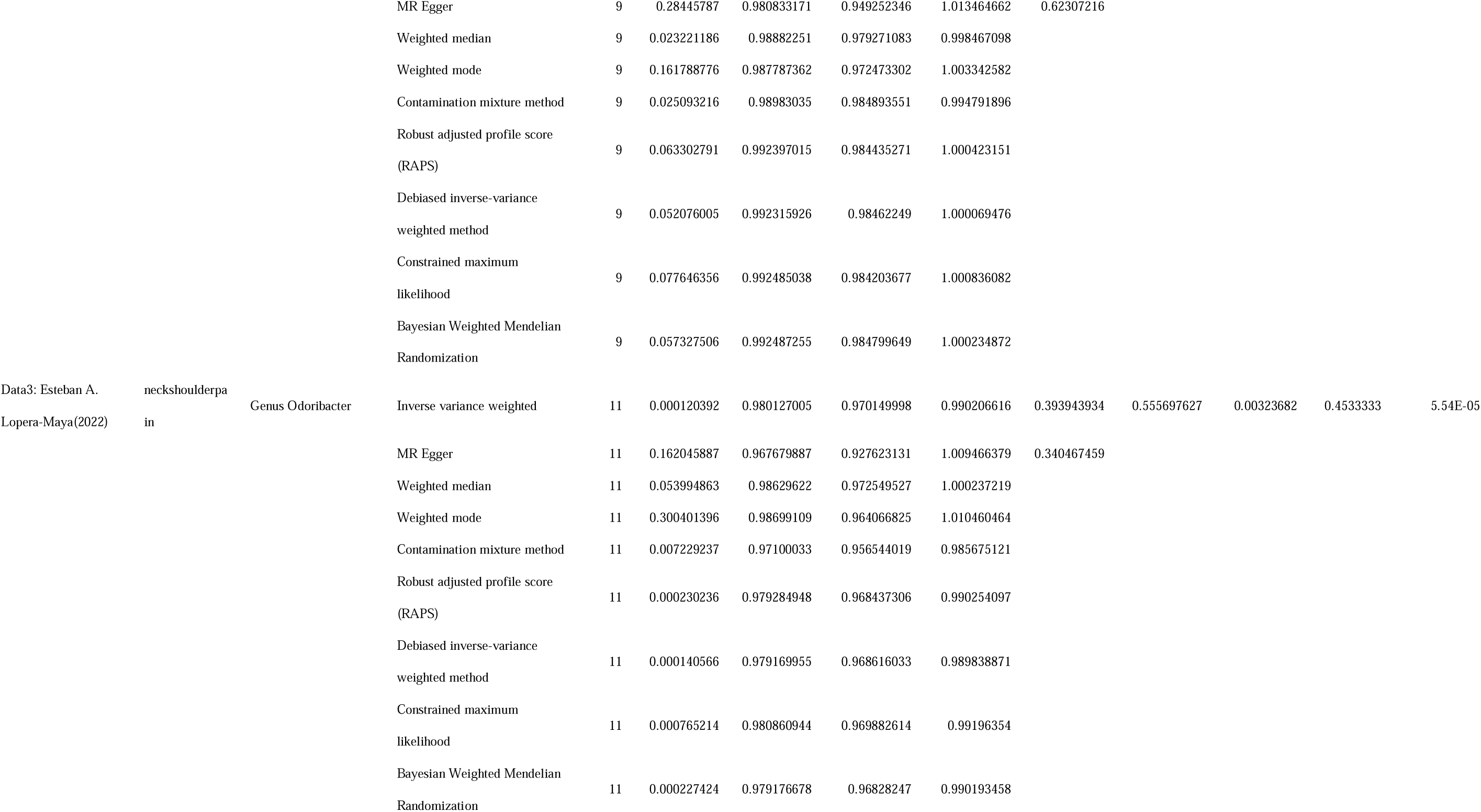
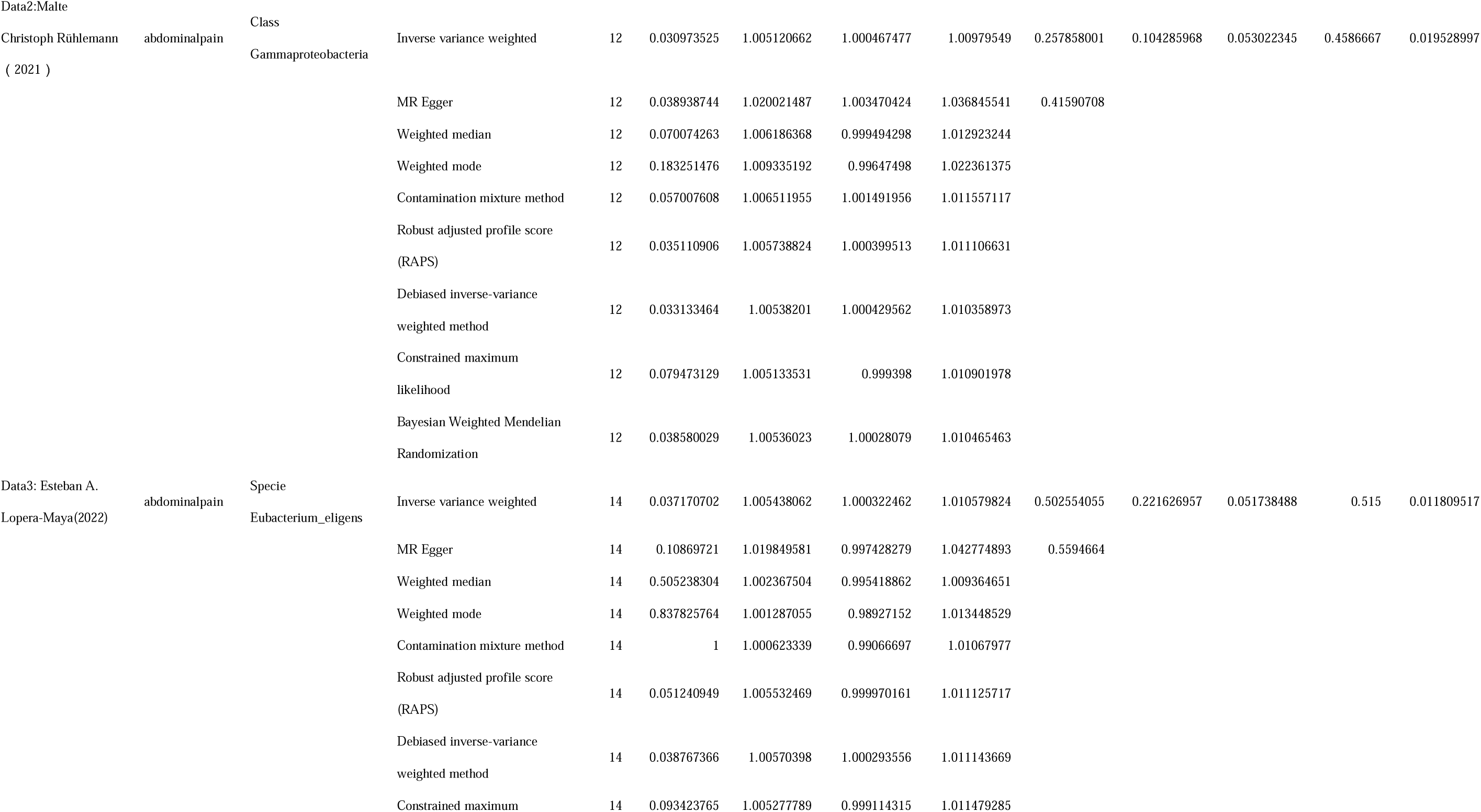
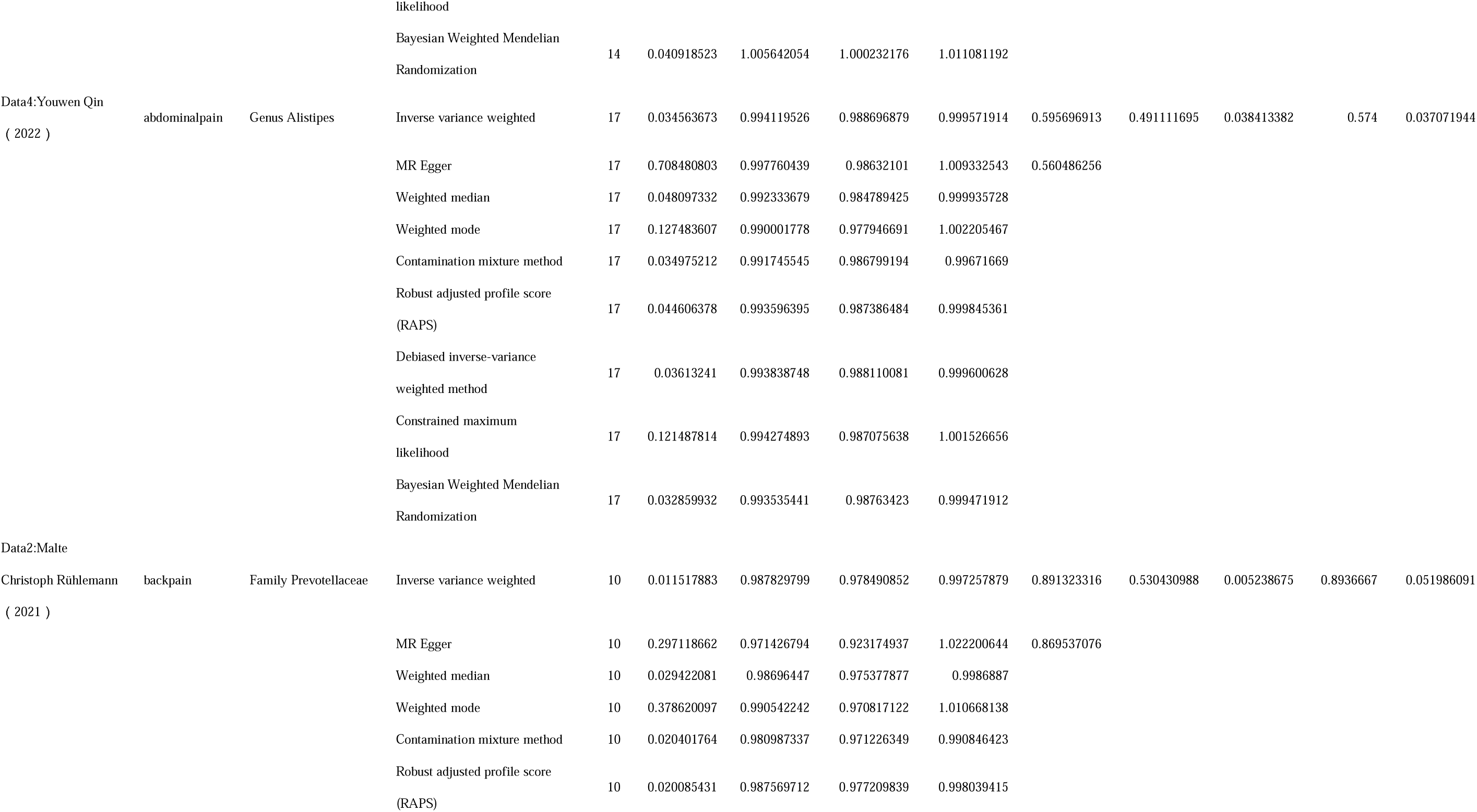
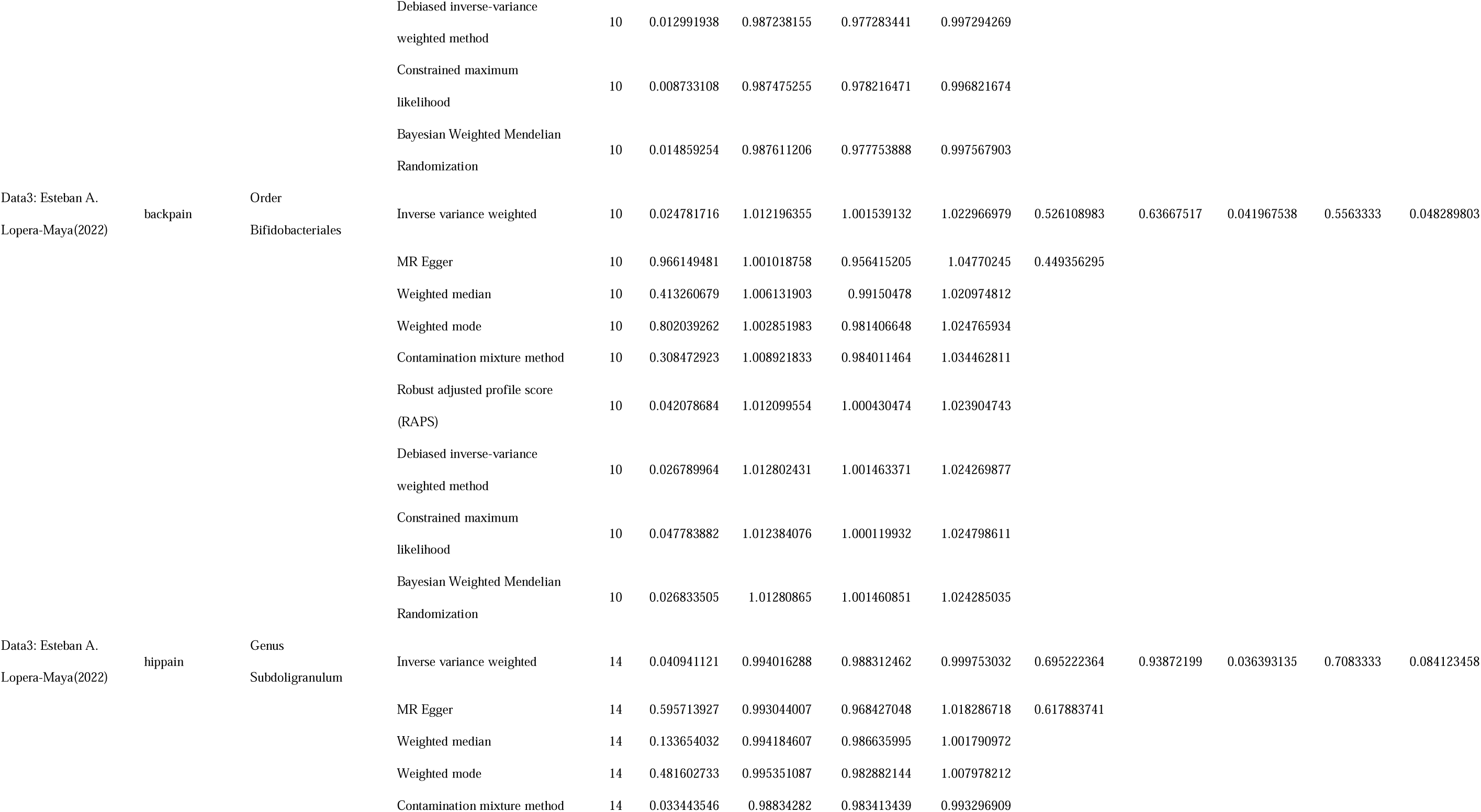
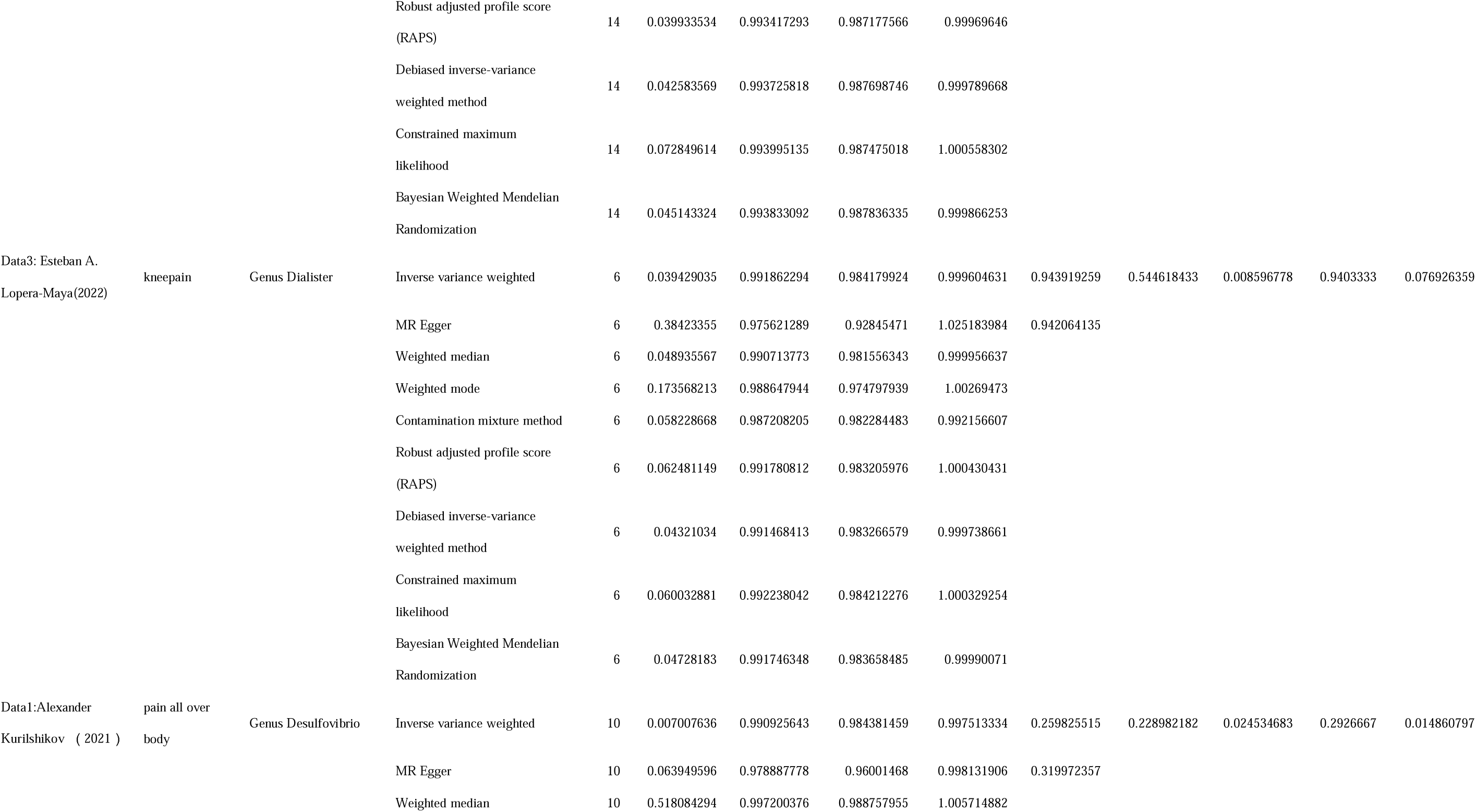
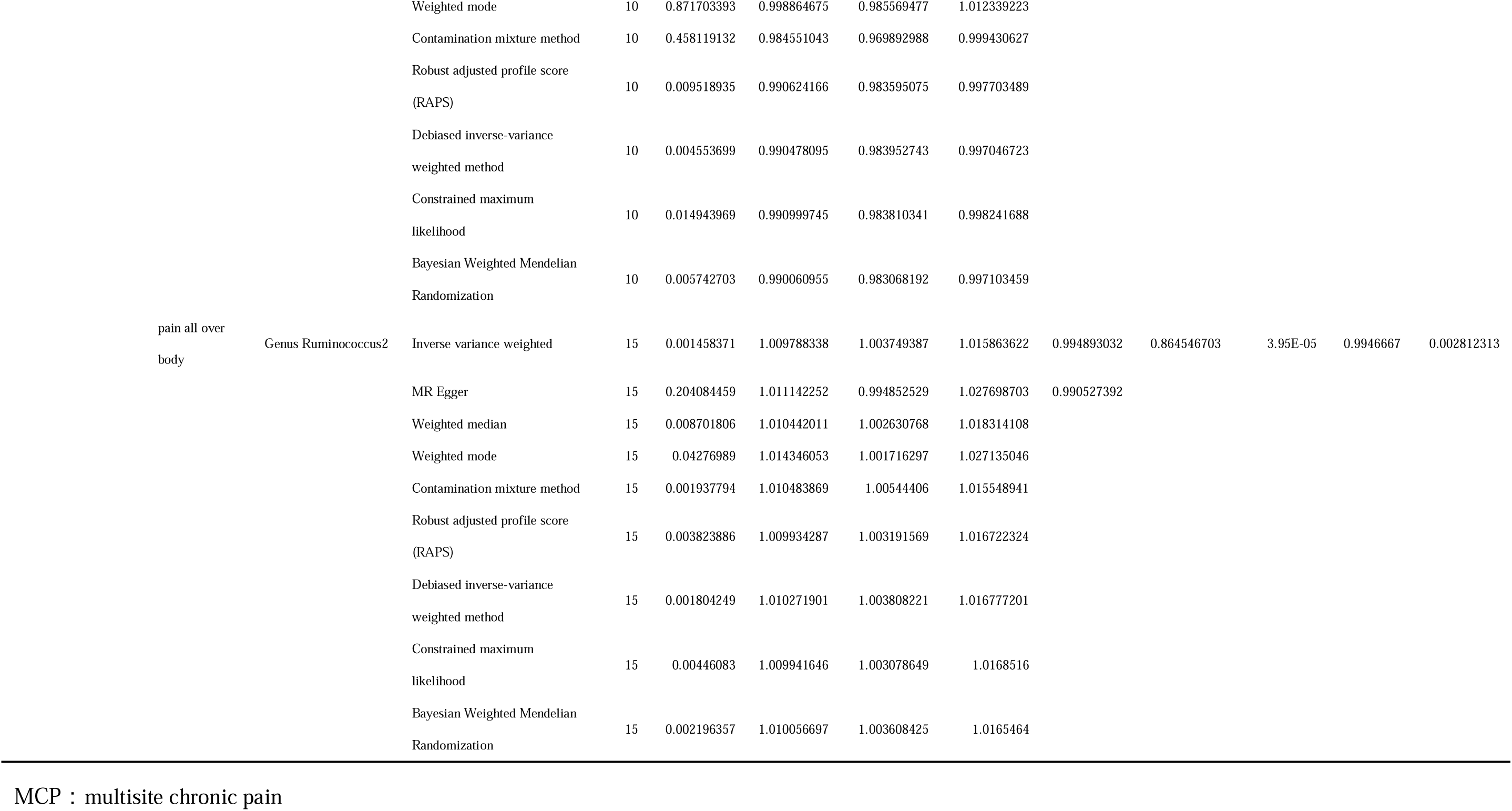
Mendelian randomization results of positive gut microbiota abundance based on Meta analysis on MCP and 8 pain phenotypes.

**Table 3:** genetic colocalisation analysis results of positive gut microbiota abundance based on Meta analysis on MCP and 8 pain phenotypes.

| Scoure | outcome | exposure | nsn<br>p | PP.H4.abf |
| --- | --- | --- | --- | --- |
| Data3: Esteban A.<br>Lopera-Maya(2022) | MCP | Phylum<br>Verrucomicrobia | 258 | 0.0018262<br>13 |
|  |  | Genus Adlercreutzia | 130 | 0.0016919<br>34 |
| Data1:Alexander Kurilshikov<br>( 2021 ) | headache | Phylum<br>Verrucomicrobia | 178 | / |
| Data4:Youwen Qin ( 2022 ) | facialpain | Genus Alistipes | 249 | 0.0422121<br>55 |
|  |  | Genus Bifidobacterium | 188 | 0.0555433<br>83 |
| Data3: Esteban A.<br>Lopera-Maya(2022) | neckshoulderpain | Genus Odoribacter | 225 | 0.0520624<br>98 |
| Data2:Malte Christoph Rühlemann<br>( 2021 ) | abdominalpain | Class<br>Gammaproteobacteria |  |  |
| Data3: Esteban A.<br>Lopera-Maya(2022) | abdominalpain | Specie<br>Eubacterium_eligens | 415 | 0.0225092<br>93 |
| Data4:Youwen Qin ( 2022 ) | abdominalpain | Genus Alistipes | 249 | 0.0558042<br>56 |
| Data2:Malte Christoph Rühlemann<br>( 2021 ) | backpain | Family Prevotellaceae |  |  |
| Data3: Esteban A.<br>Lopera-Maya(2022) | backpain | Order Bifidobacteriales | 42 | 0.0060220<br>88 |
| Data3: Esteban A.<br>Lopera-Maya(2022) | hippain | Genus Subdoligranulum | 333 | 0.0144852<br>39 |
| Data3: Esteban A.<br>Lopera-Maya(2022) | kneepain | Genus Dialister | 205 | 0.0173670<br>48 |
| Data1:Alexander Kurilshikov<br>( 2021 ) | pain all over<br>body | Genus Desulfovibrio | 210 | 0.0128547<br>44 |
|  |  | Genus Ruminococcus2 | 242 | 0.0188889<br>71 |

MCP : multisite chronic pain

**MCP**: The abundance of Genus *Adlercreutzia* (P_IVW_ = 3.71E-02, PMeta = 2.24E-02) was positively associated with MCP, while that of Phylum *Verrucomicrobia* (P_IVW_ = 1.90E-02, P_Meta_ = 1.12E-02) was negatively associated with MCP.

**Headache**: The abundance of Phylum *Verrucomicrobia* (P_IVW_ = 2.02E-02, P_Meta_ = 2.32E-02) was negatively associated with headache.

**Facial Pain**: The abundances of Genus *Alistipes* (P_IVW_ = 1.17E-02, P_Meta_ = 4.68E-02) and Genus *Bifidobacterium* (P_IVW_ = 1.43E-02, P_Meta_ = 4.48E-02) were positively associated with facial pain.

**Neck/Shoulder Pain**: The abundances of Class *Alphaproteobacteria* (P_IVW_ = 4.92E-02, P_Meta_ = 1.17E-02) and Genus *Odoribacter* (P_IVW_ = 1.20E-04, P_Meta_ = 5.54E-05) were negatively associated with neck/shoulder pain.

**Abdominal Pain**: The abundances of Class *Gammaproteobacteria* (P_IVW_ = 3.10E-02, P_Meta_ = 1.95E-02) and Species *Eubacterium eligens* (P_IVW_ = 3.72E-02, P_Meta_ = 1.18E-02) were positively associated with abdominal pain, while that of Genus *Alistipes* (P_IVW_ = 3.46E-02, P_Meta_ = 3.71E-02) was negatively associated with abdominal pain.

**Back Pain**: The abundance of Order *Bifidobacteriales* (P_IVW_ = 3.99E-02, P_Meta_ = 4.83E-02) was positively associated with back pain.

**Pain All Over the Body**: The abundance of Genus *Ruminococcus2* (P_IVW_ = 1.46E-03, P_Meta_ = 2.81E-03) was positively associated with pain all over the body, while Genus *Desulfovibrio* (P_IVW_ = 7.01E-03, P_Meta_ = 1.49E-02) was negatively associated with pain all over the body.

In this exploratory study, we expanded the P-value threshold for the meta-analysis to 0.1. The following associations were observed:

**Back Pain**: The abundance of Family *Prevotellaceae* (P_IVW_ = 1.15E-02, P_Meta_ = 5.20E-02) was negatively associated with back pain.

**Hip Pain**: The abundance of Genus *Subdoligranulum* (P_IVW_ = 4.09E-02, P_Meta_ = 8.41E-02) was negatively associated with hip pain.

**Knee Pain**: The abundance of Genus *Dialister* (P_IVW_ = 3.94E-02, PMeta = 7.69E-02) was negatively associated with knee pain.

### 5.3 Results of BWMR and coloc analysis: GM on MCP and eight pain phenotypes

To further strengthen the causal inference between the identified GM features and chronic pain phenotypes, we employed both BWMR and colocalization analyses. 13 positive GMs were still causally related to MCP and eight pain phenotypes in BWMR, except for Class *Alphaproteobacteria* on neck/shoulder pain (P_BWMR_ = 5.73E-02) (see Table 2).

The PP.H4.abf values of the 13 positive GMs ranged from 0.001691934 to 0.055804256. No data were found in the target region for the remaining three positive GMs. Colocalization analysis findings further strengthened the causal relationship between positive GM and MCP and eight pain phenotypes (See Table 2 and 3, Figure 3 and 4).

### 5.4 Results of MR: GM on BID phenotypes and human cerebral cortex

#### 5.4.1 GM on BID Phenotypes (see Supplementary 10)

##### 5.4.1.1 Data 1

**Genus *Desulfovibrio*:** Its abundance was positively associated with the rostral middle frontal area in the right hemisphere (P_IVW_ = 3.79E-02) and negatively associated with rfMRI connectivity (ICA100 edge 1064) (P_IVW_ = 3.47E-02), for which detailed information was not found.

**Genus *Ruminococcus2***: Its abundance was positively associated with the putamen volume in the right hemisphere (P_IVW_ = 2.94E-02), insula volume in the left (P_IVW_ = 3.19E-02) and right hemispheres (P_IVW_ = 4.72E-02), and G+S-occipital-inf area in the right hemisphere (P_IVW_ = 2.17E-02).

##### 5.4.1.2 Data 2

**Class *Gammaproteobacteria***: Its abundance was positively associated with the medial geniculate nucleus (MGN) volume in the right hemisphere (P_IVW_ = 1.03E-02) and negatively associated with the S-intrapariet+P-trans volume in the left hemisphere (P_IVW_ = 4.87E-02), as well as positively associated with connectivity ICA100 edge 819 (P_IVW_ = 4.90E-02) and negatively associated with connectivity ICA100 edge 1063 (P_IVW_ = 2.89E-02), for which detailed information was not found.

**Family *Prevotellaceae***: Its abundance was positively associated with the weighted mean L1 in trace left posterior thalamic radiation (P_IVW_ = 1.67E-02).

##### 5.4.1.3 Data 3

**Genus *Odoribacter***: Its abundance was positively associated with mean OD in the Fornix crescent+stria terminalis (left) (P_IVW_ = 1.49E-04).

**Genus *Subdoligranulum***: Its abundance was positively associated with the weighted mean L2 in trace right cingulate gyrus part cingulum (P_IVW_ = 3.00E-02).

**Genus *Dialister***: Its abundance was negatively associated with the mean TH of G-pariet-inf-Supramar in the left hemisphere (P_IVW_ = 4.79E-02).

**Order *Bifidobacteriales***: Its abundance was negatively associated with the hippocampal tail volume in the right hemisphere (P_IVW_ = 3.11E-03).

##### 5.4.1.4 Data 4

**Genus *Alistipes***: Its abundance was positively associated with the weighted mean diffusion tensor mode (MO) in the left superior longitudinal fasciculus (P_IVW_ = 3.01E-02) and mean OD in the Fornix crescent+stria terminalis (right) on fractional anisotropy (FA) skeleton (P_IVW_ = 4.94E-02) and negatively associated with the mean TH of the posterior cingulate in the left hemisphere (P_IVW_ = 1.49E-02), mean MO in the inferior cerebellar peduncle (left) on FA skeleton (P_IVW_ = 4.72E-02), corticoamygdaloid transition volume in the right hemisphere (P_IVW_ = 1.12E-03), and medial orbitofrontal cortex volume in the left hemisphere (P_IVW_ = 4.02E-03).

**Genus *Bifidobacterium***: Its abundance was positively associated with connectivity (ICA100 edge 405) (P_IVW_ = 2.31E-02), for which detailed information was not found, and negatively associated with the mean TH of the parstriangularis (P_IVW_ = 1.69E-03) and S-circular-insula-sup (P_IVW_ = 4.09E-03) in the left hemisphere; mean TH of the G-insular-short in the right hemisphere (P_IVW_ = 3.36E-03); mean OD in the body of the corpus callosum (P_IVW_ = 3.06E-02) and Fornix crescent+stria terminalis (right) on the FA skeleton (P_IVW_ = 4.47E-02); HATA volume (P_IVW_ = 5.89E-03) and area of isthmus cingulate (P_IVW_ = 2.66E-03) in the left hemisphere; G-front-middle volume (P_IVW_ = 4.58E-02), S-circular-insula-inf volume (P_IVW_ = 4.85E-02), and area of superior temporal (P_IVW_ = 2.95E-02) in the right hemisphere; and rfMRI amplitudes (ICA100 node 27) (P_IVW_ = 5.89E-03), for which detailed information was not found.

#### 5.4.2 GM on the Human Cerebral Cortex (see Supplementary 11)

##### 5.4.2.1 Data 1

**Genus *Desulfovibrio***: Its abundance was positively associated with mean SA of the paracentral (P_IVW_ = 4.57E-02) and mean TH of the frontal pole (PIVW = 2.60E-02) and negatively associated with mean SA of the superior parietal (P_IVW_ = 3.48E-02).

**Genus *Ruminococcus2***: Its abundance was positively associated with mean SA of the bankssts (P_IVW_ = 3.83E-02) and fusiform (P_IVW_ = 1.61E-02).

**Phylum *Verrucomicrobia***: Its abundance was negatively associated with mean SA of the lateral orbitofrontal (P_IVW_ = 3.04E-02).

##### 5.4.2.2 Data 2

**Class *Gammaproteobacteria***: Its abundance was positively associated with mean SA of the bankssts (P_IVW_ = 4.80E-02), paracentral (P_IVW_ = 4.47E-02), posterior cingulate (P_IVW_ = 1.97E-03), and precentral (P_IVW_ = 2.27E-02) and negatively associated with the mean TH of the isthmus cingulate (P_IVW_ = 4.04E-02).

**Family *Prevotellaceae***: Its abundance was negatively associated with mean TH of the inferior parietal (P_IVW_ = 2.33E-03).

##### 5.4.2.3 Data 3

**Genus *Odoribacter***: Its abundance was positively associated with the mean TH of lateral occipital (P_IVW_ = 3.22E-02) and negatively associated with the mean TH of superior frontal (P_IVW_ = 3.46E-02).

**Genus *Subdoligranulum***: Its abundance was negatively associated with the mean TH of the rostral middle frontal cortex (P_IVW_ = 4.19E-02).

**Genus *Dialister***: Its abundance was positively associated with the mean TH of middle temporal (P_IVW_ = 4.16E-02) and negatively associated with the mean SA of inferior parietal (P_IVW_ = 4.91E-02) and precentral (P_IVW_ = 4.46E-02).

**Species *E. eligens***: Its abundance was positively associated with the mean SA of posterior cingulate (P_IVW_ = 3.02E-02).

**Phylum *Verrucomicrobia***: Its abundance was positively associated with global full SA (P_IVW_ = 4.83E-02) and the mean SA of caudal middle frontal (P_IVW_ = 3.77E-02), insula (P_IVW_ = 8.68E-03), superior temporal (P_IVW_ = 3.40E-02), and TH of postcentral (P_IVW_ = 2.87E-02) and rostral anterior cingulate (P_IVW_ = 2.91E-02) and negatively associated with the mean SA of pericalcarine (P_IVW_ = 3.32E-02).

**Genus *Adlercreutzia***: Its abundance was positively associated with the mean SA of lateral occipital (P_IVW_ = 3.39E-02).

##### 5.4.2.4 Data 4

**Genus *Alistipes***: Its abundance was positively associated with the mean SA of lateral occipital (P_IVW_ = 2.61E-02) and negatively associated with the mean SA of fusiform (P_IVW_ = 1.66E-03) and postcentral (P_IVW_ = 4.62E-02).

**Genus *Bifidobacterium***: Its abundance was positively associated with the mean TH of posterior cingulate (P_IVW_ = 4.32E-02) and negatively associated with the mean SA of supramarginal (P_IVW_ = 1.83E-03) and transverse temporal (P_IVW_ = 7.67E-03).

#### 5.4.2 GM on 83 Brain Regions (see Supplementary 12)

**Class *Gammaproteobacteria***: Its abundance was negatively associated with the left superior parietal volume (P_IVW_ = 3.07E-02).

**Genus *Bifidobacterium***: Its abundance was negatively associated with the right caudal middle frontal volume (P_IVW_ = 1.97E-02). No causal effects were observed for the other 13 genetic predispositions on 83 brain regions because their P_IVW_s were >0.05.

### 5.5 Results of mediation analysis

#### 5.5.1 GM on MCP and eight pain phenotypes (See Table 4, Fig 5, Supplementary 13)

Finally, we conducted a mediation analysis to explore the primary mediating GBA model of MCP and eight pain phenotypes based on Sections 5.2 and 5.3. Our findings indicate that the causal effect between Genus *Odoribacter* and neck/shoulder pain could be mediated by mean OD in Fornix cres + Stria terminalis (left) (Table 4). For Genus *Odoribacter* - mean OD in Fornix cres+Stria terminalis (left) - neck/shoulder pain, TE = −0.020, IE = −0.003, DE = −0.017 (P = 0.027), IE_div_TE = 0.140 (0.013-0.267) (see Figure 5).

**Table 4.**
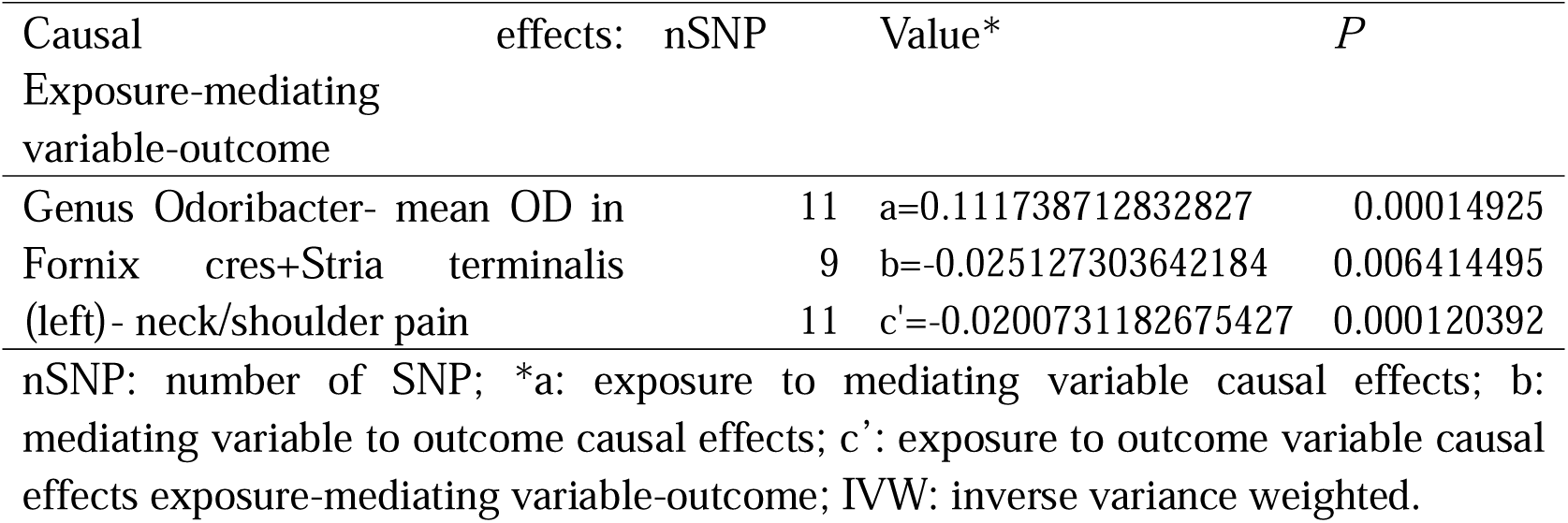
Three causal effects (a, b, c’) of the mediation analysis in IVW method.

**Figure 5.**
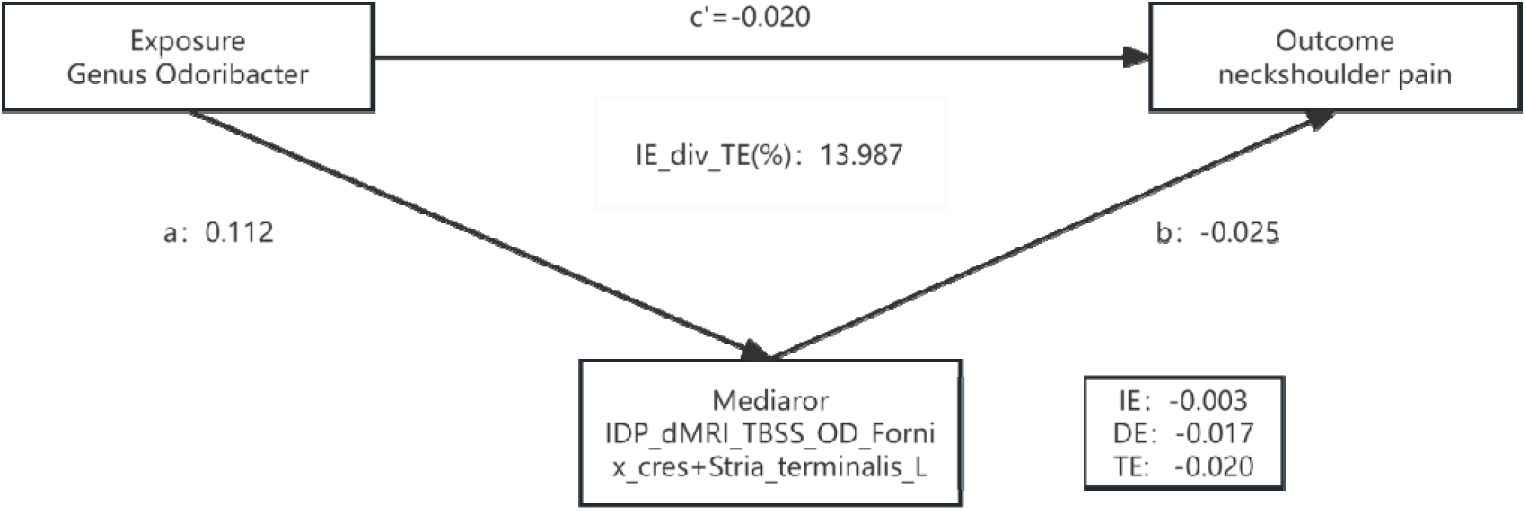
Mediator model: Genus Odoribacter to neckshoulder pain

## 6. Discussion

### 6.1. GM and pain phenotypes

This study is the first to analyze the causal association of GM with MCP and eight pain phenotypes by MR, incorporating human brain structures. Furthermore, we were able to establish a GBA model linking Genus *Odoribacter* to neck/shoulder pain. We identified 13 potential relevant GM. Importantly, different pain phenotypes were causally associated with specific GM. Seven positive and eight negative causal associations were found, among which Genus *Odoribacter* as a protective GM for neck/shoulder pain needs to be emphasized.

Previous MR studies did not identify GM overlapping with those within our study, except for Family *Prevotellaceae*. This discrepancy may stem from previous studies predominantly focusing on specific pain-related diseases as outcomes, while our study examined pain phenotypes. Notably, our study suggests that Family *Prevotellaceae* provides protection against back pain, whereas previous MR studies reported it increases the risk of low back pain [23,78]. This difference could be attributed to variations in study outcomes and methodologies; previous studies often utilized single GM databases, whereas we incorporated four different GM databases and conducted a meta-analysis.

Meanwhile, our results suggest Phylum *Verrucomicrobia* and Genus *Dialister* are protective factors for CP, while Genus *Adlercreutzia* is risk factor, consistent with previous observational studies [18,23,30,40–41,85]. Despite variations in associated pain sites, this reinforces the potential role of these GMs in pain pathogenesis. Conversely, previous observational studies involving Genus *Desulfovibrio*, Genus *Subdoligranulum*, and Genus *Odoribacter* have yielded inconsistent results [17,23,86–88]. Our study’s findings may be more compelling due to its ability to establish causation, which observational studies cannot confirm definitively as they often have small sample sizes.

Interestingly, despite its probiotic properties in inhibiting pain behaviors [55], Genus *Bifidobacterium* was found to increase the risk of facial pain in our study, aligning with previous inconsistent findings [23,54,59]. Similarly, Genus *Adlercreutzia*, recognized for its health benefits [10], has been associated with back pain [18] and identified as a risk factor for MCP in our study. A recent attempt to use *Bifidobacterium* in treating neuropathic pain was unsuccessful [28], suggesting that the effects of symbionts may be complicated through interactions with pathobionts that are associated with pain conditions. The role of GM in CP is complex, possibly involving differential effects within microbial networks [10]. Future research may benefit from constructing gut microbial networks based on key GMs to elucidate CP mechanisms.

### 6.2. The gut-brain axis

Pain is a dynamic interplay between sensory, emotional, and cognitive factors [48] processed within the pain matrix-a network of interconnected brain regions [15], especially CP. Emerging research suggests a potential influence of GM in modulating pain perception [45]. Since changes in brain structure can significantly affect its function [57], we investigated the potential connection between GM and brain regions involved in pain processing using brain imaging data. In general, except for Family *Prevotellaceae*, Genus *Subdoligranulum*, Order *Bifidobacteriales*, and Species *E. eligens*, the remaining positive GMs have been potentially causally linked to the brain regions primarily involved in sensory, emotional, and cognitive functions, which may help us partly understand how GM affect CP.

Firstly, we found that multiple positive GM could affect the area of pain-related cerebral cortex. In our study, the abundance of Phylum *Verrucomicrobia,* a protective factor against MCP and headache, was not only positively associated with the entire cortical area, but also positively correlated with the area of caudal middle frontal and insula. This finding extends the results of a recent study on CP utilizing the UK Biobank data [5]. Other studies suggest that the anterior insula plays a more important role in anticipating pain and its emotional features, while the posterior insula is more active during the physical sensations of pain [55, 58, 68]. The observed protective effect of Phylum *Verrucomicrobia* on these regions aligns with the possibility that these bacteria might help in maintaining normal brain function related to pain perception. This finding offers a potential target for future research on chronic pain management strategies.

Furthermore, our study revealed that Genus *Desulfovibrio* was not only positively correlated with the area of rostral middle frontal cortex which are crucial for emotional and cognitive processing, but also positively correlated with the paracentral area and negatively with the superiorparietal area belonging sensorimotor cortices. As a protective factor for chronic multisite pain, the interaction between these affected brain areas deserves further exploration.

Secondly, we found that multiple positive GM could affect the thickness and volume of pain-related cerebral cortex. CP patients often show reduced gray matter volume in several brain regions, including areas involved in processing pain, managing emotions, and controlling movement [3–4,6,20,32,35,46–47,51–52,64,70–71,73,91]. Certain GM may have either positive or negative effects on the volume of these brain regions. For example, Phylum *Verrucomicrobia* abundance was associated with thicker gray matter in the rostral-anterior-cingulate and postcentral regions, further supporting its protective role against CP. Additionally, multiple GM were found to significantly influence one brain region, such as the frontal lobe. This finding suggests that the observed decrease in gray matter volume in CP patients may result from the combined effects of multiple GM.

Thirdly, this study reveals that GM may influence chronic pain by affecting the integrity and direction of axonal fiber dispersion in white matter. CP patients often exhibit changes in the microstructure of white matter, like superior longitudinal fasciculus, corpus callosum, and cingulum bundle, which play crucial roles in pain severity, resilience and emotional response to pain [52,69,81,89]. Our results indicate that certain positive GM can positively (Genus *Subdoligranulum-*cingulum bundle (rh)) or negatively (Genus *Bifidobacterium*-corpus callosum) influence white matter in specific locations. Observational studies have also linked reduced white matter integrity in the splenium of the corpus callosum to chronic low back pain disability [12]. Our study found that Family *Prevotellaceae* abundance positively affected posterior-thalamic-radiation (lh),supporting its role as a protective factor against back pain.

Morphological changes in the limbic system are closely linked to persistent pain. Particularly, the Fornix cres+Stria terminalis region needs to be emphasized, which is positively affected by Genus *Odoribacter* and Genus *Alistipes* and negative affected by Genus *Bifidobacterium*. Notably, our findings suggest that the causal relationship between Genus *Odoribacter* abundance and neck/shoulder pain may be mediated by mean OD in the Fornix cres+Stria terminalis (left).

The fornix, crucial for normal cognitive function [80], has also been associated with pain abnormalities [43]. Similarly, the stria terminalis is integral to the limbic system, connecting the amygdala to the hypothalamus. Although the fornix and stria terminalis are not directly connected, they create an indirect anatomical pathway between the hippocampus and the amygdala. Through shared connections to structures like the hypothalamus and septal nuclei, these regions can influence each other, forming a corticolimbic system critical in the initiation, persistence, and amplification of chronic pain states [1,83], and influencing the transition from acute to chronic pain [91]. Continued activation of corticolimbic circuits results in functional and anatomical changes in the cortex, contributing to prolonged pain states [91].

Our study identified new avenues for investigating the GBA in chronic pain and pain phenotypes. The Fornix cres+Stria terminalis is highlighted as a new factor in the GBA and a novel key area for CP (Genus *Odoribacter* to neck/shoulder pain). Targeted therapies focusing on these areas may offer innovative approaches to pain management and treatment.

There are some contradictions in the results: Genus *Alistipes* appears to exert contrasting influences in abdominal and facial pain. Class *Gammaproteobacteria* abundance was positively correlated with the paracentral and precentral areas but acted as a risk factor for abdominal pain, and Genus *Dialister* abundance was negatively associated with the precentral area but served as a protective factor for knee pain. Various internal brain networks and regulatory systems collaboratively contribute to the experience and persistence of chronic pain [16,19,46]. Future research using multimodal fMRI to investigate how GM influences these brain network changes may provide insights into these findings.

### Limitations and strengths

Our study has some advantages. First, we employed a comprehensive set of sensitivity analyses, including meta-analyses and colocalization analyses. These analyses provide strong evidence that our findings are robust. Second, we addressed potential biases by applying methods like MR-Egger and MR-PRESSO to assess pleiotropy (where a genetic variant influences multiple traits) and identify outliers. Those steps helped to ensure the reliability of our MR results. Finally, the analysis utilized powerful IVs (F-statistics > 10), minimizing the influence of weak instruments that could bias the MR results.

However, our study also has certain limitations. Firstly, the data originated from participants of European ancestry. Secondly, we did not conduct reverse MR analyses or MR analyses directly analyzing the relationship between pain phenotypes and brain structure, which will be explored in future studies. Thirdly, to ensure a sufficient number of IVs for GM, we employed a significance threshold of P<1×10^-5^, which is less stringent than the genome-wide significance level (P<5×10^-8^).

## Conclusions

Among European ancestry, we identified 13 GM that were causally related to chronic pain; 8 GM decreased risk towards MCP and pain phenotypes, and 7 GM increased risk. Core GM may influence CP by affecting complex brain networks associated with emotional, cognitive, and motor functions. Specifically, we constructed a basic GBA model for neck/shoulder pain with Genus *Odoribacter* - Fornix cres+Stria terminalis (left) interactions playing a key role in neck/shoulder pain. Our study contributes to novel insights into the relationships between specific GM and pain phenotypes, supporting that GBA play a key role in the pathogenesis of CP. Consequently, the GBA is a new drug target to treat CP through neuroregulatory therapies and targeted treatments with protective symbionts.

## Data Availability

All relevant data are within the manuscript and its Supplementary Material.

## Acknowledgements

We wish to acknowledge the participants and investigators of the UK Biobank, FINRISK 2002 study, Enhancing NeuroImaging Genetics through Meta-Analysis Consortium, the Integrative Epidemiology Unit (IEU, https://gwas.mrcieu.ac.uk/), the GWAS Catalog (https://www.ebi.ac.uk/gwas/).

